# Deep Learning on Histopathological Images to Predict Breast Cancer Recurrence Risk and Chemotherapy Benefit

**DOI:** 10.1101/2025.05.15.25327686

**Authors:** Gil Shamai, Shachar Cohen, Yoav Binenbaum, Edmond Sabo, Alexandra Cretu, Chen Mayer, Iris Barshack, Tal Goldman, Gil Bar-Sela, António Polónia, Frederick M. Howard, Alexander T. Pearson, Dezheng Huo, Joseph A. Sparano, Ron Kimmel, Dvir Aran

## Abstract

Genomic testing has transformed treatment decisions for hormone receptor-positive, HER2-negative (HR+/HER2-) early breast cancer; however, it remains inaccessible to many patients worldwide due to high costs and logistical barriers. Here, we developed an artificial intelligence (AI) model using a multimodal deep learning approach that estimates Oncotype DX 21-gene recurrence scores (RS) from routine histopathology images and clinicopathologic variables, including age at diagnosis, tumor size, and receptor status. Using a foundation model pre-trained on 171,189 histopathological slides, we fine-tuned and validated our AI model on the TAILORx randomized trial (n=8,284). Among 2,407 patients in the TAILORx validation, the model classifies 45.6% of patients as low-risk, 42.4% as intermediate risk, and 12.0% as high-risk. For predicting high genomic risk disease (RS≥26), occurring in 15.9% in the TAILORx validation set, the model achieves AUC=0.898. Patient stratification by our model shows strong prognostic value across multiple clinical endpoints, including recurrence-free interval, distant recurrence-free interval, and disease-free survival. Importantly, chemotherapy benefit is demonstrated for premenopausal patients classified by our model as high AI risk and chemotherapy benefit is ruled out for postmenopausal patients classified as low AI risk. External validation across six independent cohorts (n=5,497 patients) demonstrates robust generalization of the AI model for prognostication and prediction of RS. Notably, in postmenopausal patients, the AI model reclassifies approximately 30% of clinically high-risk cases, defined by the MINDACT criteria, as low-risk. These findings demonstrate that artificial intelligence applied to standard histopathology can be a valuable tool for chemotherapy decision-making in HR+/HER2- early breast cancer. This approach can help reduce unnecessary chemotherapy and extend precision medicine, particularly in resource-limited settings, where genomic testing is not widely accessible.

## Main

Breast cancer is the most frequently diagnosed cancer worldwide^1^. For patients with hormone receptor-positive, HER2-negative (HR+/HER2-) early-stage breast cancer, which accounts for approximately 70% of cases, a critical challenge is identifying those who will benefit from adjuvant chemotherapy^2^. Treatment decisions were traditionally based on clinical features such as tumor size, receptor expression, grade, and node involvement, but these criteria alone lack sufficient accuracy for optimal patient stratification^3^. Consequently, some patients may have received chemotherapy without deriving clear benefits, while others who could have benefited from systemic therapy may have been missed.

The development of multigene expression assays has revolutionized treatment decision-making in HR+/HER2-early breast cancer^4–7^. Among these, the Oncotype DX 21-gene recurrence score (RS) assay is the only test recommended by the National Comprehensive Cancer Network (NCCN) guidelines as predictive of chemotherapy benefit^8,9^. The initial validation studies, NSABP B-20 and SWOG-8814, demonstrated significant chemotherapy benefit in patients with high RS (≥31)^6,10^. Subsequent reanalysis of the B-20 cohort, excluding patients with HER2-positive disease, indicated that RS≥26 provided a more appropriate threshold for the prediction of chemotherapy benefit^11,12^. The subsequent landmark prospective TAILORx trial, which enrolled 10,273 women with HR+/HER2- node-negative disease, was one of the largest randomized trials in breast cancer^13^. Using updated thresholds now standard in clinical practice (low < 11, intermediate 11–25, and high > 25), the TAILORx trial found that endocrine therapy alone was as effective as chemoendocrine therapy for patients with mid-range RS (11–25), whereas the substantial benefit of chemotherapy was found and is recommended by current guidelines for RS≥26^9,11^. A posthoc analysis of TAILORx indicated some chemotherapy benefit for node-negative premenopausal women or those 50 or younger with 16≤RS<26, with the magnitude of that benefit increasing with increasing clinical risk. For women with HR+/HER2- breast cancer and 1-3 positive axillary nodes, the RxPONDER trial showed that premenopausal women with low genomic risk breast cancer (RS<26) derived some chemotherapy benefit, whereas no chemotherapy benefit was observed in postmenopausal women^9,14–16^.

Despite these advances and strong recommendations in clinical practice guidelines, the global adoption of genomic testing remains limited^17–19^. While adoption exceeds 70% in the United States and approaches universal coverage in Israel through national health programs, these rates are significantly lower elsewhere. In Europe, reimbursement policies vary widely, with recent expansion in Western European countries like Germany, France, and the UK, but more limited implementation in Eastern regions^20^. In resource-limited settings, utilization remains minimal - approximately 5% in India, below 10% across most of Asia, and negligible throughout most of Africa^19^. The primary barriers include high cost (approximately $4,000 per test), extended turnaround times (2-3 weeks), and logistical challenges such as sample shipping and reimbursement procedures, despite potential downstream savings from more tailored therapy^21^.

Hematoxylin and eosin (H&E) staining, a fundamental method in pathology, is widely accessible and routinely applied to solid tumors for assessing tissue morphology. With the increasing availability of digital pathology, including in resource-limited settings^22^, artificial intelligence (AI) approaches offer promising alternatives. Significant progress has been made in analyzing H&E-stained whole-slide images (WSIs) to diagnose tumors^23^, predict receptor status^24–29^, identify genetic mutations^30–32^, and predict patient survival and disease-specific outcomes^33–36^. Several studies have shown that high Oncotype DX RS can be identified from H&E images and clinicopathologic variables^37–43^. However, none of these studies validated their models using high quality randomized clinical trial data, a critical step to address treatment related confounding in observational settings and assess the model’s ability to predict chemotherapy benefit. This validation is especially important given the primary goal of the Oncotype DX assay to predict chemotherapy benefit. Moreover, these approaches typically had limited validation, both in the size and number of independent cohorts and, importantly, in the extent of external validation and generalizability across different datasets. External validation is especially critical given the sensitivity of deep learning models to dataset variations, which often necessitates calibration or fine tuning.

Recent advances in deep learning have dramatically improved the analysis of histological images through large-scale foundation models^44–46^. These models incorporate self-supervised learning techniques and transformer architectures. Transformers^47^, originally developed for natural language processing, excel in capturing complex spatial patterns and long-range dependencies in histological slides, outperforming traditional convolutional neural networks. Self-supervised learning methods allow models to learn salient image features without relying on manual annotations, greatly expanding the diversity and volume of available training data^48–50^. By learning robust, general-purpose image representations, foundation models demonstrate high performance in histopathology and are particularly strong in generalizability to external data.

Here, we present a multimodal deep learning approach for inferring the Oncotype DX 21-gene RS from H&E images and clinicopathologic features, leveraging data from TAILORx. Our study makes several key contributions. First, the TAILORx trial dataset provides a unique opportunity to validate our predictions against patient outcomes and chemotherapy benefit and compare them directly to Oncotype DX RS on a large randomized trial (**Figure 1a**). Second, our extensive data collection consists of 25,436 H&E slides from 17,812 patients across seven independent cohorts, six of which are external, representing diverse data used to validate the model in a real-world setting (**Figure 1b**). Third, our method leverages a foundation model^45^, pre-trained on 171,189 histological slides, which enables robust feature extraction despite variations in slide preparation and scanning protocols across institutions (**Figure 1c**). Fourth, to overcome variations in data across cohorts and enable practical deployment, particularly in resource-limited settings, we applied a calibration method that does not require local genomic testing data. Finally, and most importantly, we evaluated how our model’s predictions align with observed chemotherapy benefit across premenopausal and postmenopausal subgroups, and assessed how frequently the model reclassified patients’ risk status relative to standard clinical risk criteria, which is highly relevant when genomic tests are inaccessible. Our results suggest that the AI multimodal model based on deep learning analysis of H&E images and clinicopathologic covariates provides immediate risk stratification for a substantial proportion of patients, potentially expanding access to precision oncology globally and impacting many of the patients diagnosed with breast cancer each year.

**Figure 1:**
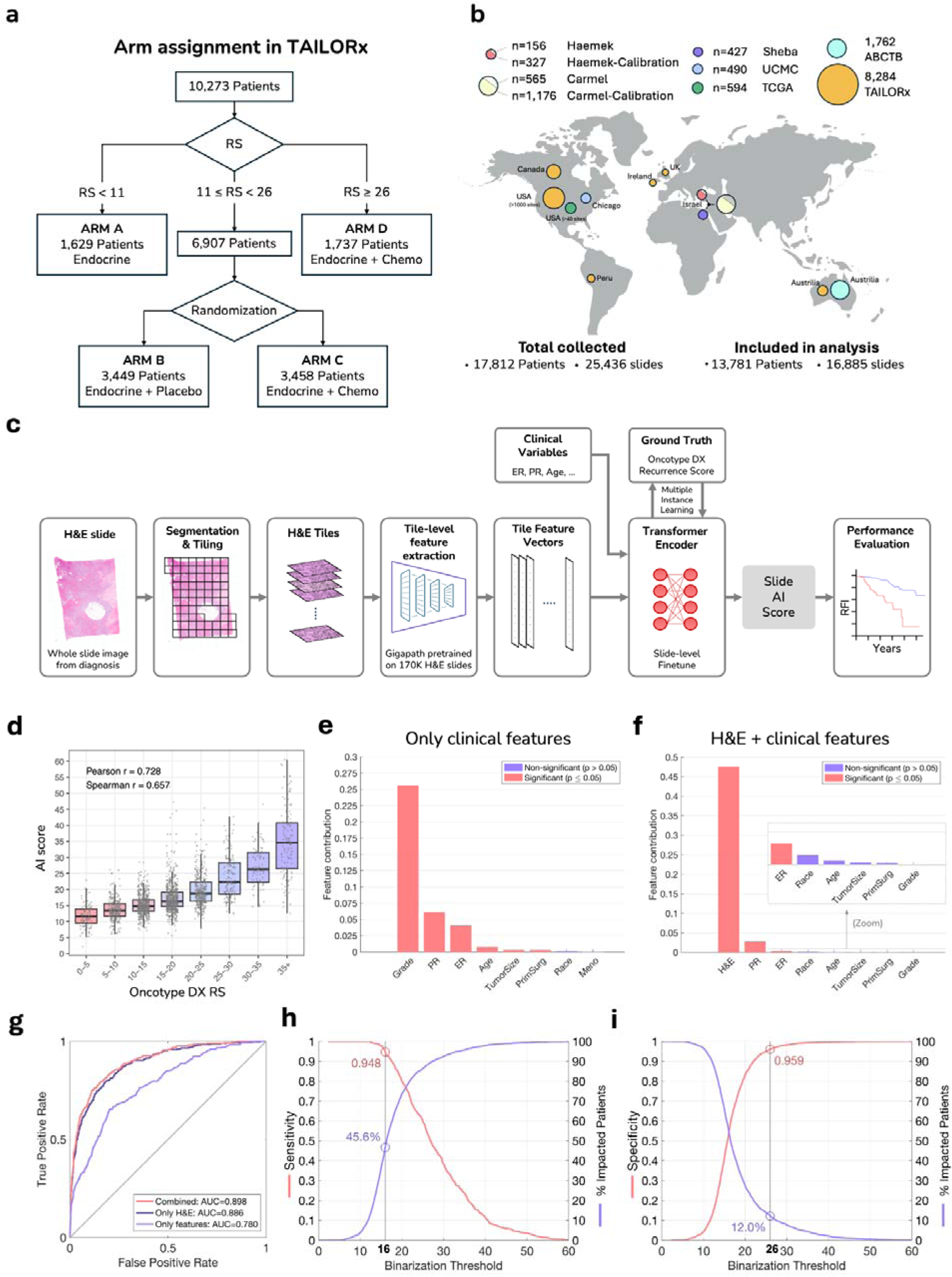
Study cohorts, deep learning system architecture, and performance evaluation on TAILORx-validation. **(a)** Design of the TAILORx trial, showing patient stratification by recurrence score (RS) and randomization of intermediate-risk patients (RS 11-25) to endocrine therapy alone or with chemotherapy. **(b)** Geographic distribution of study cohorts. TAILORx served as the primary tuning and validation cohort. External validation was performed on six independent cohorts: Carmel, Haemek, Sheba, UCMC, ABCTB, and TCGA. Additional patient subsets from Carmel and Haemek were used for calibration experiments. Exclusions were limited to slides with corrupted scans or insufficient tissue for analysis (Supplementary Figure 1). **(c)** Pipeline of the deep learning system: H&E slides undergo automated tissue segmentation and tiling, followed by feature extraction using the Gigapath foundation model pre-trained on 171,189 H&E slides via self-supervision. Combined with clinicopathologic variables, these features are then used for tuning a transformer-based multiple-instance learning model to generate slide-level scores. **(d)** Distribution of the AI scores versus Oncotype DX RS on the TAILORx held-out validation data, showing high correlation (Pearson r=0.728, Spearman r=0.657). **(e-f)** Quantitative assessment of feature contributions to RS estimation using sequential forward selection, for the model using only clinical features (e) and the AI multimodal model incorporating both H&E images and clinical features (f). Clinical features evaluated include: histological grade, hormone receptor status (PR, ER), patient age, tumor size (dichotomized at 2cm), surgery type (mastectomy/lumpectomy), self-reported race, and menopausal status. Bar heights represent additive R-squared values, with statistical significance indicated. **(g)** Receiver operating characteristic (ROC) curves for identifying high genomic risk (RS≥26) for the three models: the AI model combining both H&E and clinical features (AUC=0.898), the image model using only H&E images (AUC=0.886), and the clinicopathologic model using clinical features alone (AUC=0.780). **(h)** Using a low threshold of 16, the model classifies 45.6% of the patients as low AI risk. Sensitivity is measured for the task of identifying RS≥26. **(i)** Using a high threshold of 26, the model classifies 12.0% of the patients as having high AI risk. Specificity is measured for the task of identifying RS≥26.

### Development and Validation Using TAILORx Clinical Trial Data

From the TAILORx trial, we included 9,383 whole-slide images from 8,284 patients with node-negative HR+/HER2- early breast cancer who had digitized H&E slides from diagnosis (**Figure 1b, Supplementary Table 1**). Exclusions were limited to slides with corrupted scans or insufficient tissue for analysis (**Supplementary Figure 1)**. Patients were enrolled on the trial between 2006 and 2010; among the subset of 8,284 included in this analysis (80.6% of the trial population), median age was 56 years, 31% were 50 or younger, 34% were premenopausal, median tumor size was 1.5cm, 18% had poor histologic grade, 17% had high RS (≥26), and 70% had low clinical risk by the MINDACT criteria (**Supplementary Table 2**). Patients included in the analysis were randomly split into a tuning set (70%, 5,877 patients) and a validation set (30%, 2,407 patients). The tuning set was used to fine-tune the pre-trained foundation model to infer the Oncotype DX RS from H&E slides and clinicopathologic variables in five-fold cross-validation. Clinicopathologic variables included age at diagnosis (continuous), menopausal status, tumor size (dichotomized at 2cm), histologic grade, ER and PR status by immunohistochemistry, type of surgery (mastectomy/lumpectomy), and self-reported race. During cross-validation, hyperparameter tuning was limited to fixing the number of epochs at 3 for all models. After cross-validation, the models were validated using the held-out validation set.

For tuning and inference, each digitized H&E image underwent automated tissue detection and segmentation into tiles, followed by feature extraction using a deep learning foundation model developed for H&E analysis (**Figure 1c**). Our approach uses the GigaPath foundation model^45^ that was pre-trained on 171,189 histopathological slides using self-supervised learning, enabling robust feature extraction from tile images without requiring manual annotations. In the downstream task, the model was tuned using transformer-based multiple instance learning to predict Oncotype DX RS from the tile features, enabling the integration of information from multiple regions of the tissue simultaneously. The clinicopathologic variables were integrated into the tuning process, resulting in a multimodal model (‘the AI model’).

At inference, the AI model achieved a correlation of 0.728 (p<0.001) on the validation set (**Figure 1d**). To better understand the contribution of different features to Oncotype DX RS prediction, we conducted a feature importance analysis, evaluating both the AI model and a similar model using only the clinicopathologic variables (**Figure 1e-f**). When using clinical variables alone, grade contributed the most, followed by progesterone receptor (PR) status, estrogen receptor (ER) status, age, and tumor size. However, when H&E images were incorporated, the image features made the predominant contribution. PR and ER status still added significant contributions, while other clinical features, including grade, showed negligible additional contributions. This suggests that morphological characteristics captured in the H&E images already encode most of the information provided by histological grade.

When evaluated for the clinically important task of identifying high genomic-risk disease (RS≥26), the AI model had an AUC performance of 0.898 (**Figure 1g and Supplementary Table 3a**). We further compared the AI model to the clinicopathologic model based only on clinical variables, and an image model based only on the H&E images, which were trained and validated similarly to the multimodal model. The clinicopathologic model achieved an AUC of 0.780 on the validation set, and the image model achieved an AUC of 0.886. The AI model using both images and clinicopathologic variables significantly improved performance over the model using only H&E images (P<0.001), as well as over the model using only clinicopathologic variables (P<0.001).

To make the model translational, we leveraged the continuous AI model scores to establish classification thresholds that could enable therapeutic decisions (**Figures 1h-i and Supplementary Table 3b-c**). Using an AI low-risk threshold of 16 and an AI high-risk threshold of 26, the AI model classifies 45.6% of the patients as low AI risk, 42.4% as intermediate AI risk, and 12.0% as high AI risk. Classification of the patients using the low-risk threshold resulted in a sensitivity of 0.948 and a negative predictive value (NPV) of 0.982 against RS≥26. This means that only 1.8% of patients classified as low AI risk had high genomic risk (RS≥26). Classification using the high AI risk threshold resulted in specificity=0.959 and positive predictive value (PPV)=0.716 for identifying high genomic risk (RS≥26), favoring the utilization of chemotherapy or CDK 4/6 inhibitors such as ribociclib for these patients. Our analysis shows that for patients classified as high AI risk or low AI risk, consisting of 57.6% of the cohort, the model was able to accurately identify if their genomic risk was high or not, respectively. Performance on the validation set was consistent with performance during cross-validation (**Supplementary Figure 2a-d and Supplementary Table 3**), indicating no overfitting occurred during the tuning process.

For clarity and brevity, throughout the remainder of this paper, we refer to the AI multimodal model simply as ‘the model’ or ‘the AI’, and to Oncotype DX RS simply as ‘RS’.

### Risk Stratification by the AI Model and by the Genomic RS Demonstrates Robust Concordance

Comparing the AI scores to RS is insufficient as a standalone measure; correlation with clinical outcomes must also be established to validate the model’s performance. To evaluate the ability of the AI model to provide prognostic information regarding patient outcomes, we analyzed long-term outcomes in the TAILORx cohort (**Figure 2)**. When stratifying patients using an AI threshold of 26, the survival curves on AI scores closely mirrored those based on RS. This concordance was maintained across multiple clinical endpoints: distant recurrence-free interval (DRFI), recurrence-free interval (RFI), and disease-free survival (DFS; defined as freedom from invasive disease recurrence, second primary cancer, or death) across TAILORx-validation patients (**Figure 2a**), with comparable hazard ratios between the AI scores (DRFI HR=2.9, RFI HR=2.6, DFS HR=1.3) and RS (DRFI HR=2.6, RFI HR=2.4, DFS HR=1.3). A similar pattern was observed when including the cross-validation patients (**Supplementary Figure 3a**), as well as for additional endpoints of overall survival and breast cancer-specific survival (**Supplementary Figure 3b**).

**Figure 2:**
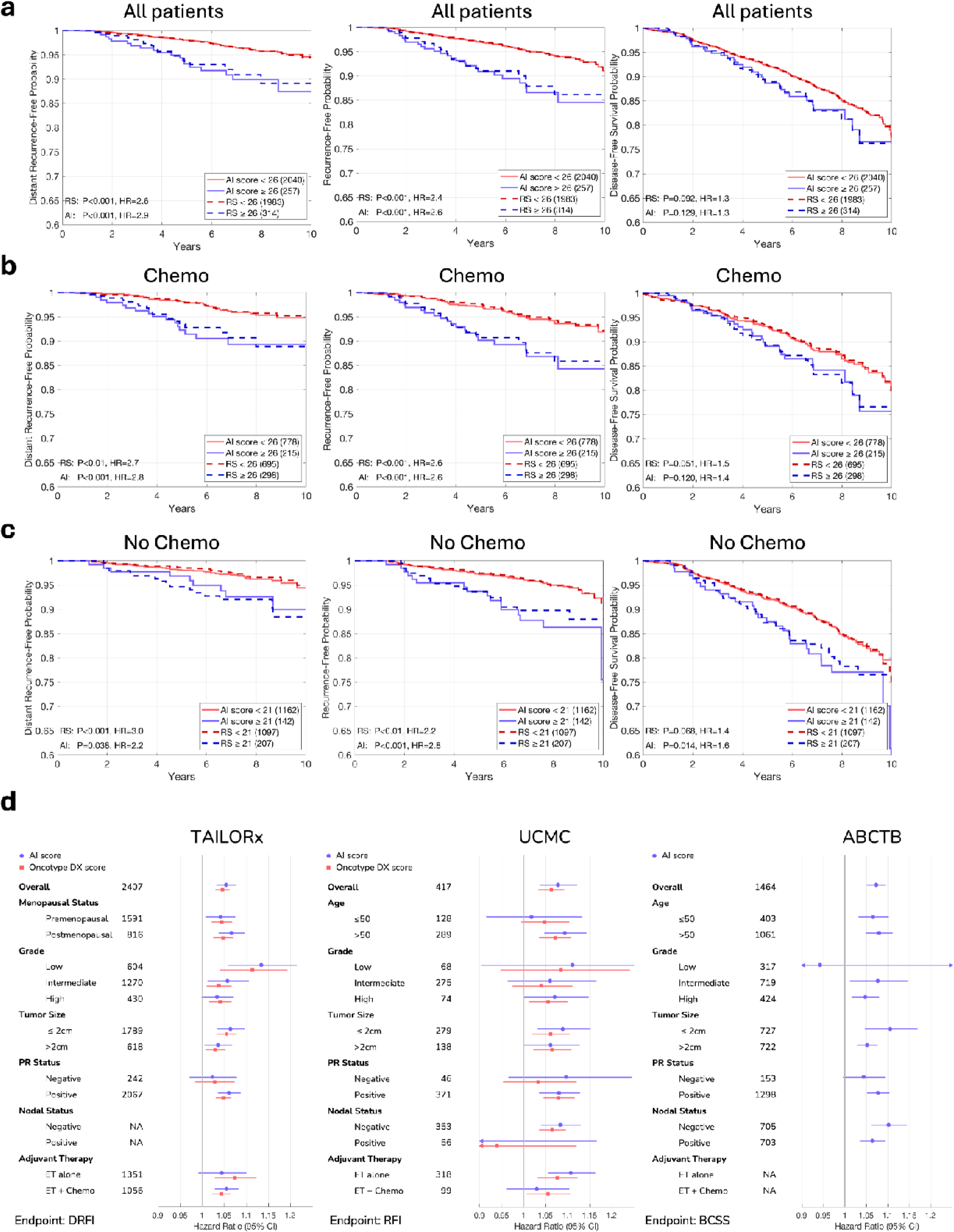
Survival analysis comparing RS and AI for prognostication in TAILORx-Validation. **(a)** Kaplan-Meier plots comparing patient stratification using RS versus AI scores across TAILORx validation patients, using distant recurrence-free, recurrence-free, and disease-free survival probabilities and a strat fication threshold of 26. **(b)** Analysis of the same endpoints in patients who received chemotherapy. **(c)** Corresponding analysis for patients who did not receive chemotherapy, for a lower classification threshold of 21. For each subgroup analysis, patient numbers, P values for the stratification, and hazard ratios (HR) are shown in the legends. The consistent alignment between the survival curves derived from RS and AI scores across all analyses demonstrates the robust ability of the AI model to infer patient outcomes. **(d)** Forest plots comparing hazard ratios with 95% confidence intervals for the AI scores and RS across multiple patient subgroups in three cohorts with long-term outcome data. Analysis shows HRs per unit increase in the score (continuous variable) for inferring distant recurrence-free interval (DRFI) in the TAILORx validation set, recurrence-free interval (RFI) in UCMC, and breast cancer-specific survival (BCSS) in ABCTB. The number of patients in each subgroup is indicated. Note that TAILORx included only node-negative patients and that ABCTB lacked RS data, showing only AI prognostic performance. ET: endocrine therapy.

We further evaluated the stratification performance in patient subgroups based on treatment. For patients who received chemotherapy, stratifications based on the AI scores and those based on RS maintained strong concordance across all endpoints (**Figure 2b, Supplementary Figure 3c**), with similar hazard ratios. For patients who did not receive chemotherapy, where accurate risk assessment is particularly crucial, the AI and RS again demonstrated similar trends (**Figure 2c, Supplementary Figure 3d).** For this analysis, we used a lower classification AI threshold of 21, since patients with RS≥26 were all assigned to receive chemotherapy. Other stratification thresholds resulted in similar trends (**Supplementary Figure 4**).

The prognostic performance of the AI model demonstrated hazard ratios comparable to those of RS across clinical subgroups, including menopausal status, tumor grade, tumor size, PR status, and treatment, as measured by distant recurrence-free interval (**Figure 2d**). This subgroup analysis is particularly relevant for premenopausal and postmenopausal patients, given that post-hoc analyses of the TAILORx trial revealed distinct chemotherapy benefit profiles between these groups^16^.

It is important to interpret the AI model’s prognostic performance with caution, particularly when compared to RS since arms in TAILORx were assigned based solely on RS scores–not AI scores–potentially introducing bias. Nevertheless, these analyses across multiple endpoints and patient subgroups suggest that the AI model, based on routinely available H&E slides and pathology data, captures biological signals consistent with those assessed by the established Oncotype DX genomic testing.

### The AI Score is Predictive for Chemotherapy Benefit in Pre- and Post-Menopausal Patients

To evaluate the clinical utility of the AI model, we examined whether risk classification based on the AI could predict chemotherapy benefit. For this analysis, ideally, one should compare the outcomes of patients randomized to receive chemotherapy versus not in each risk group. However, in the design of the TAILORx trial, only patients with 11≤RS<26 were randomized to receive chemotherapy plus endocrine therapy (Arm C) or endocrine therapy alone (Arm B), which created a methodological challenge for this analysis (**Figure 1a**). We thus examined if we could draw conclusions from the data despite this challenge.

In our analysis of the TAILORx validation set, 12% of the patients were classified as high-risk by our model (AI≥26). Among these patients, 71.6% had RS≥26, 27.0% had 16≤RS<26, and only 1.4% had RS<16 (**Figure 3a**). The prospective TAILORx study showed that node-negative premenopausal women or those aged 50 or under with 16≤RS<26 derived some benefit from chemotherapy, and other prospective-retrospective studies, including the NSABP B-20 and SWOG-8814 cohorts, showed that women with RS≥26 derived even greater benefit^6,10,11,16^. Since 98.6% of the TAILORx patients classified by our AI model as high-risk had RS≥16, it would be unexpected if the remaining 1.4% had a large effect on the overall benefit estimates that would change this conclusion. It is thus very likely that chemotherapy significantly improves overall outcomes for premenopausal patients with AI≥26.

**Figure 3:**
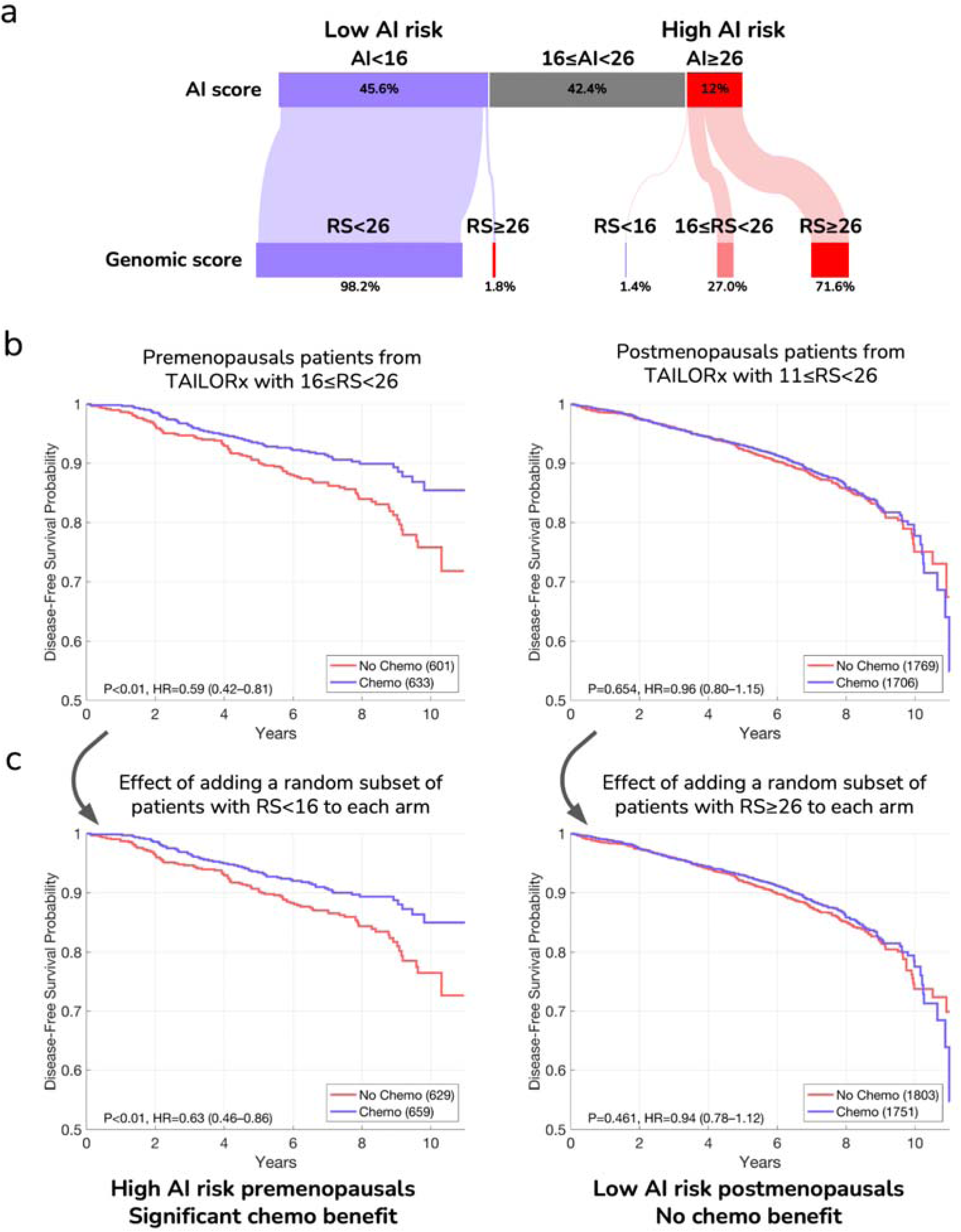
AI prediction of chemotherapy benefit by menopausal status. **(a)** Sankey diagram showing the relationship between AI score categories and genomic recurrence score (RS) categories on the TAILORx-validation set. The width of the flows represents the proportion of patients in each category. 1.4% of the high AI risk patients were RS<16, and 1.8% of the low AI risk patients were RS≥26. **(b)** Survival curves showing disease-free survival probability for patients receiving endocrine therapy alone (red) versus endocrine therapy plus chemotherapy (blue) for patients in TAILORx. Left: Chemotherapy benefit is observed for premenopausal patients with 16≤RS<26. Right: no chemotherapy benefit is observed for postmenopausal patients with 11≤RS<26. **(c)** Left: the same premenopausal patients with 16≤RS<26 from (b), with additional r sampled patients with RS<16 assigned to the two curves, showing no substantial effect on the ndomly overall chemotherapy benefit. The number of sampled patients patients was set to match 1.4% of the total patients in the analysis. Right: the same postmenopausal patients with 11≤RS<26 from (b), with additional r ndomly sampled patients with RS≥26 assigned to the two curves after correcting for the potential benefit of chemotherapy (Methods), showing no substantial effect on the overall chemotherapy benefit. The number of sampled patients patients was set to match 1.8% of the total patients in the analysis. Hazard ratios (HR) with 95% confidence intervals and P-values are shown for all comparisons.

Among patients classified as low-risk by our model (AI<16, 45.6% of all patients), 98.2% had RS<26, and only 1.8% had RS≥26. It was previously shown that node-negative postmenopausal patients with RS<26 derive no benefit from chemotherapy^13^. It would thus be highly unexpected that the 1.8% of patients with AI<16 but RS≥26 would have a substantial effect on the overall benefit estimates and change the conclusion. Thus, chemotherapy is not expected to improve overall outcomes for postmenopausal patients with AI<16.

To evaluate if these expectations hold, we estimated the effect of these small groups of patients (1.4% with RS<16 and 1.8% with RS≥26) on chemotherapy benefit within the remaining patients. As a reference point, we first evaluated chemotherapy benefit in pre- and post-menopausal patients in TAILORx using arms B and C for disease-free survival **(Figure 3b)**, distant recurrence-free interval, and recurrence-free interval **(Supplementary Figure 5a**), i.e., without the effect of the small groups.

For premenopausal patients, to estimate the effect of the 1.4% patients with AI≥26 but RS<16 on the overall chemotherapy benefit estimates, we randomly sampled a small subset of patients with RS<16 and assigned them to arms B and C (Methods). Indeed, with the addition of patients with RS<16, chemotherapy benefit remained significant across all clinical endpoints: disease-free survival (HR=0.63, 95% CI: 0.46–0.86, **Figure 3c**), distant recurrence-free interval (HR=0.55, 95% CI: 0.33–0.91), and recurrence-free interval (HR=0.55, 95% CI: 0.36– 0.82) **(Supplementary Figure 5b)**. For postmenopausal patients, to estimate the effect of 1.8% patients with AI<16 but RS≥26 on the overall chemotherapy benefit estimates, we randomly sampled a small subset of patients with RS≥26 and assigned them to arms B and C, while correcting for the potential benefit of chemotherapy (Methods). Indeed, with the addition of patients with RS≥26, chemotherapy benefit remained not significant across all clinical endpoints, with hazard ratios consistently near 1.0 (**Figure 3c and Supplementary Figure 5b**). Importantly, one-sided type I error of 10% corresponded to a 20% higher risk for disease-free survival when not receiving chemotherapy, which is well below the 32.2% threshold established in TAILORx for determining non-inferiority.

It should be noted that this analysis employed conservative methods that likely understate the true effect sizes. For premenopausal high AI risk patients, we primarily analyzed patients with 16≤RS<26, though most patients classified as AI≥26 had RS≥26 with potentially greater chemotherapy benefit. Additionally, we included a higher proportion of RS<16 patients than would be present in the actual AI≥26 group (Methods). For postmenopausal low AI risk patients, we excluded those with RS<11 who would likely further reduce any chemotherapy benefit, and the small fraction with AI<16 but RS≥26 predominantly had scores below 31, where chemotherapy benefit remains uncertain^51^. These conservative analytical choices strengthen confidence in our conclusions.

### External Validation on Real-world Data Demonstrates Robust Model Generalization

For external validation, we assembled data from six independent cohorts totaling 15,817 slides from 7,539 patients (**Figure 1b** and **Supplementary Table 1**), of which 7,502 slides from 5,497 patients met our HR+/HER2- inclusion criteria and slide quality standards (**Supplementary Figure 1**). Four cohorts had matched RS data: Carmel Medical Center (N=565 patients), Haemek Hospital (N=156 patients), Sheba Medical Center (N=427 patients), and the University of Chicago Medical Center (UCMC; N=490 patients). The Australian Breast Cancer Tissue Bank cohort (ABCTB; N=1,762 patients) provided extensive clinical information and long-term survival data without RS measurements. The Cancer Genome Atlas dataset (TCGA; N=594 patients) included RNA sequencing data that enabled the estimation of RS values. Notably, while TAILORx had only node-negative patients, the external cohorts had both node-negative and node-positive patients.

To overcome dataset variations and facilitate robust deployment of the AI model across external cohorts, we applied a distribution-matching calibration approach. The calibration does not require local genomic testing data, which is especially important in regions lacking access to routine genomic testing. The method uses a simple linear scaling transformation that requires an estimate of the mean RS in the new cohort and can be computed from clinical features alone (Methods, **Supplementary Figure 6**). To calibrate the model to each external cohort, we used 100 patients from this cohort. The 100 calibration patients were excluded from the subsequent analyses unless stated otherwise. We chose 100 samples to retain sufficient data for downstream experiments, and because our analysis showed stable performance at this calibration size with diminishing returns beyond it (**Supplementary Figure 7**).

We applied the calibrated model to Carmel, Haemek, Sheba, and UCMC, and evaluated its performance. Our analysis demonstrated high correlations between the RS and AI scores across all validation cohorts, with Pearson correlations ranging from 0.688 to 0.754 (**Supplementary Figure 8)**. The AUC performance for identifying RS≥26 ranged from 0.858 to 0.903 **(Figure 4a-d, Supplementary Table 3)**. For comparison, we further applied an image model using only H&E images, which resulted in an AUC performance range of 0.837–0.884, showing that, as in the TAILORx data, the AI multimodal model outperformed the image-only model across all external datasets.

**Figure 4:**
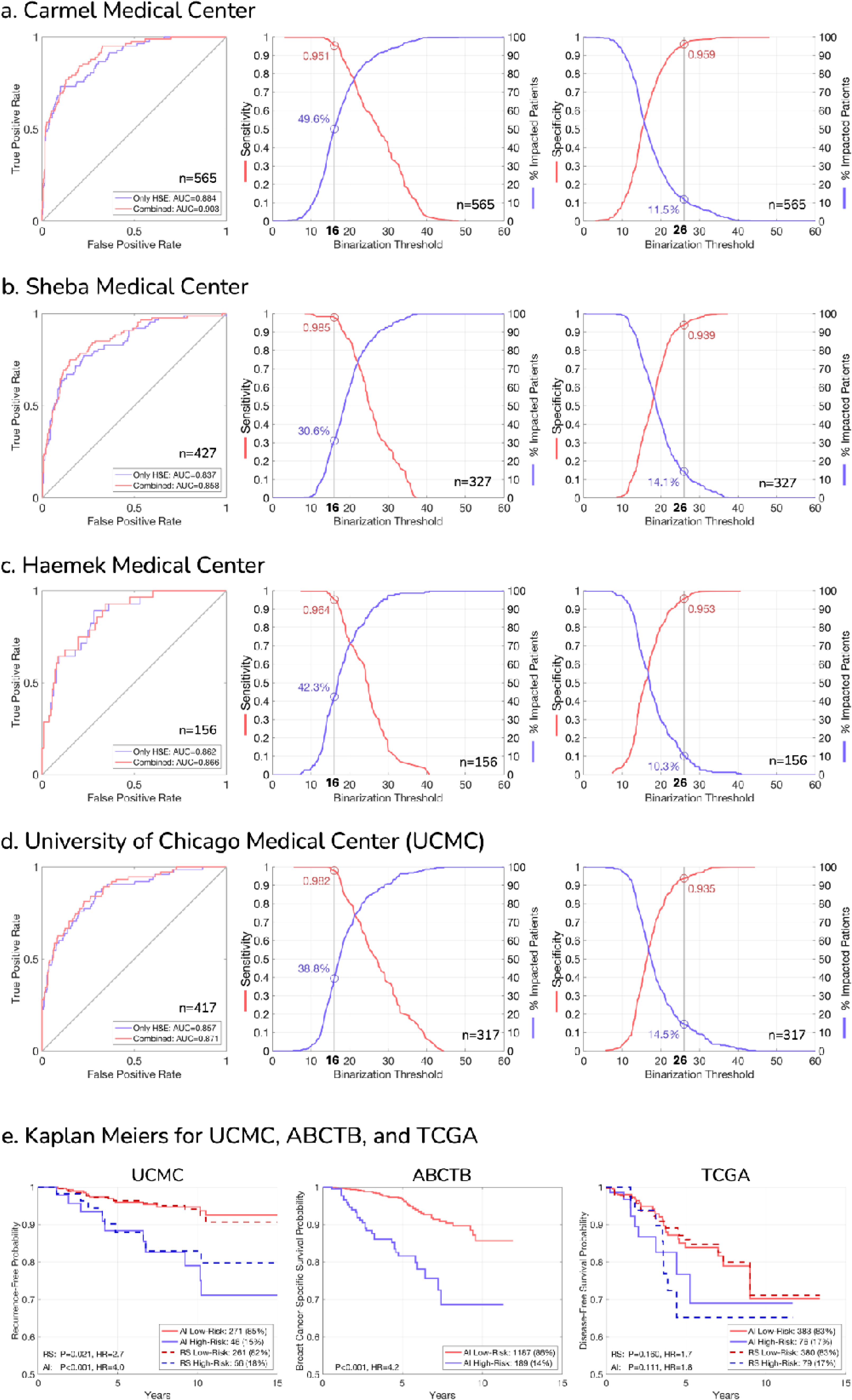
external validation of the deep learning AI model across independent cohorts and pr gnostic performance. **(a-d)** AI model performance at Carmel Medical Center, Sheba Medical Center, Haemek Medical Center, and the University of Chicago Medical Center (UCMC), each with matched recurrence scores (RS). For each institution: **Left:** Receiver operating characteristic (ROC) curves comparing the AI multimodal model using both H&E images and clinicopathologic variables (“combined”) with the image model (only H&E) for identifying high genomic risk disease (RS≥26). **Middle:** Sensitivity analysis showing the proportion of patients classified as low AI risk, which could potentially avoid genomic testing (blue line), and the sensitivity for identifying RS≥26 (red line) at different classification thresholds. Values corresponding to the low classification threshold 16 are indicated. **Right:** Specificity analysis showing the proportion of patients classified as high AI risk, which could potentially proceed directly to chemotherapy (blue line), and the specificity for identifying RS≥26 (red line) at different classification thresholds. Values corresponding to the high classification threshold 26 are indicated. Note: For sensitivity/specificity analyses at UCMC and Sheba, the calibration set patients (n=100) were excluded. These patients were included in ROC analyses as calibration does not affect ranking-based metrics. **(e)** Prognostic performance of RS and AI scores across external cohorts with follow-up data. Kaplan-Meier curves show patient stratification using the high-risk threshold (AI≥26) for UCMC (recurrence-free interval), ABCTB (breast cancer-specific survival), and TCGA (disease-free interval). For UCMC and TCGA, RS-based stratification (RS≥26) is shown for direct comparison. For TCGA, RS values were estimated from the gene expression data. Hazard ratios (HR) with 95% confidence intervals and P-values demonstrate the consistent prognostic value of AI-derived risk stratification across diverse patient populations.

We then applied the low (16) and high (26) classification thresholds for clinical utility **(Figure 4a-d, Supplementary Figure 9, Supplementary Table 3).** Across the external cohorts, the model classified 30%– 50% of the patients as low-risk (AI<16) with the following performance: Carmel (sensitivity=0.951, NPV=0.986), Sheba (sensitivity=0.985, NPV=0.990), Haemek (sensitivity=0.964, NPV=0.985), UCMC (sensitivity=0.982, NPV=0.992). Similarly, the model classified 10%–15% of the patients as high-risk (AI≥26) with the following performance: Carmel (specificity=0.959, PPV=0.692), Sheba (specificity=0.939, PPV=0.652), Haemek (specificity=0.953, PPV=0.625), and UCMC (specificity=0.935, PPV=0.630), potentially supporting immediate initiation of chemotherapy for these subsets of patients. Notably, the performances and proportions of low and high AI risk patients remained consistent across the cohorts, despite the dataset variations, and despite having node-positive patients, which were not included in the TAILORx data. Comparing the AI model’s performance before and after calibration demonstrated improved consistency across cohorts, highlighting the importance of our calibration approach (**Supplementary Figures 10-11**).

For additional validation, we analyzed the TCGA cohort. As direct Oncotype DX results were not available, we estimated RS values from the available RNA sequencing data (**Methods**). Despite using these computed RS values and working with highly heterogeneous patient data and specimens, the AI model maintained good discrimination (AUC=0.832) and preserved consistent operating points, classifying 44.7% of patients as low AI risk (sensitivity=0.920, NPV=0.968) and 16.7% as high AI risk (specificity=0.899, PPV=0.500; **Supplementary Figures 8, 9, 12**).

Overall, when applied to external cohorts, the AI model showed consistent performance between the cohorts, and showed strong generalization and maintained performance, despite data variation and the addition of node-positive patients.

To evaluate the generalizability of our AI multimodal model for prognostication, we conducted survival analyses across three external cohorts with follow-up data: UCMC, ABCTB, and TCGA. The UCMC cohort, which included both matched genomic testing results and outcome data, enabled a direct comparison of our model’s prognostic value to that of the genomic test (**Figure 4e**). When stratifying patients using RS≥26, we observed significant risk separation for UCMC (HR=2.7, p-value=0.021). Stratification using our AI scores demonstrated comparable hazard ratios (HR=4.0, p-value<0.001), identifying a similar proportion of high-risk patients (15% vs 18%). In the ABCTB cohort, which lacked genomic testing data but provided long-term follow-up, our model also demonstrated significant stratification of breast cancer-specific survival (HR=4.2, p-value<0.001). As in the TAILORx cohort, the AI model’s prognostic performance yielded hazard ratios comparable to those of RS across various clinical subgroups in both UCMC and ABCTB cohorts (**Figure 2d**). For the TCGA cohort, where RS values were computationally estimated from RNA sequencing data, our model retained prognostic value (HR=1.8, p-value=0.111) for disease-free interval, although the result was not statistically significant. However, we also observed that stratification based on the estimated RS similarly resulted in non-significant risk separation (HR=1.7, p-value=0.160). This result may reflect the heterogeneity of the TCGA dataset and the use of outdated treatment protocols.

It is important to note that these survival analyses rely on observational data, in which treatment acts as a confounding variable. As such, this data is suboptimal for evaluating the true prognostic power of the AI model or for direct comparison to RS. Nonetheless, the robust stratification of survival outcomes across these diverse cohorts provides compelling evidence that our model captures clinically relevant risk factors and generalizes well across external datasets.

### The AI Model Reclassifies Clinical Risk in a Substantial Subset of Patients

We next examined the potential clinical utility of our model in settings where genomic testing has limited accessibility. In such scenarios, treatment decisions typically rely on conventional clinicopathologic variables, such as tumor size, histological grade, lymph node status, and receptor expression. While several clinical risk classification methods are in routine use^52,53^, these tools do not capture the unique biological characteristics of each tumor. Consequently, many breast cancer patients are likely overtreated and unnecessarily exposed to the toxicities of adjuvant therapy without substantial benefit. We thus assessed the potential impact of our model on treatment decisions based on clinical risk. Specifically, we evaluated clinical risk as defined in the MINDACT trial^7^, and compared those assessments with the AI risk.

Comparing the AI risk to traditional clinical risk assessments revealed substantial reclassification. For node-negative patients, in both the TAILORx dataset and external cohorts, 4.5–4.9% of clinically low-risk premenopausal patients were reclassified as high AI risk by our model. More substantially, 31.3–32.7% of clinically high-risk postmenopausal patients were reclassified as low AI risk. For node-positive postmenopausal patients, our model downgraded 40.7% of the clinically high-risk patients (**Figure 5**).

**Figure 5:**
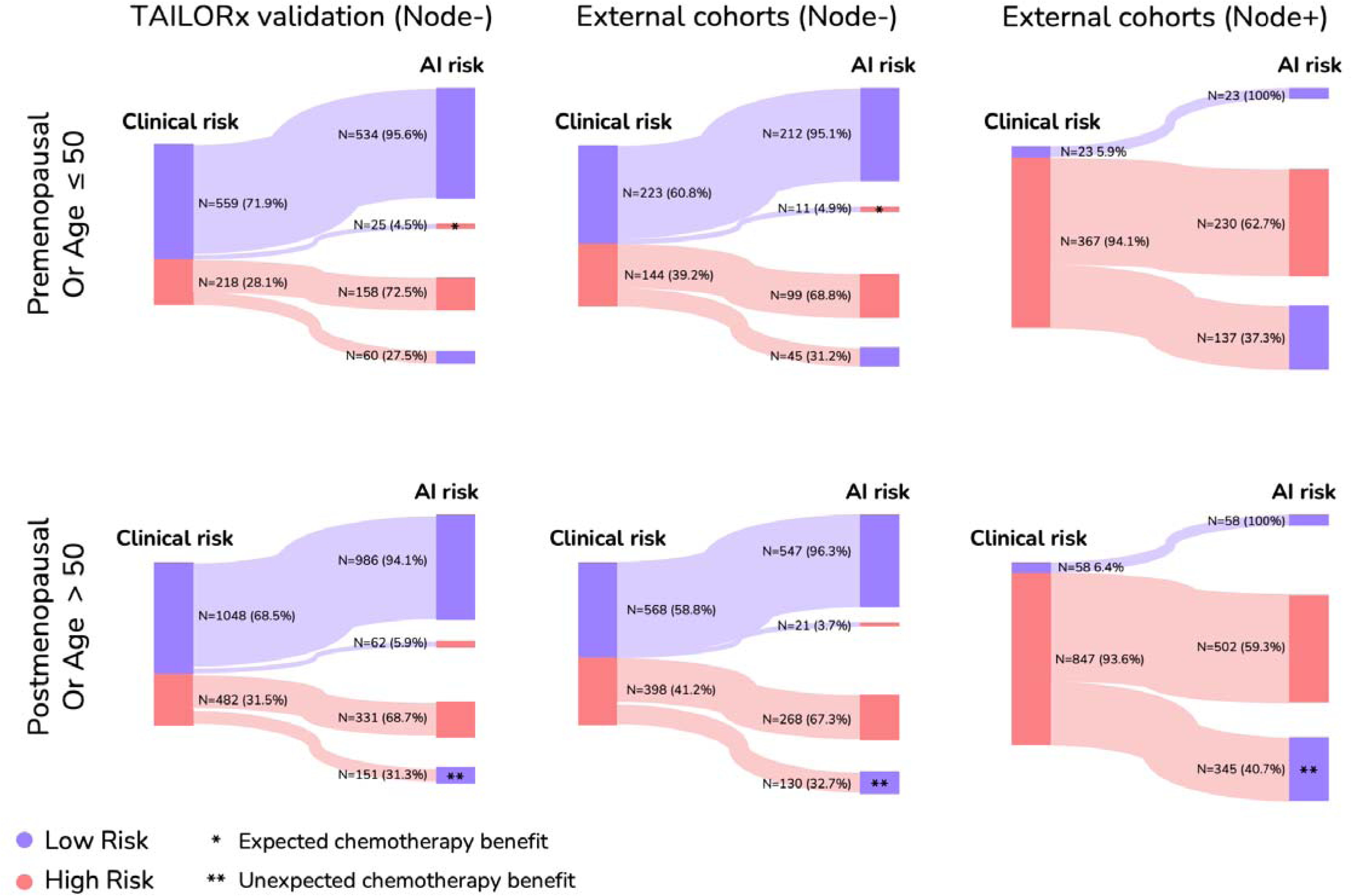
Clinical risk reclassification. Sankey diagrams showing how patients classified as high or low risk by traditional clinical risk assessment (MINDACT definition) would be reclassified using the AI model across three patient cohorts: TAILORx validation set (left), node-negative patients in the external cohorts (middle), and node-positive patients in the external cohorts (right). Diagrams are stratified by menopausal status for TAILORx and by Age for the external cohorts (premenopausal/age ≤50, top; postmenopausal/age >50, bottom). The width of connecting flows indicates the proportion of patients, with exact numbers and percentages shown. Blue represents low-risk classification, red represents high-risk classification. Asterisks indicate a potential impact on chemotherapy decisions based on clinical risk, following our survival analyses (Figure 3) and observations (*Expected chemotherapy benefit, **Unexpected chemotherapy benefit).

Our earlier analysis (**Figure 3**) provided strong evidence that node-negative postmenopausal patients classified as low AI risk by our model did not benefit from chemotherapy. For node-positive patients, however, we could not directly evaluate chemotherapy benefit using TAILORx, which included only node-negative disease. Nonetheless, important insights can still be drawn. For node-positive premenopausal patients, current guidelines recommend chemotherapy for all, irrespective of genomic risk, based on findings from the RxPONDER trial^15^. In contrast, node-positive postmenopausal patients with RS<26 showed no chemotherapy benefit (disease-free survival HR = 1.02, 95% CI: 0.82–1.26). Among patients classified as low-risk by our AI model in this group, only 3.4% had RS≥26. Furthermore, the vast majority had RS<31 (NPV=0.994 for RS<31, meaning that only 0.6% of the patients classified as high AI risk by our model had RS≥31), a range in which chemotherapy benefit remains uncertain^51^. Thus, in line with our previous analysis (**Figure 3**), it is highly unlikely that this small fraction of high-RS cases would impact the hazard ratios in RxPONDER such that significant chemotherapy benefit would be observed.

These findings demonstrate that our AI model has the potential to impact treatment decisions based on clinical risk, especially in postmenopausal patients, and to expand access to precision oncology in settings where genomic testing is inaccessible.

### Interpretable Model Scores

To understand the biological basis of the AI scores and gain insights into how the model makes decisions, we visualized risk scores across whole-slide images. We applied our model to local regions (10×10 tiles) of the H&E image, treating each region as a bag of tiles within the multiple-instance learning framework. This approach generated both local scores for each region and gradient-based attention scores indicating which areas most influenced the model’s decisions (**Figure 6**).

**Figure 6:**
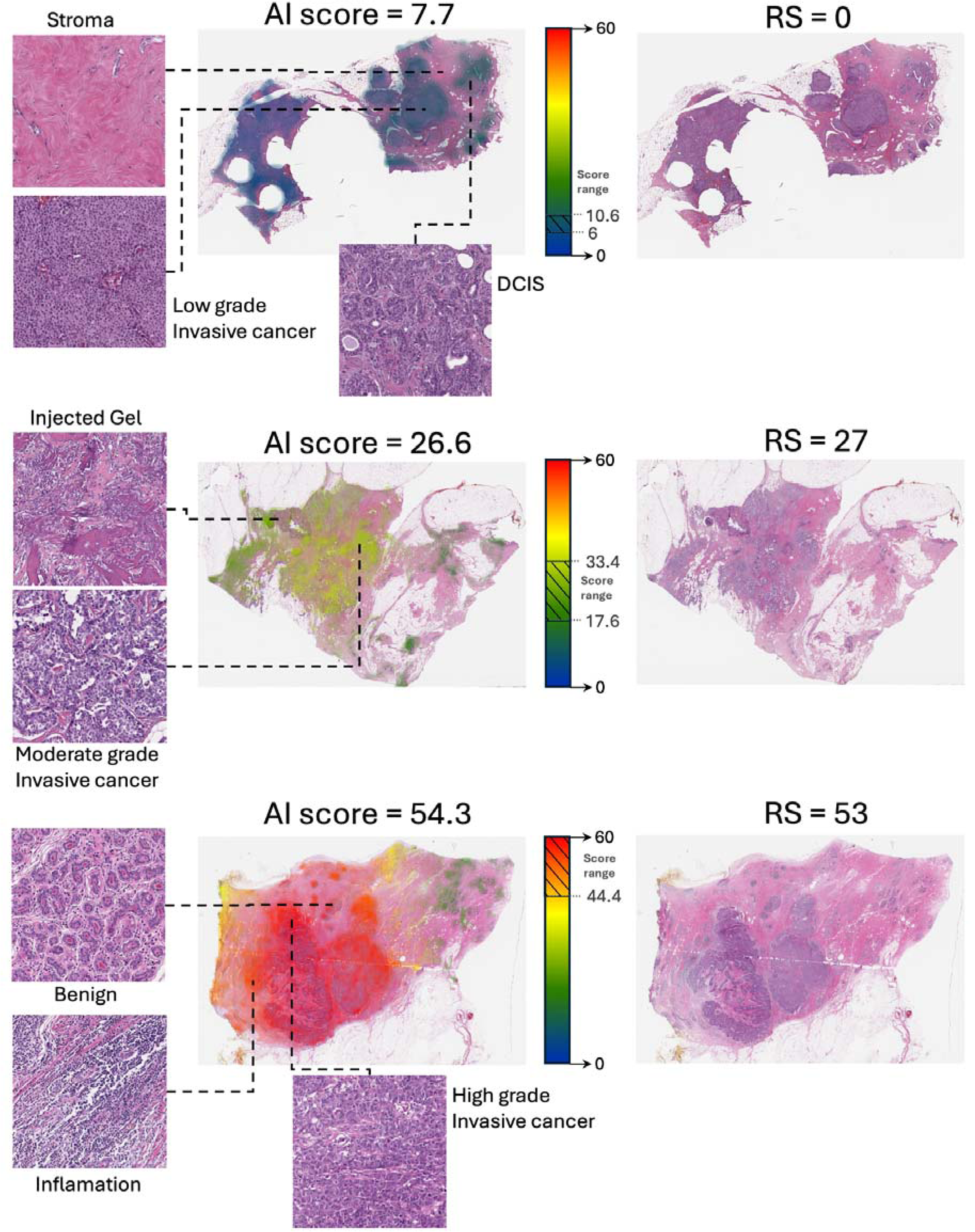
Visualization of AI scores at tissue-level granularity reveals biological patterns. Hematoxylin & eosin (H&E) whole-slide images are shown both in their original form and with AI scores heatmaps for three representative cases spanning the risk spectrum. Left panels show magnified regions highlighting key morphological features, including stromal tissue, invasive cancer of varying grades, ductal carcinoma in situ (DCIS), benign tissue, and regions of inflammation. Center panels display heatmaps overlaid on the H&E images, where color intensity indicates the model’s attention and hue represents local AI scores (scale shown in color bars). The right panels show the original H&E images.

Visualization results revealed that the model primarily focused on tumoral regions, including areas of in situ carcinoma, though with less intensity than regions of invasive cancer. In contrast, stromal tissue, benign regions, and non-tissue elements (e.g., injected gel) had minimal influence, suggesting that the model draws on clinically relevant morphological features. Importantly, localized AI scores closely matched the whole-slide risk assessments. Even small regions of tumor tissue produced consistent results, indicating that prognostic signals are detectable at a fine-grained level. This implies the model identifies features within individual tumor cells or small clusters that are prognostic for recurrence risk, rather than relying on large-scale architectural patterns. Consequently, even limited tumor sampling may suffice for accurate risk estimation.

Visual inspection of cases with high versus low AI scores revealed clear morphological distinctions. High AI scores were associated with increased peritumoral inflammation, high histological and nuclear grades (including larger, more variable nuclei), and elevated mitotic activity. In contrast, low AI scores corresponded to minimal inflammation, lower histological and nuclear grades, and reduced mitotic activity. These findings align with known correlations between tumor grade and recurrence risk. However, the model occasionally identified low RS in high-grade tumors and high RS in low-grade cases, suggesting that it detects additional, more subtle morphological features that go beyond standard histopathological grading and may not be readily apparent on manual review.

## Discussion

In this study, we developed a multimodal deep learning model aimed at estimating Oncotype DX recurrence scores from H&E-stained histopathology images and clinical variables. Our model was tuned and validated on the TAILORx clinical trial dataset, offering controlled treatment assignment that enables estimation of chemotherapy benefit while minimizing confounding. By leveraging a foundation model pre-trained on 171,189 histopathological slides and implementing a distribution-matching calibration method, we achieved robust generalization across six independent cohorts comprising over 5,000 patients. Despite variations in patient populations, laboratory protocols, and scanning equipment, our model maintained consistent performance and operating characteristics, classifying 30-50% of patients as low AI risk with >94% sensitivity and 10-17% as high AI risk with >95% specificity. These innovations significantly reduce implementation barriers, as centers in resource-limited settings can deploy our model with just 100 slides for calibration with no need for genomic labels.

Of particular clinical importance, our findings show that our model’s risk classification is correlated with chemotherapy benefit in premenopausal and postmenopausal patients. For node-negative postmenopausal patients classified as low-risk by our model (AI<16), 98.2% had Oncotype DX RS<26, aligning with the TAILORx finding that such patients derive no benefit from chemotherapy. Among node-negative premenopausal patients classified as high-risk by our model (AI≥26), 98.6% had RS≥16, aligning with the TAILORx finding of chemotherapy benefit in this group. Our analysis estimated the effect of the small percentage of discordant classifications and demonstrated that these are unlikely to alter the expected treatment benefits. While we did not directly test for chemotherapy benefit based on AI predictions alone, the strong concordance with genomic risk categories and the consistency of these observations across multiple clinical endpoints suggest that our model could effectively guide treatment decisions in these specific clinical contexts.

Our model demonstrated generalizability to node-positive patients on the external cohorts, despite being tuned exclusively on node-negative cases from TAILORx. This cross-domain generalization suggests that our model captured biological signals reflective of the genomic score, regardless of the nodal status. While we could not directly assess chemotherapy benefit in node-positive patients using TAILORx data, the RxPONDER trial^15^ results provide external context - all premenopausal node-positive patients are recommended chemotherapy regardless of RS, while postmenopausal node-positive patients with RS<26 showed no chemotherapy benefit. Other observational studies^51^ showed an uncertain chemotherapy benefit for postmenopausal patients with 26≤RS<31. For node-positive postmenopausal patients classified as low-risk by our model (AI<16), 96.6% had RS<26, and notably, only 0.6% had RS≥31, suggesting that, as for node-negative patients, node-positive postmenopausal patients classified as low AI risk are not expected to derive benefit from chemotherapy.

These implications are particularly relevant for low-income countries, where genomic testing remains inaccessible or prohibitively expensive. In many regions, such as parts of Asia, Africa, and South America, clinical decisions rely primarily on traditional histopathological assessments, which lack the predictive accuracy of genomic assays. For node-negative premenopausal patients, our model identified approximately 4-5% of the clinically low-risk patients who may benefit from chemotherapy. More substantially, our model downgraded 30-40% of clinically high-risk postmenopausal patients, suggesting that these patients are likely overtreated and unnecessarily exposed to the toxicities of adjuvant therapy without substantial benefit. These results indicate that H&E-based deep learning could serve as a viable alternative, enabling precise risk stratification without the need for expensive molecular testing. Even in high-income countries, reducing the need for genomic testing in a substantial proportion of patients could lower healthcare costs and streamline treatment initiation. Beyond cost reduction, there are additional advantages. Results are available within minutes rather than weeks, potentially accelerating treatment decisions. Additionally, our method requires only routine H&E slides, avoiding the need for additional tissue sampling that genomic tests require. This is particularly important given concerns about tissue utilization and preservation, especially in smaller tumors or resource-constrained settings.

Our findings build upon and extend several important prior works in this field. Previous studies have demonstrated the feasibility of identifying high Oncotype DX recurrence scores from histopathology images using various computational approaches^37–43,54–58^. While these studies have laid important groundwork, our approach distinguishes itself in several key aspects. First, our validation on the TAILORx dataset enabled the evaluation of our model on one of the largest randomized control trials in breast cancer, specifically designed for Oncotype DX RS. Crucially, TAILORx enabled addressing treatment confounders and assessing chemotherapy benefit in patient subgroups. Second, our extensive external validation across six diverse cohorts comprising over 5,000 patients exceeds the validation scope of prior work, establishing robust generalization across different clinical settings and addressing calibration issues. Third, our foundation model approach, pre-trained on 171,189 histopathological slides, demonstrates significantly superior performance compared to the state-of-the-art fully supervised learning methods across all validation cohorts (AUC of 0.858-0.903 versus 0.774-0.861, **Supplementary Table 4**), and particularly in external validation where domain shifts typically challenge generalization. For example, in direct comparison on the UCMC dataset for identifying high-risk (RS≥26), our model achieved AUC=0.871, substantially outperforming previously published model^37^ (AUC=0.823) on the same dataset.

Other studies aimed to infer patient outcomes directly from H&E images, including regulatory-approved commercial tools like PreciseDX, Stratipath, Ataraxis, and RlapsRisk^33,35,59–64^. Although this approach may hold important advantages, such as outperforming existing genomic tests and expanding to broader populations, its clinical impact remains uncertain without validation using randomized controlled trials, where treatment effects are accounted for. Trials having all patients randomized to chemotherapy are scarce and have limited availability. In modern datasets, nearly all high-RS patients receive chemotherapy, making it impossible to observe the outcomes of untreated high-risk disease and, consequently, to evaluate the ability of these tools to predict treatment benefit. Instead, training the models to directly infer RS bypasses these challenges by building upon its already established clinical utility using previous clinical trials.

Several important limitations should be considered when interpreting our results. First, our analysis of chemotherapy benefit is constrained by the TAILORx trial design, which only randomized patients with intermediate RS (11-25), limiting our ability to directly assess treatment effects in patients with RS≥26. Future validation using data from trials like NSABP B-20, which included randomization of high-risk patients to chemotherapy versus no chemotherapy, would provide additional evidence for the model’s predictive ability in this population and may enable expansion of the model’s low and high-risk groups. Second, while our model demonstrated encouraging performance on node-positive patients in external validation cohorts, validation on randomized data like the RxPONDER trial would strengthen our conclusions for node-positive patients. Third, the development and validation were conducted retrospectively, and prospective implementation studies would provide stronger evidence of the model’s ability to guide chemotherapy decisions in real-world clinical practice. Fourth, high-quality testing of ER, PR, and HER2 is required for the identification of eligible HR+/HER2- patients, which may be lacking in some low and middle income countries. Future efforts may require the development and integration of methods for accurate molecular subtype classification. Finally, while our patient populations span diverse geographic regions, certain populations remain underrepresented, particularly from low-income countries where the need for cost-effective risk stratification may be greatest. Future work should address these gaps through targeted validation studies in these specific populations and clinical contexts.

In conclusion, deep learning applied to routine H&E slides provides rapid, cost-effective risk stratification with prognostic accuracy comparable to genomic assays. Our approach is extensively validated across diverse populations, requires no specialized genomic infrastructure, and has the potential to democratize precision oncology in resource-limited settings. By eliminating the need for genomic testing in low-risk patients and expediting chemotherapy for high-risk patients, this method could significantly reduce healthcare costs while maintaining high standards of care. Future work should focus on prospective validation to further establish this approach as an accessible, efficient, and globally scalable tool for breast cancer treatment decisions.

## Methods

### Patient Cohorts

Our data collection consists of 25,436 H&E slides from 17,812 patients across seven independent cohorts, of which 16,885 H&E slides from 13,781 patients were used for the analysis (**Supplementary Tables 1-2**). The variety of scanners, magnifications, and staining reagents offers a robust framework for assessing the system’s generalizability and accuracy in diverse clinical settings. The TAILORx cohort served for tuning and validating our AI models. This data was unique in several aspects: it is one of the largest clinical trials ever done in breast cancer, it included randomization for chemotherapy plus endocrine therapy versus endocrine therapy alone, and it was designed to evaluate the Oncotype DX assay. These characteristics enabled a robust validation of our models and a direct comparison of our model against Oncotype DX on unbiased data and using multiple clinical endpoints. Our external validation cohorts consisted of Carmel (n=565), Haemek (n=156), and Sheba (n=427) from Israel, UCMC (n=490) from Chicago, USA, ABCTB (n=1,762) from multiple sites in Australia, and TCGA-BRCA (n=594) from multiple sites across the USA. Additionally, the Carmel-Calibration (n=1,176) and Haemek-Calibration (n=327) datasets were collected for calibrating the AI models to the Carmel and Haemek cohorts and for additional calibration experiments.

As the TCGA-BRCA cohort does not have the Oncotype DX scores of the patients, we estimated the scores using transcripts per million (TPM) expression data. Formulas from the published development of Oncotype DX were then applied to the mRNA expression data to estimate the RS^65^.

Inclusion and exclusion criteria (**Supplementary Figure 1**): we included patients with HR+/HER2- invasive breast cancer. For TAILORx, Carmel, Haemek, Sheba, and UCMC, only patients who had Oncotype DX scores were included. For Carmel-Calibration, Haemek-Calibration, TCGA, and ABCTB, the analysis did not require Oncotype DX scores. Exclusions were limited to slides that had physical scan issues, resulting in a blurred or corrupted image. To have at least 100 tiles in a bag in our MIL configuration, we also excluded slides that contained fewer than 100 tissue tiles, representing 1.63 square millimeters of tissue. Such slides were rare, constituting only 28 (0.1%) slides in all cohorts. Other than these exclusions, no data curation was done.

All H&E slides were obtained from formalin-fixed paraffin-embedded tissues, except for TCGA, which also included slides prepared from frozen tissues. The diversity in data collection, spanning multiple geographic locations, time periods, and preparation protocols, ensures that our cohorts are representative of real-world clinical settings and provides a comprehensive framework for evaluating our AI model’s performance and generalizability.

### Slide segmentation and tiling

Digitized WSIs were segmented into tissue and background regions utilizing Otsu’s method^66^, enabling the exclusion of background areas. The segmented slides were then divided into non-overlapping tiles, each measuring 256×256 pixels. Tiles were extracted at a magnification of micron per pixel (MPP)=0.5.

### Model architecture, hyperparameters, and optimization

The methodology consists of three parts - feature extraction foundation model, downstream finetuning, and aggregation of clinical features.

#### Feature extraction foundation model

Prov-GigaPath foundation model^45^ was trained using self-supervised learning, which enables training the model using a vast amount of data with no specific task. The model was trained on a pan-cancer dataset of 1.3 billion 256×256 pathology image tiles from 171,189 H&E WSIs originating from over 30,000 patients covering 31 major tissue types. The foundation model employed a novel vision transformer architecture designed specifically for pretraining on gigapixel pathology slides. The tile encoder was pre-trained using DINOv2, which is the state-of-the-art image self-supervised learning framework^48^. To effectively model whole slides containing tens of thousands of image tiles (as many as 70,121 in the Providence data), Prov-GigaPath adapts the LongNet method, which utilizes dilated self-attention to efficiently capture both local patterns in individual tiles and global patterns across the entire slide.

#### Downstream task

We extracted feature vectors from all foreground 256×256 tile images at MPP=0.5 using the pre-trained foundation model. We then finetuned a transformer-based multiple-instance-learning architecture to predict the slide-level Oncotype DX scores from the tile features. This model is trained to predict the slide-level RS from bags of tile feature vectors. For efficient and effective aggregation of the ultra-large collection of image tiles, the downstream model employs a slide encoder based on the LongNet architecture^67^, which enables ultra-long-context modeling that can handle sequences of tens of thousands of tile embeddings. We used a mean squared error (MSE) loss, an AdamW optimizer with an initial learning rate of 2.5e-4, a weight decay of 0.05, a minibatch size of 32, and a cosine learning rate scheduler. The number of epochs was set to 3. In the case of multiple slides per patient, we averaged their predictions to obtain patient-level scores.

#### Integration of clinical features

To incorporate clinical features within the tuning process, we obtained slide scores for the tuning set using the trained feature extraction and downstream models. We then trained a classifier using linear regression with no regularization to predict the Oncotype DX score from both the image-based slide scores and the clinical features. Clinical features included histological grade, hormone receptor status (PR, ER), patient age, tumor size (dichotomized at 2cm), surgery type (mastectomy/lumpectomy), self-reported race (White, Black, Asian, Native Hawaiian or Pacific, Islander, or Native American), and menopausal status.

The multimodal model (referred to in the paper as ‘the model’ or ‘the AI model’), which predicts the RS from H&E images and clinical variables, consists of all three components, i.e., the feature extraction foundation model, the downstream model, and the linear regression. To predict the RS from the clinical features alone, without the H&E images, we used the same linear regression approach, only without adding the H&E scores as a feature. The total inference time of the multimodal model averages approximately 3 minutes per slide, with the feature extraction part consuming 99.96% of the overall time.

### Tuning and Inference

The TAILORx data was randomly partitioned into tuning (70%) and validation sets (30%). The tuning set was further randomly subdivided into five folds for 5-fold cross-validation. All partitions were made at the patient level to ensure no overlapping patients between the folds and the validation set. The number of epochs, which was fixed to 3, was the only hyperparameter tuned during the cross-validation process. Post-tuning, an ensemble approach was adopted, averaging the prediction scores of the five cross-validation models to enhance robustness and generalizability across both the TAILORx validation set and external cohorts, without any additional parameter tuning.

### Feature Contribution

To compute the feature importance, we iteratively added features to the model based on their contribution. We measured the contribution of a feature as the difference in the R-squared performance of the model on the validation set when incorporating or not the feature during training.

### Kaplan Meier Estimates for Chemotherapy Benefit

To estimate if chemotherapy benefit could be determined for premenopausal patients classified as high-risk (AI≥26), we remember that out of AI≥26 patients, 71.6% had RS≥26, 27.0% had 16≤RS<26, and only 1.4% had RS<16. As we did not have randomization for RS≥26, we focused on estimating chemotherapy benefit within RS<26, which would serve as a lower bound for the benefit in the entire group. Note that when constraining to RS<26, the portion of patients who had RS<16 rises from 1.4% to 4.9%. We also remember that in a post-analysis of the TAILORx, outcomes were compared between Arm B (endocrine therapy alone) and Arm C (chemoendocrine therapy) for premenopausal patients with 16≤RS<26, showing significant chemotherapy benefit^16^. To estimate if the addition of a small subset of patients from RS<16 would change this conclusion, we first repeated the TAILORx analysis as a reference point (**Figure 3b and Supplementary Figure 5a**). Then, we sampled patients with RS<16 and assigned them to these two groups, such that their relative proportion would match 4.9%. For this, patients sampled from 11≤RS<16 were assigned to the two treatment groups according to their original trial arm (B or C). Patients sampled from RS<11 (Arm A, received endocrine therapy alone) were randomly assigned to the two groups. Assigning patients who did not receive chemotherapy to both groups can only decrease the observed chemotherapy benefit, serving as a conservative lower bound for chemotherapy benefit. Following this construction, observing chemotherapy benefit for premenopausal patients within 16≤RS<26 and with additional patients with RS<16 should serve as a lower bound for determining chemotherapy benefit in premenopausal patients classified as high-risk (AI≥26).

This analysis relied on several key assumptions that warrant discussion. First, we estimated chemotherapy benefit by examining TAILORx patients with their proportions adjusted to match the AI risk distributions. This approach transformed our question from a conditional analysis (chemotherapy benefit given AI score) to an unconditional analysis (chemotherapy benefit in a population with a specific RS distribution). We argue this assumption is reasonable because there is no reason to believe that chemotherapy benefit in a population selected for high AI scores (AI≥26) would be lower than that of an unselected population with the same RS distribution; rather, it is expected to be higher. Second, our analysis is conservative in several ways: we primarily analyzed patients with RS<26, even though the majority of patients classified as AI≥26 had RS≥26, which is associated with greater chemotherapy benefit. Additionally, we included a higher proportion of RS<16 patients (4.9%) in our simulation than would actually be present in the AI≥26 group (1.4%), which would tend to underestimate the true benefit. These methodologically conservative choices strengthen confidence in our conclusions about chemotherapy benefit.

To estimate if chemotherapy benefit could be ruled out for premenopausal patients classified as low-risk (AI<16), we remember that only 1.8% of these patients had RS≥26, which is unlikely to significantly change the TAILORx conclusion, that postmenopausal patients with RS<26 do not benefit from chemotherapy. We estimated the potential effect of this small subset of patients on the conclusion. Since no randomization was done for RS<11, we focused on patients with RS≥11, which would serve as an upper bound for chemotherapy benefit in the entire group. Constraining to RS≥11, 2.1% of the patients had RS≥26. As before, we repeated the TAILORx analysis as a reference point for patients with 11≤RS<26, showing no observed chemotherapy (**Figure 3b and Supplementary Figure 5a**). Then, we sampled patients with RS≥26 (Arm D, all received chemoendocrine) and assigned them to these two groups, such that their relative proportion would match 2.1%.

However, assigning patients who received chemotherapy to the no-chemo group could falsely reduce the observed chemotherapy. Therefore, we applied a correction to account for the potential improvement in prognosis due to chemotherapy in these patients. Previous studies estimated the hazard ratio for chemotherapy benefit in patients with RS≥26^11,12^. HR for DRFI was estimated as 0.44, and HR for DFS was estimated as 0.45. HR for RFI was not reported but can be roughly estimated to be higher than that of DRFI from the Kaplan-Meier plots, thus, we used 0.44 as a lower bound. To apply the correction, we estimated the survival probability *S_Chemo_*(*t*) for patients with RS≥26 based on the outcomes in TAILORx and estimated their survival probability with no chemotherapy as

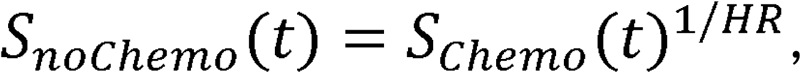

following the standard proportional hazards assumption. The patients assigned to the no-chemo group were then sampled from this corrected distribution. Following this construction, ruling out chemotherapy benefit within postmenopausal patients within 11≤RS<26, and with additional patients with RS≥26, should serve as a higher bound for chemotherapy benefit in postmenopausal patients classified as low AI risk. For the non-inferiority assessment, we used the same 32.2% margin established in TAILORx for DFS, with one-sided type I error of 10%.

As before, the conditional probability assumption was made. Additionally, we assumed that HR estimates from previous studies, which were based on the NSABP-20 trial, hold for the TAILORx population. However, we should stress that in fact, only 0.7% of the AI<16 patients had RS≥31, where chemotherapy benefit is more established, while benefit in RS<31 remains uncertain^51^. Proportional Hazards were assumed for the correction within Arm D in TAILORx. However, this common assumption was also made in the TAILORx trial analysis for computing the hazard ratios. Finally, we excluded those with RS<11 who would likely further reduce any chemotherapy benefit, and included a higher proportion of RS≥26 patients (2.1%) than would be present in the actual AI≥26 group (1.8%).

### Model Calibration

Although our model demonstrated excellent generalization on the external cohorts in terms of overall performance, we observed that the distributions of the AI scores often diverged from the RS distribution. Despite the true RS distribution remaining relatively consistent across datasets, the AI score distributions varied from one cohort to another, with a general tendency for higher AI values than the true RS, on average. This phenomenon is common when applying deep learning to external datasets and should be considered. To address these calibration issues, we aimed to develop a minimally parameterized *linear scaling* approach to align the AI scores with RS.

Importantly, Oncotype DX is not routinely performed in many regions, making direct RS-based calibration unfeasible. Hence, we sought a solution that does not require direct knowledge of RS. Subsequently, this strategy also enabled us to calibrate the model on some of our own datasets that lacked RS measurements (ABCTB, Carmel-calibration, Haemek-calibration, and TCGA). We adopted a *linear scaling* strategy by introducing a single scaling factor a, such that:

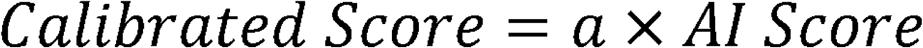

where a is computed as the ratio of the mean RS to the mean of the AI scores:

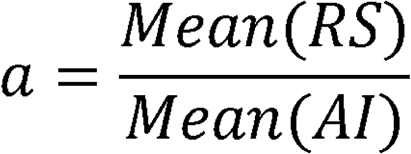

This approach enabled reliance only on an estimate of the mean RS in each new cohort. In pilot comparisons, more flexible affine transformations (including both scale and shift) yielded comparable results, supporting the sufficiency of a single-parameter calibration, which is preferred for robustness.

We further showed that the mean RS could be estimated from basic clinicopathologic parameters **(Supplementary Figure 6)**. Subsequently, we used the clinicopathologic-based model (trained using only clinicopathologic variables) to predict an approximate RS for patients in each external cohort and then took the average of these predictions as our estimated mean RS.

We used the Carmel-Calibration and Haemek-Calibration cohorts for empirically measuring the effect of the number of calibration patients on the performance measures **(Supplementary Figure 7)**. Increasing the number of calibration patients yields narrower confidence intervals in both the scaling factor and subsequent performance metrics. Subsequently, for each external cohort, we reserved *k=100* patients for the calibration step, as it proved sufficient for stable calibration in our experiments, still leaving adequate data for further analyses. For Haemek and Carmel, we used the calibration patients from Carmel-Calibration and Haemek-Calibration to perform the calibration due to two advantages: (1) it allowed us to include all patients from Haemek and Carmel for subsequent analyses, and (2) the calibration cohorts consisted of consecutive diagnoses (i.e. not only patients who had Oncotype DX) and enabled to further minimize biases in the estimation of mean RS for these cohorts.

The full calibration method for applying the system in a new medical center can be summarized in the following algorithm:

(1) Extract digitized H&E slides and clinicopathologic information on *k* patients with node-negative HR+/HER2- invasive breast cancer. Clinicopathologic information includes Grade (1: low, 2: intermediate, 3: high), PR status (0 or 1), ER status (0 or 1), tumor size (1: ≤ 2cm, 2: > 2cm), and age, which were found to contribute to RS prediction. We believe using subsequent diagnoses from distinct patients over a fixed period and at least *k=100* is preferable.
(2) Apply the clinicopathologic-based model to the clinicopathologic features of the *k* calibration patients and compute the mean across the *k* results to derive an estimation for the mean of RS, Mean(*RS_estimated_*).
(3) Obtain the AI scores by applying the multimodal model to the H&E images and clinical variables of the *k* calibration patients. Compute the mean of the AI scores, *Mean*(*AI*).
(4) Compute the scaling factor 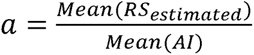
(5) For a new patient from the cohort, apply the multimodal model to obtain the AI score and multiply it by the scaling factor a to obtain the calibrated AI score.

### Clinical risk

Clinical risk stratification follows MINDACT trial criteria, which is based on a modified version of Adjuvant! Online^7^. Specifically, low-risk includes node-negative cases with either low-grade tumors ≤3 cm, intermediate-grade tumors ≤2 cm, high-grade tumors ≤1 cm, or node-positive cases with low-grade tumors ≤2 cm; high-risk encompasses all other cases with complete grade, size, and nodal status data.

### Interpretation Heatmaps

We derived heatmaps highlighting the regions the model relied on for its decisions and reflecting the local AI scores on the regions. To derive the heatmaps, we applied the model to regions of 10×10 tiles in the H&E image, treating each region as a bag of tiles in the MIL downstream task (**Supplementary Figure 13**). For each region, we derived a single score reflecting the local RS estimation, and 100 vectors of size 1,526 corresponding to the gradients with respect to each tile’s feature vector. We took the norm of the gradient vectors, resulting in 10×10 scores representing the contribution of each tile to the model’s decision-making. We then averaged the results across the slide, using a sliding window approach with a stride of 4 tiles.

### Endpoints

Time-to-event endpoints included distant recurrence-free interval probability, recurrence-free interval probability, disease-free survival (DFS) probability, overall survival probability, and breast cancer-specific survival probability. All endpoints were defined as in the TAILORx study^13^. Specifically, DFS was defined as freedom from invasive disease recurrence, second primary cancer, or death.

### Statistical Analysis

We assessed the performance of our models using the following statistical measures: area under the ROC curve (AUC), Pearson correlation coefficient, and Spearman correlation coefficient were chosen to measure the overall performance. Sensitivity, specificity, negative predictive value (NPV), and positive predictive value (PPV) were chosen to measure the potential clinical utility of the model and its ability to identify Oncotype DX’s low-risk and high-risk patients. For the Kaplan-Meier curves, P values, hazard ratios (HR), and 95% confidence intervals (CIs) were computed to quantify the effect size under the Cox proportional hazards assumption, with a significance threshold of 0.05. For performance measures and feature contribution, 95% CIs and P values were computed using bootstrapping with replacements and 1,000 replicates.

### Ethical approvals for the datasets

Data from ABCTB, TAILORx, Carmel, Haemek, Sheba, and UCMC were collected in compliance with the Helsinki Declaration and relevant institutional policies and Data Use Agreements (DUAs). Ethical approval for these datasets was granted by the respective Institutional Review Boards (IRBs). All data were de-identified. Specifically, the ABCTB dataset received ethical and scientific approval according to the Australian Breast Cancer Tissue Bank access policy is subject to ethical and scientific approvals as described in their access policy at https://nsw.biobanking.org/biobanks/view/7. For the TAILORx slides, DUA was signed with the ECOG-ACRIN Cancer Research Group. For the TAILORx clinical data, DUA was signed with the NCTN/NCORP Data Archive.

### Data availability

The data were composed of seven independent cohorts: (1) The TCGA-BRCA dataset, publicly available at https://portal.gdc.cancer.gov. (2) The ABCTB dataset, accessible from the Australian Breast Cancer Tissue Bank, is subject to ethical and scientific approvals as described in their access policy: https://nsw.biobanking.org/biobanks/view/7. (3) The TAILORx dataset can be requested from the ECOG-ACRIN Cancer Research Group and the NCTN/NCORP Data Archive. The remaining data collected from medical centers are not available for public access due to privacy and ethical considerations, in alignment with the Helsinki agreements and institutional policies. Interested researchers may request access directly from the respective institutions: (4) Carmel data from the Carmel Medical Center, Israel. (5) Haemek data from the Haemek Medical Center, Israel. (6) Sheba data from Sheba Medical Center, Israel. (7) UCMC data from the University of Chicago Medical Center, USA. Sample sizes and descriptions for each cohort are provided throughout the manuscript. Data used in this study were collected in accordance with relevant regulations and ethical approvals from the respective institutions.

### Code availability

The source code used in this study has been archived in Zenodo^68^ and can be accessed at the following link: https://github.com/shachar5020/TransformerWSI4OncoDXPrediction. Statistical analysis was performed using Matlab 2024b and R 4.4.0. Preprocessing, tuning of the models, and inference were done using Python 3.10.6 and Pytorch library version 1.12.1. Additional Python libraries used for database management, graphical plotting, scientific calculations, and other tasks include Numpy v.1.23.1, Pandas v.1.4.4, Scipy v.1.9.1, Pytorch Lightning v.1.9.3, and Openslide v.3.4.1.

## Acknowledgments

This research was supported by the Israel Innovation Authority—Kamin 69997 (R.K. and G.S.), the Zimin Institute for Artificial Intelligence Solutions in Healthcare grant (R.K. and G.S.), and the Israel Precision Medicine Partnership program (IPMP) grant 3864/21 (R.K. and G.S.). We would like to thank Karin Stoliar for helping with the data acquisition and quality assurance, Hen Davidov for supporting the deep learning experiments, and Liat Dizengoff for managing the Helsinki approvals in Carmel Medical Center. This manuscript was prepared using data from Datasets [PACCT-1] from the NCTN/NCORP Data Archive of the National Cancer Institute’s (NCI’s) National Clinical Trials Network (NCTN). Data were originally collected from clinical trial NCT number [NCT00310180] [Program for the Assessment of Clinical Cancer Tests (PACCT-1): Trial Assigning Individualized Options for Treatment: The TAILOR Trial]. All analyses and conclusions in this manuscript are the sole responsibility of the authors and do not necessarily reflect the opinions or views of the clinical trial investigators, the NCTN, the NCORP or the NCI. TAILORx was conducted by the ECOG-ACRIN Cancer Research Group (Peter J. O’Dwyer, MD and Mitchell D. Schnall, MD, PhD, Group Co-Chairs) and supported by the National Cancer Institute of the National Institutes of Health under award numbers U10CA180820 and U10CA180794. The content is solely the responsibility of the authors and does not necessarily represent the official views of the National Institutes of Health. Additional support was provided by the Breast Cancer Research Foundation under awards CONS-21-007, CONS-20-006, CONS-19-006, and CONS-18-007.

## Contributions

G.S. conceptualized the study and designed the experiments. G.S. and D.A. conducted the statistical analysis. G.S. and S.C. collaborated on data collection, data processing, and machine learning model development. E.S., A.C., T.G., G.B.S., C.M., and A.P. contributed to data collection, annotation, and pathology data interpretation. G.S., D.A., and R.K. supervised the project. All authors contributed to drafting the manuscript and provided critical revisions.

---

**Supplementary Table 1:**
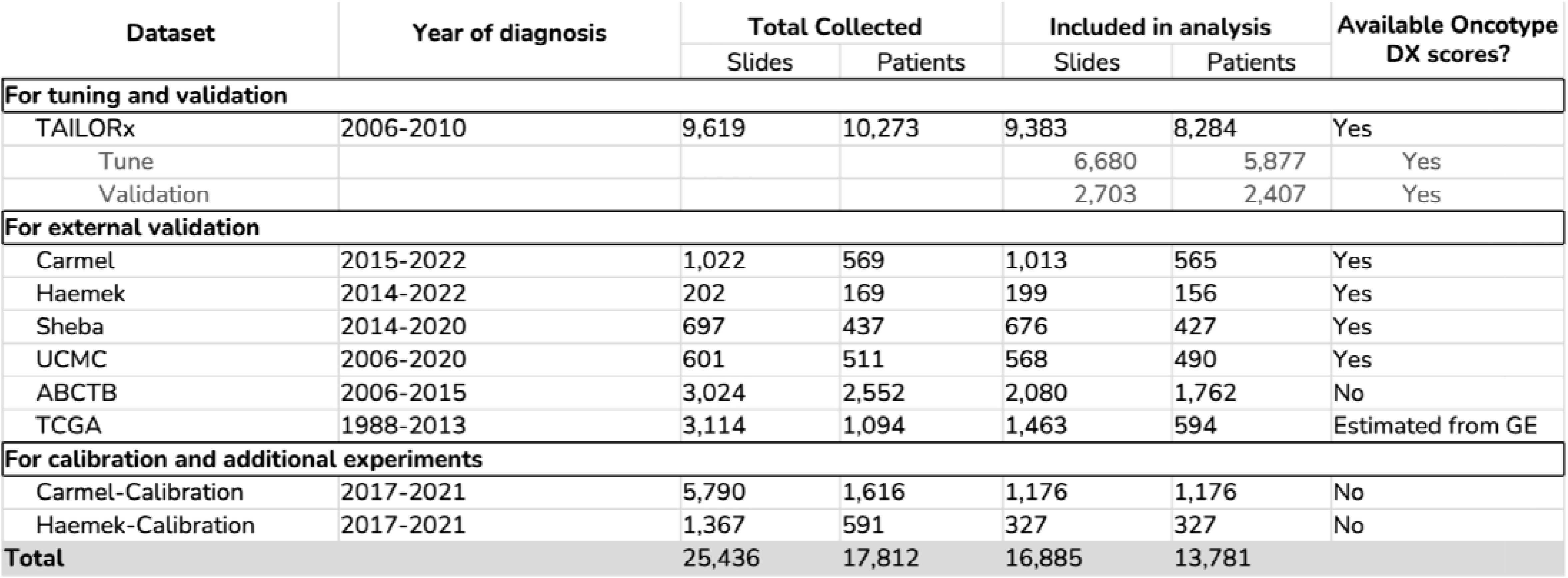
Overview of study cohorts and patient inclusion. Summary of all cohorts used in the study, including TAILORx and the external validation cohorts. For each cohort, we present the years of diagnosis, the total number of collected slides and patients, and the final numbers included in the analysis. The table is organized into three sections: (1) TAILORx data used for tuning using cross-validation and for validation on a held-out validation set. (2) External validation cohorts - comprising three Israeli medical centers (Carmel, Haemek, Sheba), two from the United States (UCMC, TCGA), and one from Australia (ABCTB). (3) Additional calibration cohorts - used for validating the distribution-matching calibration method and for calibrating the model for Carmel and Haemek. For TAILORx, Carmel, Haemek, Sheba, and UCMC, Oncotype DX scores were available from clinical testing. For TCGA, RS values were computationally estimated from RNA sequencing gene expression (GE) data. Detailed inclusion/exclusion criteria and quality control steps are shown in Supplementary Figure 1.

**Supplementary Table 2:**
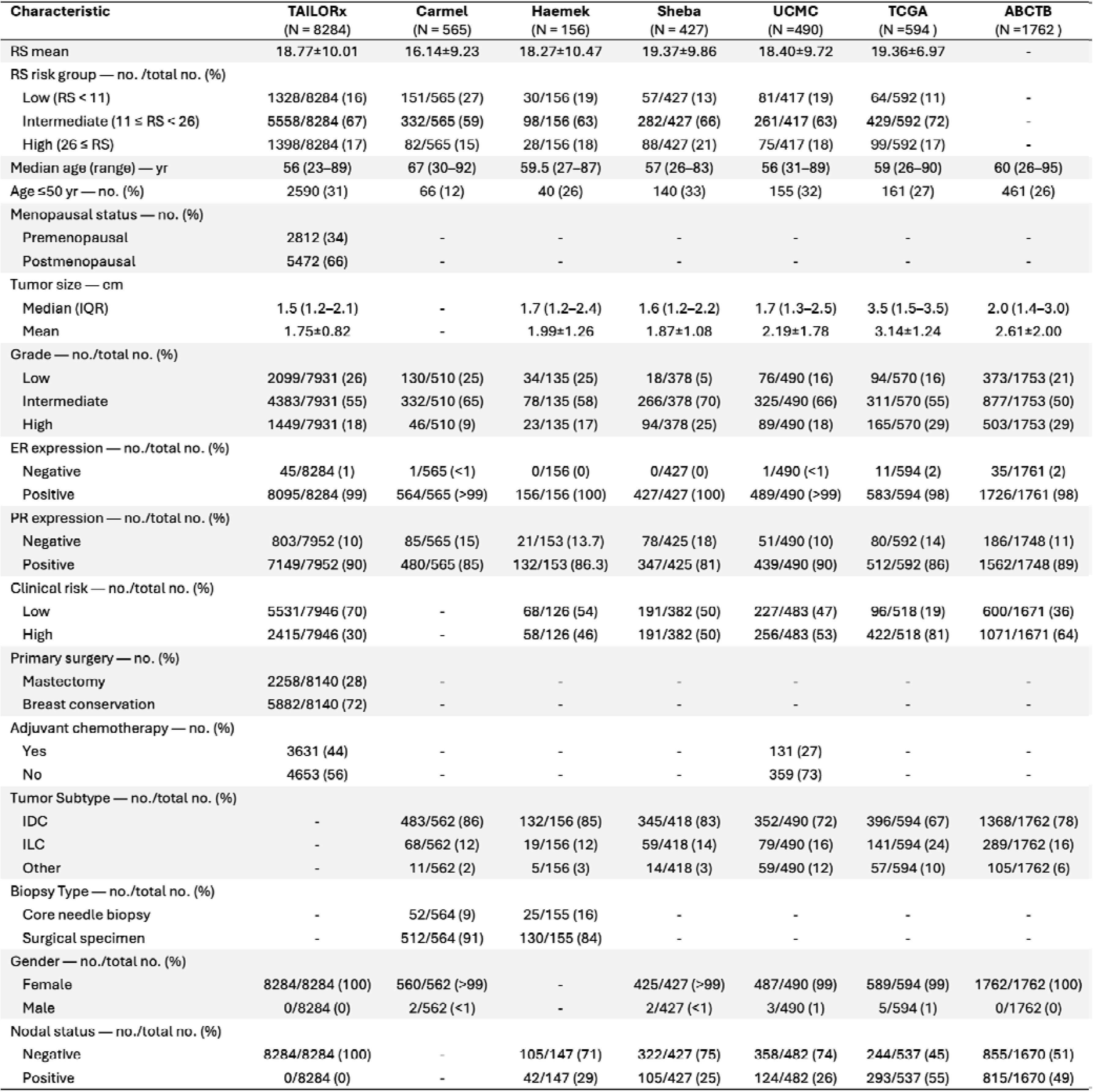
Clinicopathological characteristics across study cohorts. Comparison of patient and tumor characteristics across all study cohorts. For continuous variables (age, tumor size, RS), data are presented as means ±SD or median (range) where appropriate. For categorical variables, data are shown as the number (percentage) of patients. Clinical risk stratification follows MINDACT trial criteria: low-risk includes node-negative cases with either low-grade tumors ≤3 cm, intermediate-grade tumors ≤2 cm, high-grade tumors ≤1 cm, or node-positive cases with low-grade tumors ≤2 cm; high-risk encompasses all other cases with complete grade, size, and nodal status data. Missing data fields are indicated by dashes (−). Some characteristics were not consistently available across all cohorts, reflecting real-world data collection practices. Tumor subtypes are classified as IDC (invasive ductal carcinoma), ILC (invasive lobular carcinoma), or other histological types. IQR: interquartile range.

**Supplementary Table 3:**
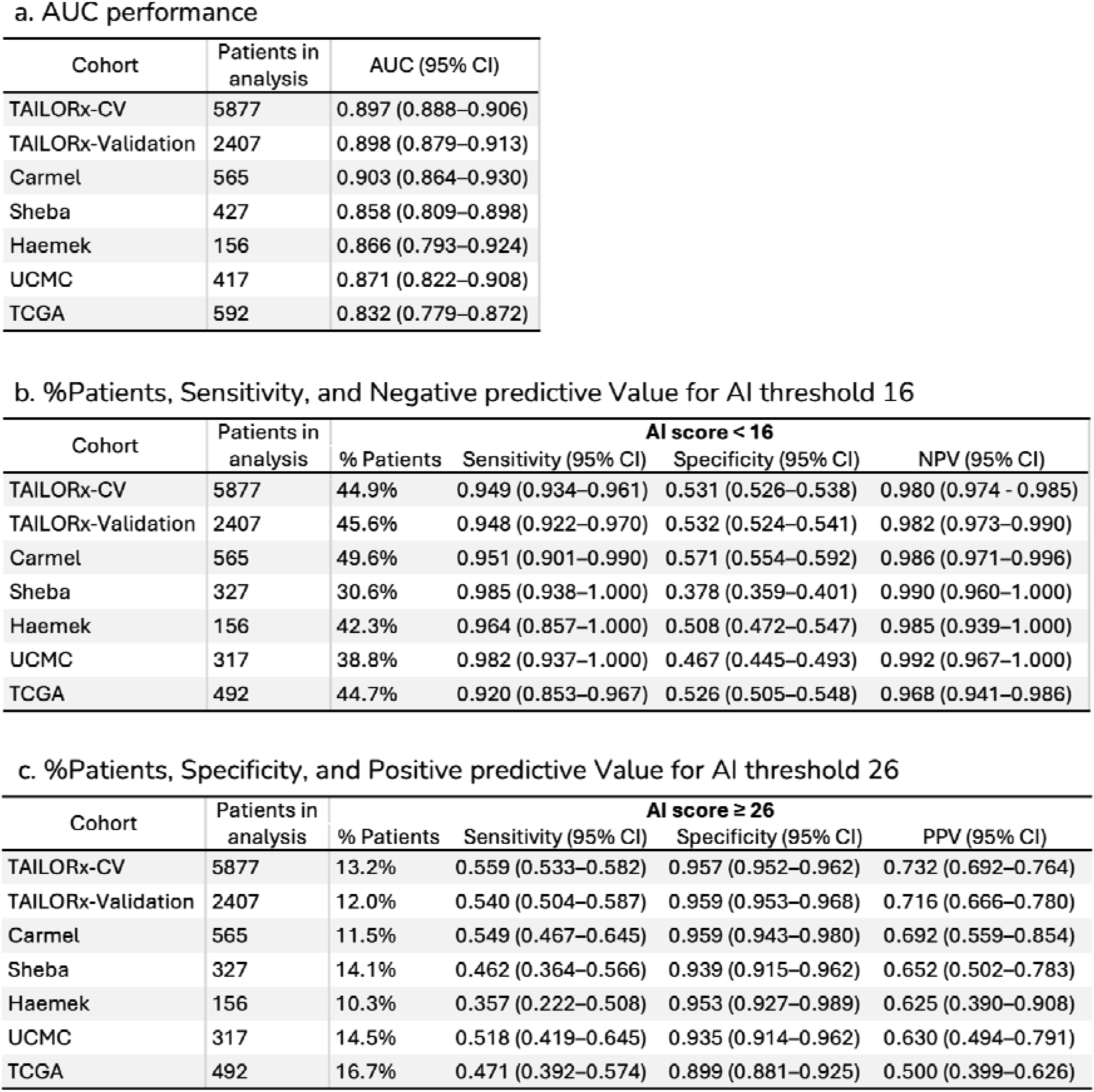
Performance metrics of the AI multimodal model across all cohorts, for identifying high genomic risk (RS≥26). Summary of AI multimodal model performance across the cohorts, presented in three complementary analyses: (a) Discrimination performance measured by area under the ROC curve (AUC) with 95% confidence intervals (CI) for identifying high genomic risk (RS≥26). TAILORx-CV refers to the average performance of the five cross-validation AI models on their corresponding validation folds, in the TAILORx tuning set. For the external cohorts, patients in this analysis also included patients used for calibration since calibration does not affect ranking-based metrics. (b) Low-risk classification performance (AI<16), reporting the percentage of patients classified as low AI risk, sensitivity, specificity, and negative predictive value (NPV) with 95% CIs for identifying RS≥26. This threshold represents patients who could potentially avoid chemotherapy. (c) High-risk classification performance (AI≥26), showing the percentage of patients classified as high AI risk, sensitivity, specificity, and positive predictive value (PPV) with 95% CIs for identifying RS≥26. This threshold represents patients who might benefit from immediate chemotherapy initiation. Calibration-set patients (n=100 each) were excluded from analyses in (b) and (c) for UCMC, Sheba, and TCGA to ensure an unbiased evaluation of threshold-dependent metrics. AI: artificial intelligence, representing the multimodal model.

**Supplementary Table 4:**
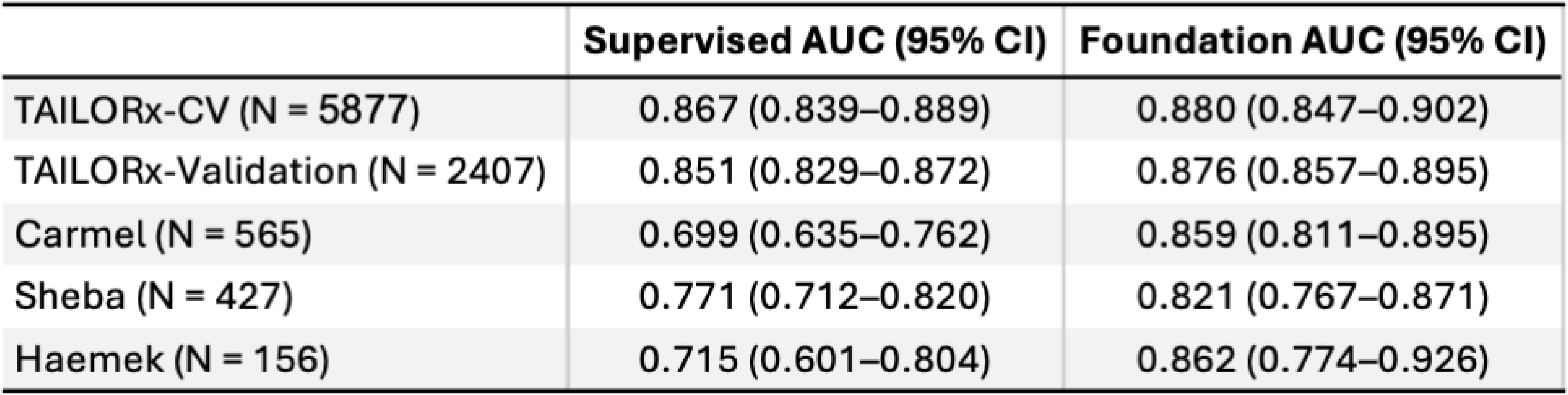
Comparative analysis of foundation model versus traditional supervised learning approaches. Performance comparison between our SSL-based foundation approach and a fully supervised learning model. Both models were tuned on the TAILORx training set to classify cases as low-risk (RS<26) or high-risk (RS≥26) from H&E images. TAILORx-CV refers to the average performance of the five cross-validation AI models on their corresponding validation folds, in the TAILORx tuning set. The supervised model follows the architecture and training protocol described in ^29^. The table presents the area under the ROC curve (AUC) with 95% confidence intervals for both approaches across TAILORx cross-validation, validation set, and all external validation cohorts. Despite having access to the same large-scale tuning data, the foundation model demonstrates consistently superior performance, particularly in external validation, highlighting the benefits of transfer learning from a trained foundation model for robust generalization across different clinical settings.

**Supplementary Figure 1:**
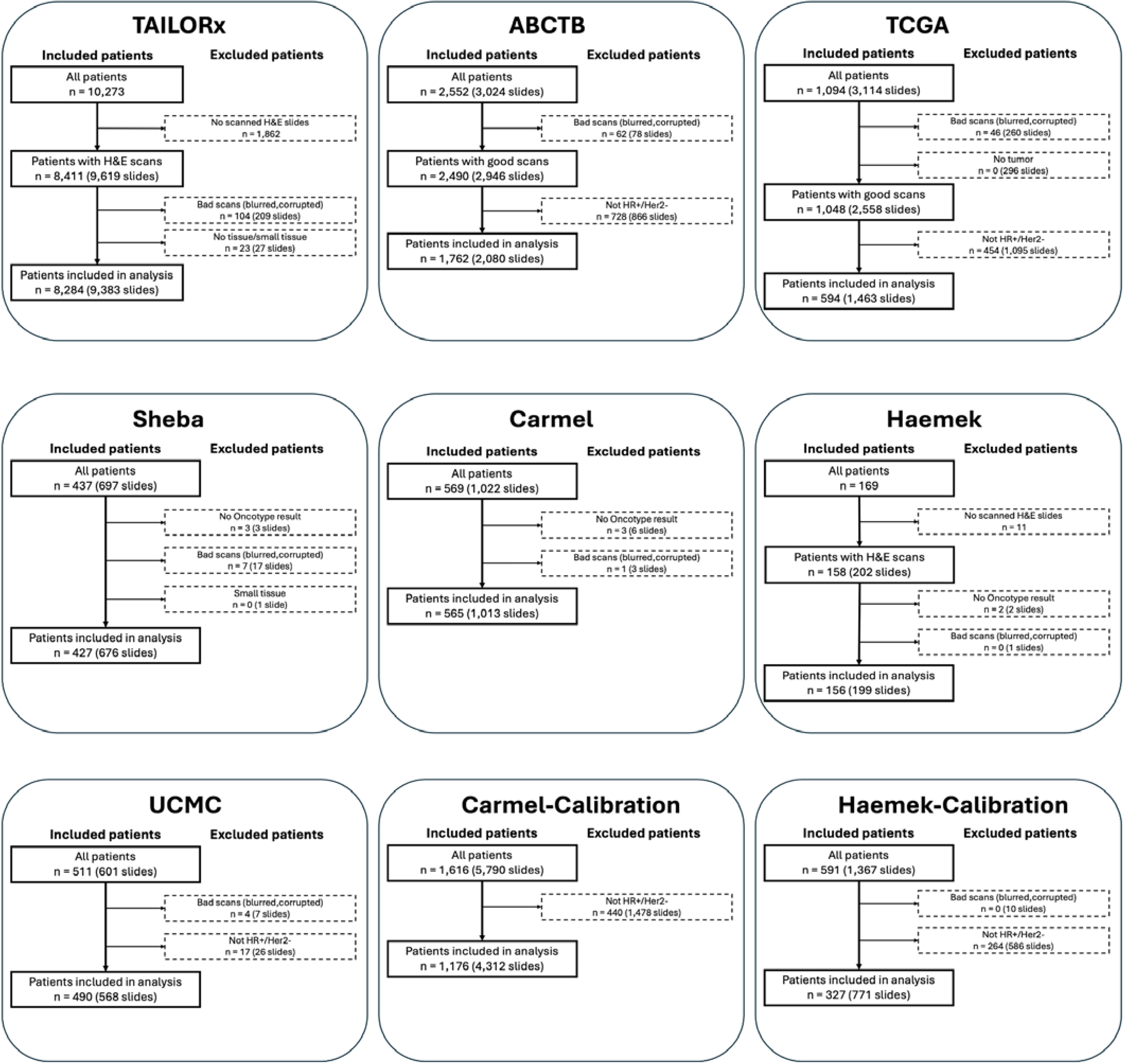
Cohort inclusion criteria and patient flow diagrams. Flow diagrams showing patient inclusion and exclusion criteria for all study cohorts. For each cohort, we show the initial number of patients and slides collected, followed by sequential exclusion steps with specific reasons and numbers. Starting with TAILORx (N=10,273 patients), we detail exclusions due to missing H&E slides and quality control failures (blurred/corrupted scans, tissue-deficient slides). For the external validation cohorts, we show a similar workflow for Carmel (N=569), Sheba (N=437), Haemek (N=169), UCMC (N=511), ABCTB (N=2,552), and TCGA (N=1,094). The calibration cohorts (Carmel-Calibration and Haemek-Calibration) had different inclusion criteria as they did not require matched RS data. Quality control criteria were consistently applied across all cohorts, focusing on slide scan quality and tissue adequacy. Final numbers of included patients and slides are provided for each cohort. HR: hormone receptor status.

**Supplementary Figure 2:**
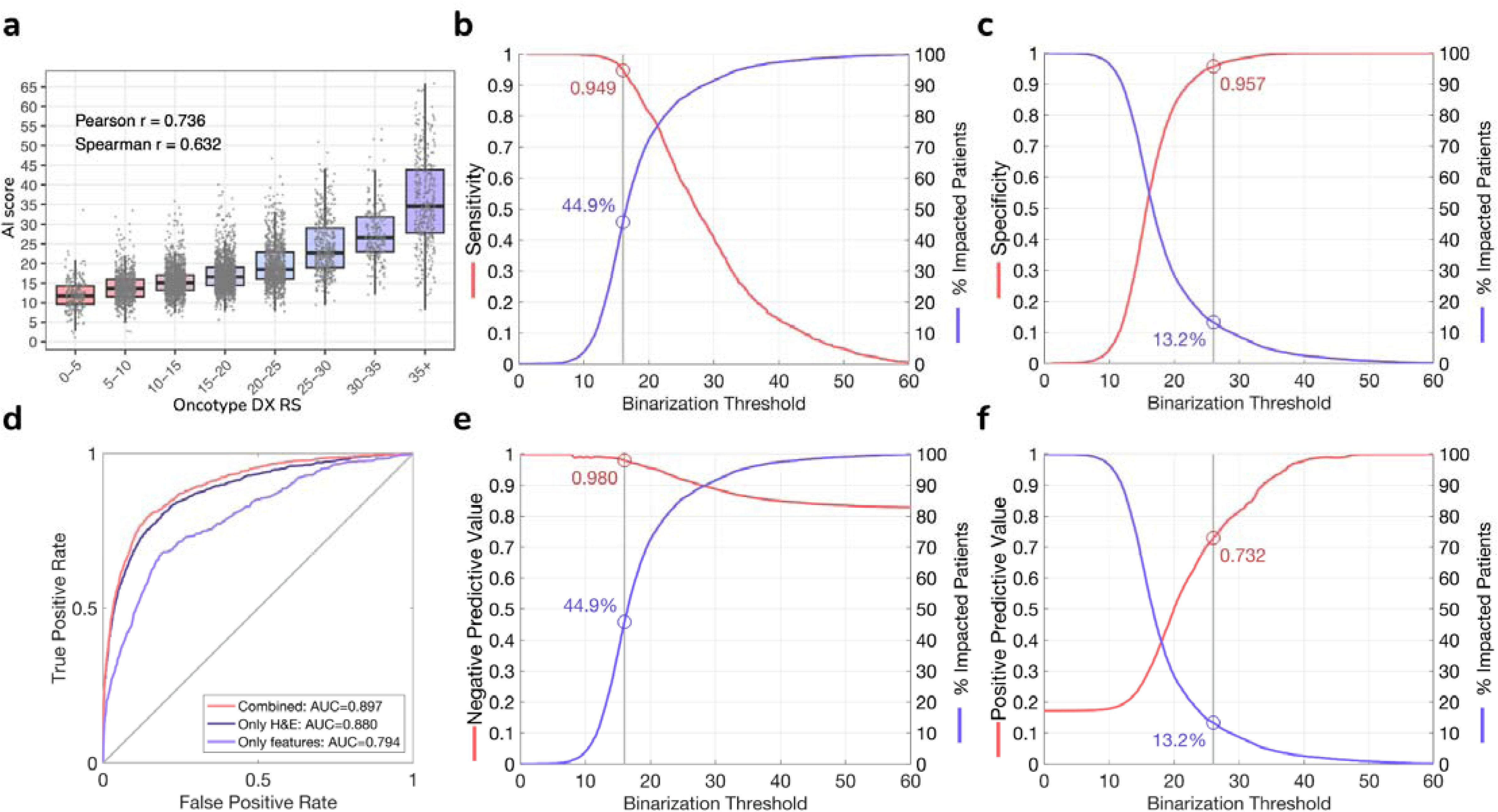
Performance evaluation of the deep learning models on the TAILORx cross-validation. Model performance assessment during five-fold cross-validation on the TAILORx cohort (n=5,877 patients). **(a)** Distribution of the AI multimodal model’s scores versus Oncotype DX RS on the TAILORx, showing high correlation (Pearson r=0.736, Spearman r=0.652). **(b)** Clinical utility assessment for low AI risk identification, showing sensitivity and proportion of impacted patients across AI score thresholds. Using a low classification threshold of 16, the model classifies 44.9% of patients as low AI risk with 94.9% sensitivity for identifying RS≥26. **(c)** Clinical utility assessment for high-risk identification, showing specificity and proportion of impacted patients across AI score thresholds. Using a high classification threshold of 26, the model classifies 13.2% of patients as high AI risk with 95.7% specificity for identifying RS≥26. **(d)** Receiver operating characteristic (ROC) curves for identifying RS≥26, comparing the multimodal model combining both H&E images and clinicopathologic variables (AUC=0.897), the image model using only H&E images (AUC=0.880), and the clinicopathologic model using only the features (AUC=0.794). **(e-f)** Negative Predictive Value (NPV) and Positive Predictive Value (PPV) for the low and high classification thresholds.

**Supplementary Figure 3:**
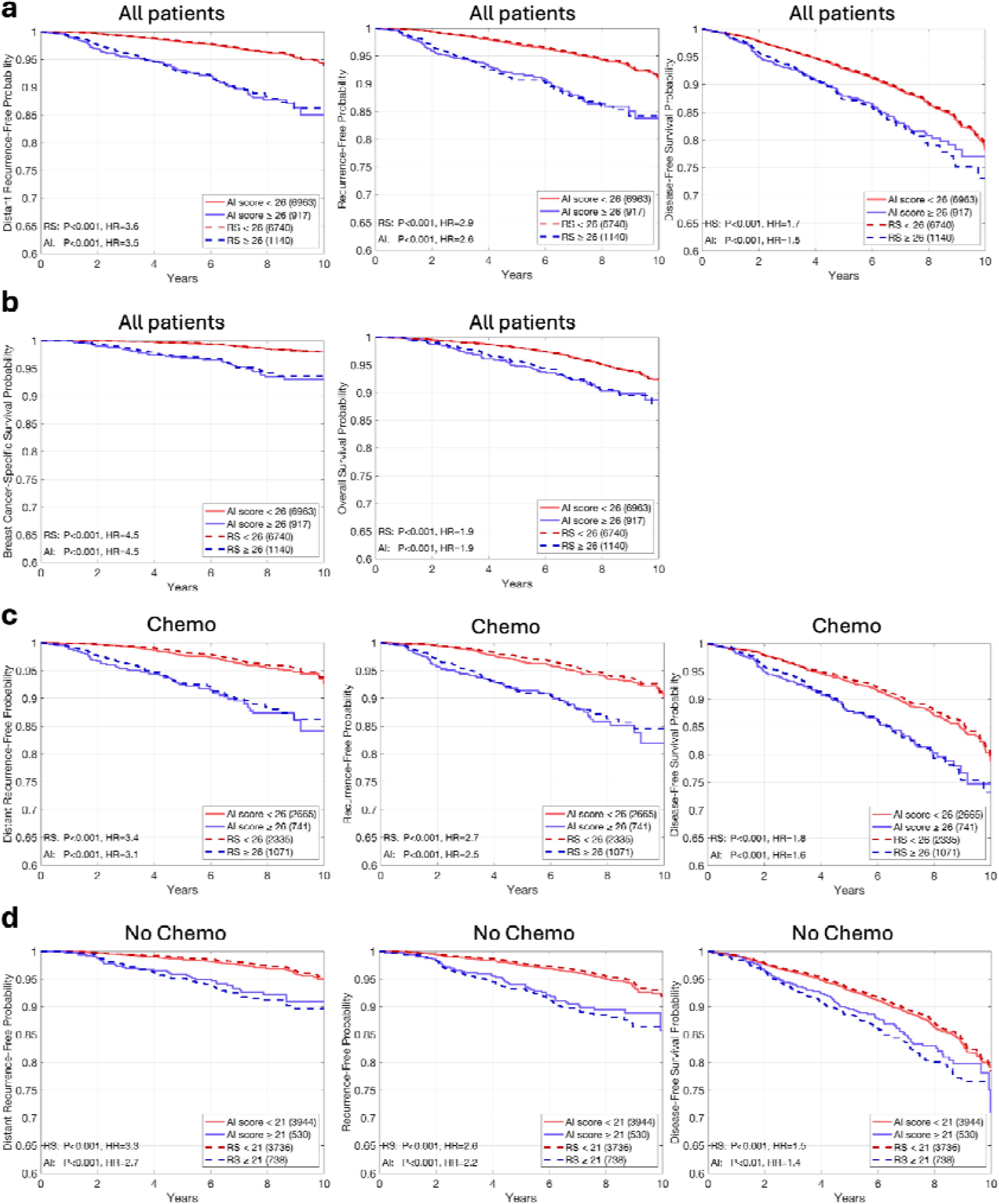
Extended survival analysis when including cross-validation patients. Kaplan-Meier plots comparing patient stratification using recurrence scores (RS) versus AI scores in the entire TAILORx cohort, including both validation set and cross-validation patients. **(a)** Overall cohort analysis shows distant recurrence-free interval (DRFI), recurrence-free interval (RFI), and disease-free survival (DFS) probabilities using a stratification threshold of 26. **(b)** Additional endpoints: breast cancer-specific survival and overall survival. **(c)** Analysis of the same endpoints in patients who received chemotherapy. **(d)** Corresponding analysis for patients who did not receive chemotherapy, using a classification threshold of 21. For each subgroup analysis, patient numbers, P values for the stratification, and hazard ratio (HR) values are shown in the legends. The consistent alignment between survival curves derived from RS and AI scores across all analyses demonstrates the robust prognostic ability of the model.

**Supplementary Figure 4:**
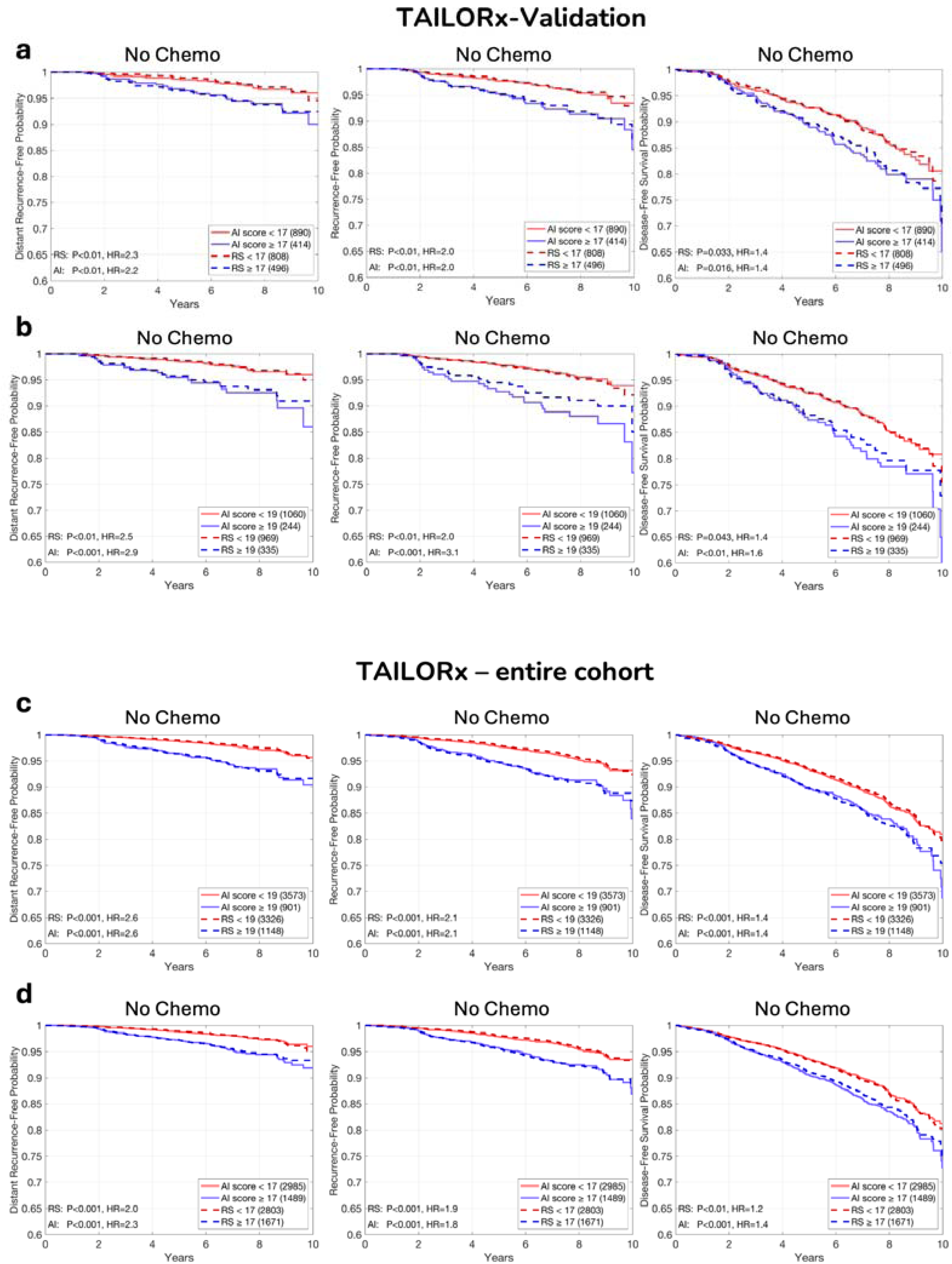
Extended survival analysis with additional classification thresholds. Kaplan-Meier plots comparing patient stratification using recurrence scores (RS) versus AI scores in TAILORx patients who did not receive chemotherapy. This figure complements Figure 2c and Supplementary Figure 3d by evaluating additional classification thresholds (17 and 19) beyond the primary threshold of 21. For each endpoint (distant recurrence-free interval, recurrence-free interval, and disease-free survival) and threshold value, patient stratification is shown using both RS and AI scores. Each Kaplan-Meier plot includes the number of patients per risk group and hazard ratios (HR) with corresponding P-values from log-rank tests. The consistent performance across multiple thresholds demonstrates the robustness of the AI model’s prognostic capability at various decision points.

**Supplementary Figure 5:**
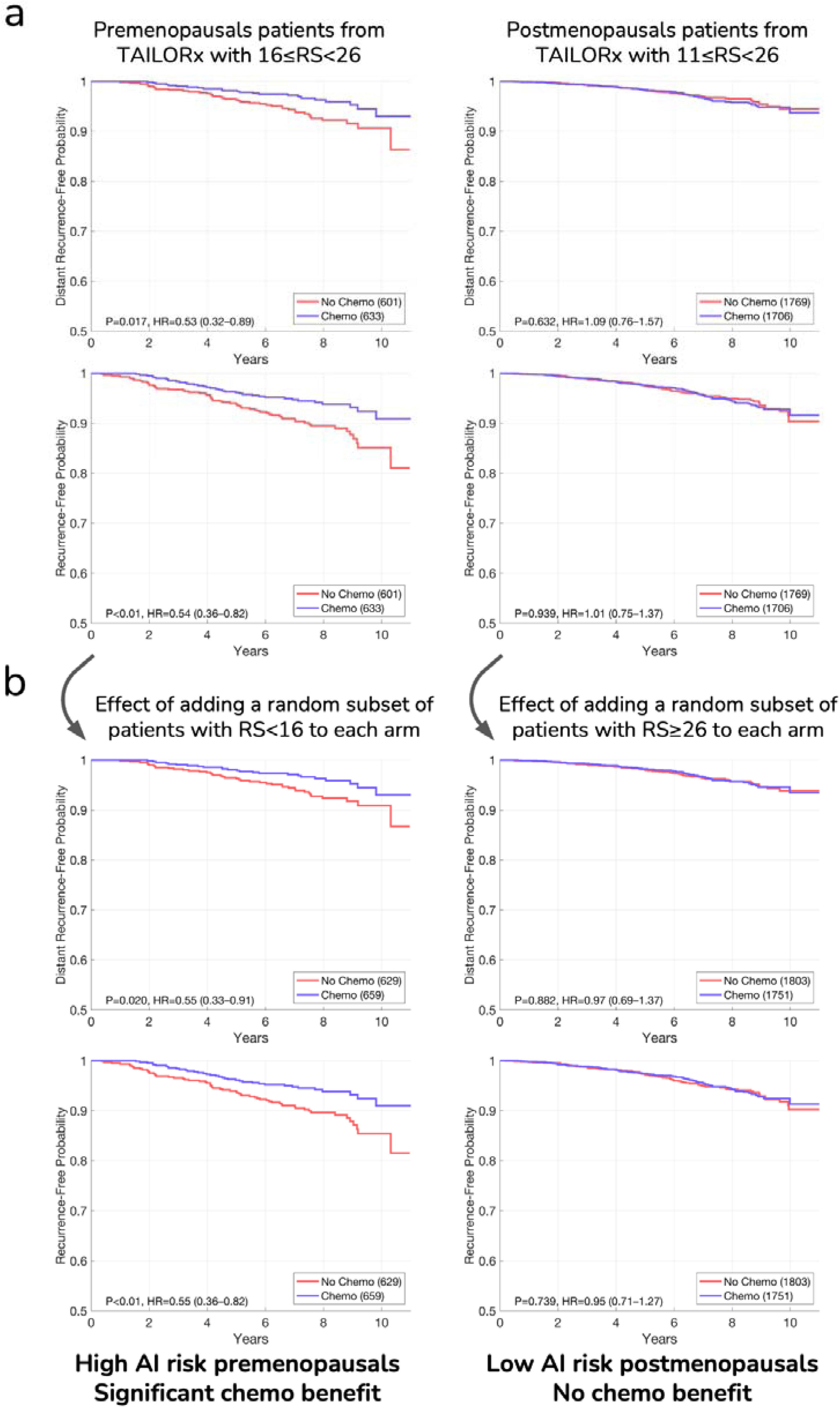
Chemotherapy Benefit for Pre- and Post-Menopausals with Intermediate RS: Additional Endpoints. This figure complements Figure 3 in the main paper, using additional endpoints: distant recurrence-free interval and recurrence-free interval.

**Supplementary Figure 6:**
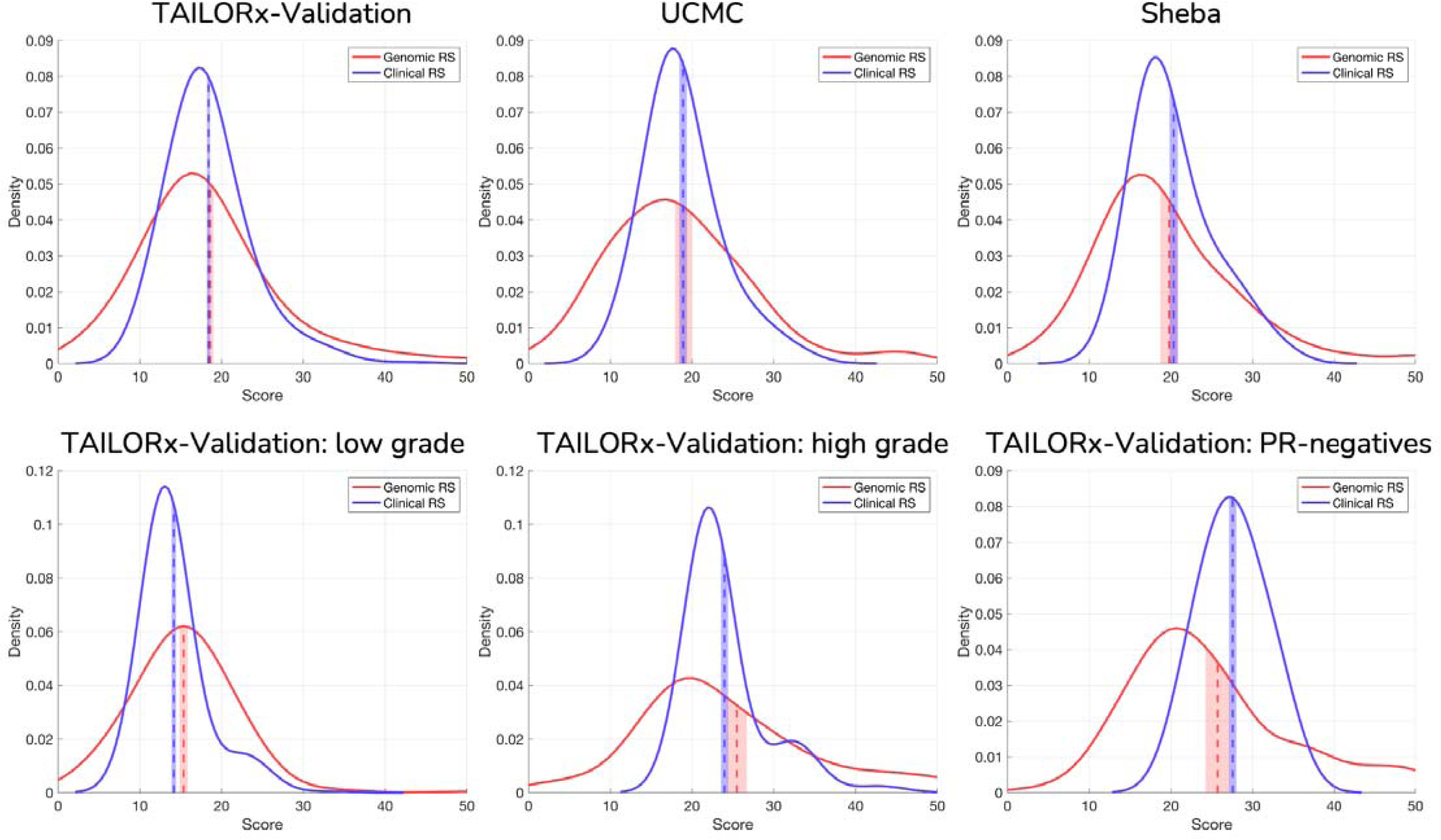
Distribution analysis of genomic and model-estimated recurrence scores. Probability density distribution analysis comparing Oncotype DX RS (‘genomic RS’) and estimation of RS using the clinicopathologic model, which was trained using only clinical features (‘Clinical RS’). **Top row:** Distribution comparison for the TAILORx validation set (n=2,407), UCMC (n=490), and Sheba (n=427) cohorts. **Bottom row:** Analysis of specific patient subgroups within the TAILORx validation set, stratified by histological grade (low-grade and high-grade) and PR status (PR-negative). For each plot, means are displayed with their 95% confidence intervals (vertical dashed lines), and kernel density estimation curves show the full distribution of scores. Despite differences in the overall shape of distributions between genomic and estimated RS, the mean values show strong concordance. This analysis demonstrates that while the clinicopathologic model may not accurately infer the RS, it reliably captures its average risk score across different cohorts and patient subgroups.

**Supplementary Figure 7:**
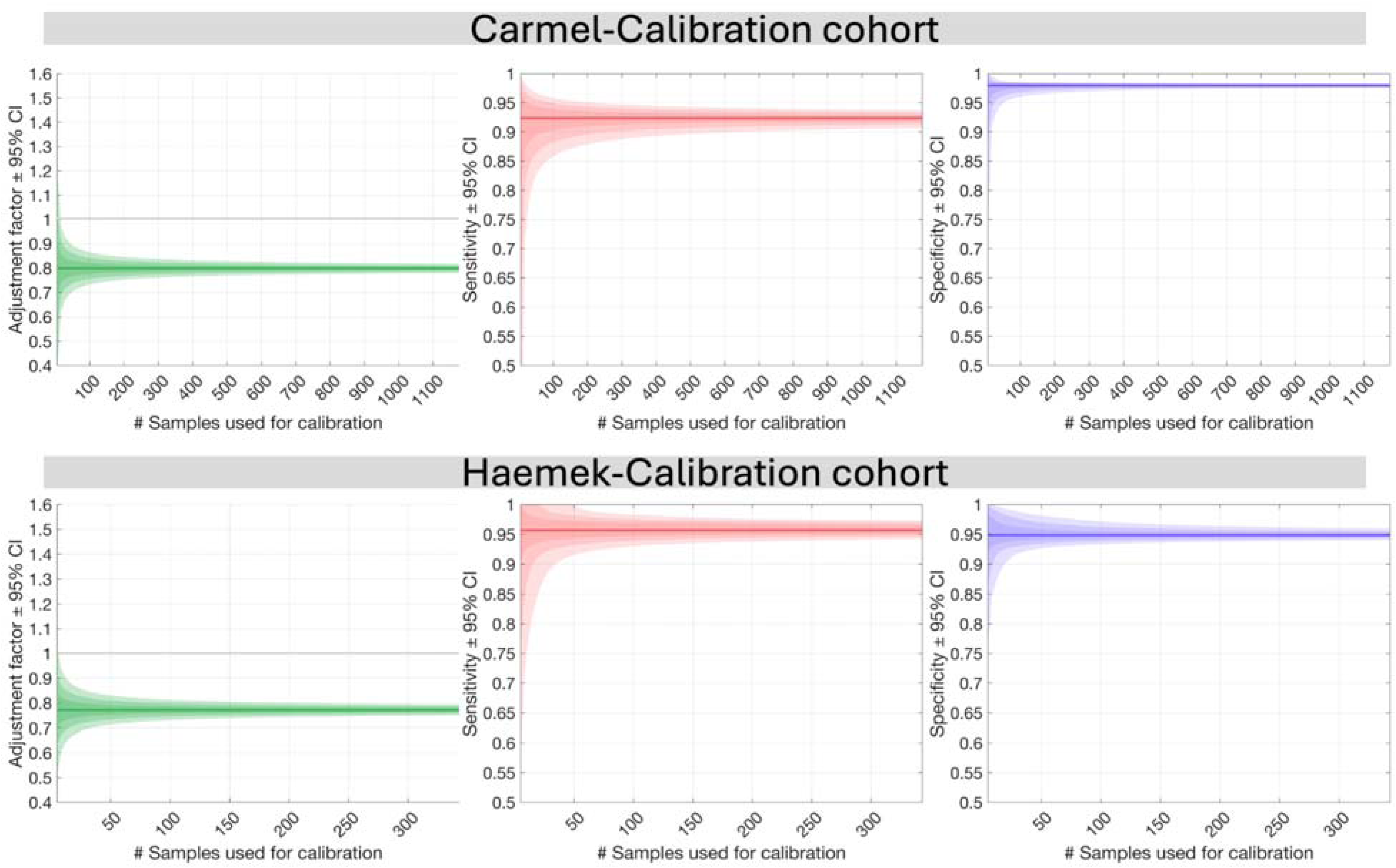
analysis of calibration sample size requirements. Investigation of how the number of samples used for calibration affects model performance. Results are shown for two independent calibration cohorts: Carmel-Calibration (n=1,176) and Haemek-Calibration (n=327). For each cohort, three key metrics are analyzed as a function of calibration sample size. **Left**: Adjustment factor with 95% confidence intervals (CI), showing the stability of the calibration coefficient. **Middle**: Sensitivity for identifying RS≥26 on Carmel and Haemek cohorts using the low classification threshold (16), along with 95% CIs. **Right**: Specificity for identifying RS≥26 on Carmel and Haemek cohorts using the high classification threshold (26), along with 95% CIs. Results demonstrate that the CIs become smaller with more samples used for calibration. We used 100 patients from each cohort for calibrating our multimodal model. The consistency across both calibration cohorts, despite their different sizes and patient characteristics, validates the robustness of our calibration approach. These calibration samples do not require matched genomic testing facilitating implementation in resource-limited settings. results,

**Supplementary Figure 8:**
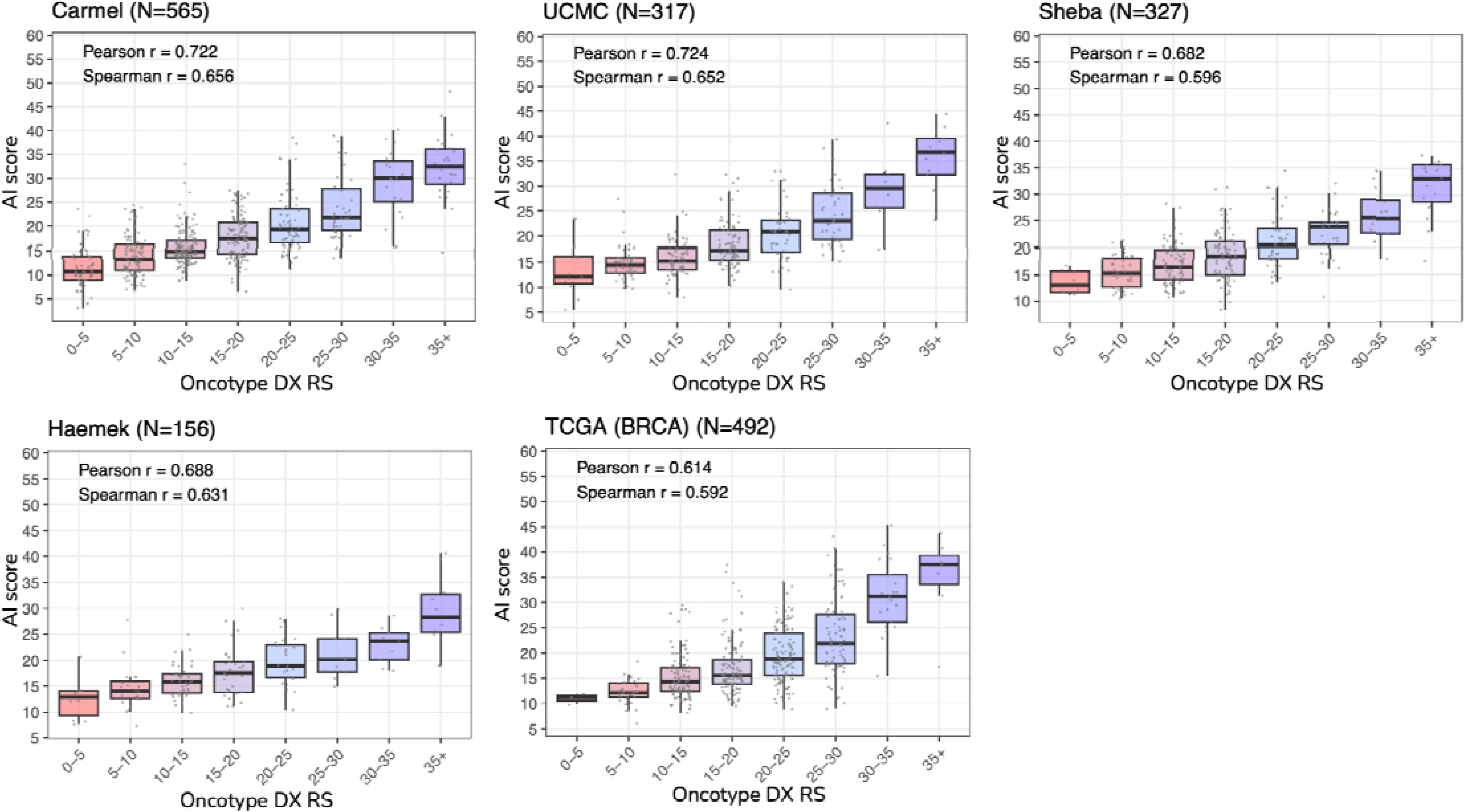
Correlation between predicted and genomic recurrence scores across external validation cohorts. Box plots demonstrating the relationship between AI scores and Oncotype DX RS values across the five external validation cohorts: Carmel (N=565), UCMC (N=317), Sheba (N=327), Haemek (N=156), and TCGA (N=492). For each cohort, the AI score distributions are shown as box plots grouped by RS ranges, with Pearson (r) and Spearman (ρ) correlation coefficients displayed. The boxes show quartiles (25th, 50th, and 75th percentiles), whiskers extend to 1.5 times the interquartile range, and outliers are shown as individual points. The strong correlation between the AI score and RS values is maintained across all cohorts (Pearson r ranging from 0.688 to 0.754), despite differences in patient populations, timeframes, and laboratory protocols. For TCGA, RS values were estimated from RNA sequencing data, as direct Oncotype DX results were not available.

**Supplementary Figure 9:**
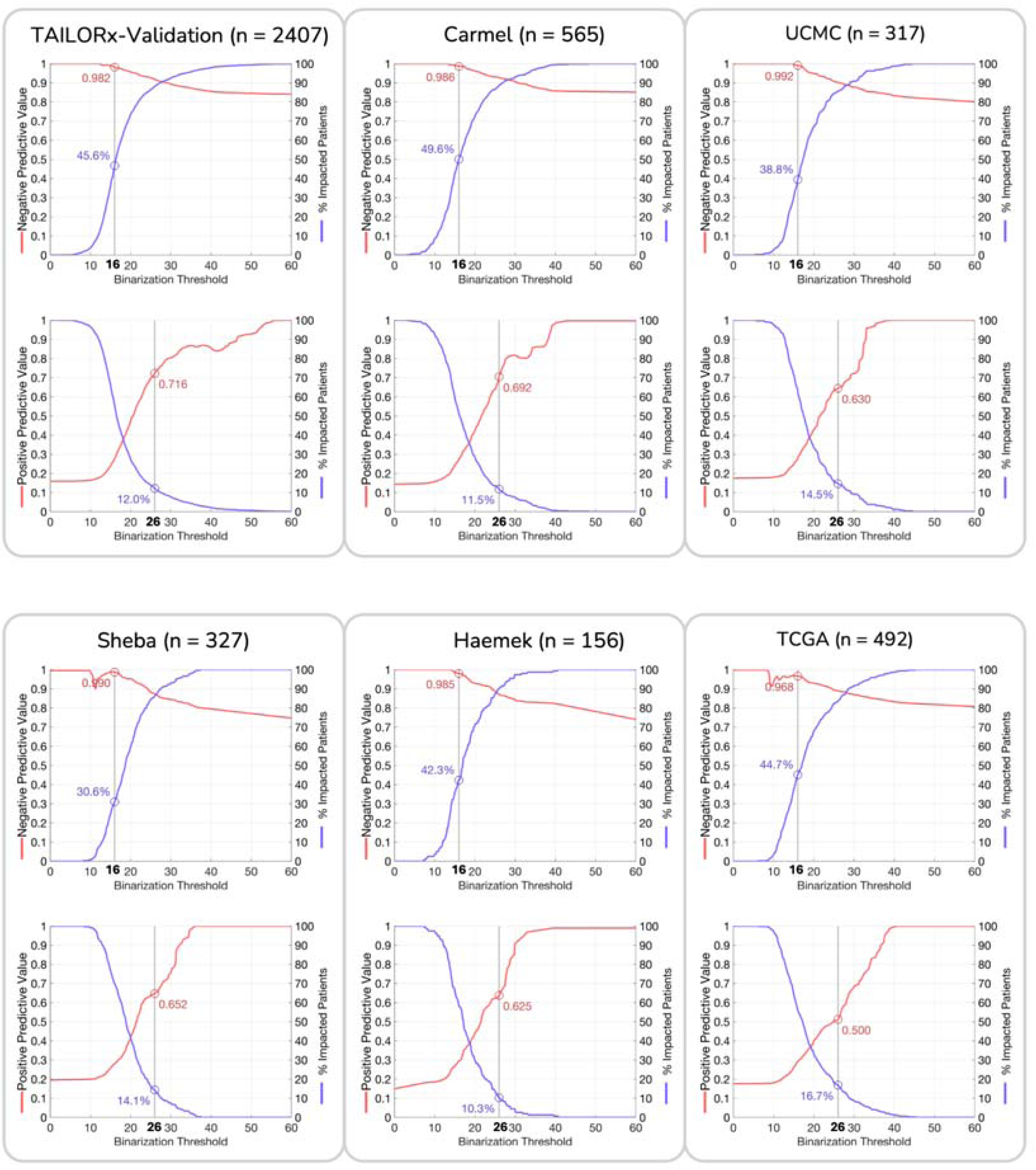
Negative and Positive Predictive value analysis of AI model thresholds across validation cohorts. Assessment of the negative and positive predictive values across all validation cohorts: TAILORx validation set (n=2,407), Carmel (n=565), UCMC (n=317), Sheba (n=327), Haemek (n=156), and TCGA (n=492). **Left panels**: Negative predictive value (NPV) analysis for identifying low genomic risk disease (RS<26), showing the relationship between NPV and percentage of patients classified below each threshold, with an operating point at 16 highlighted. **Right panels**: Positive predictive value (PPV) analysis for identifying high genomic risk disease (RS≥26), showing the relationship between PPV and the percentage of patients classified above each threshold, with an operating point at 26 highlighted. The consistency of negative and positive predictive values across cohorts demonstrates the robustness of these decision thresholds. The plots show that substantial proportions of patients (30%–50% for low AI risk and 10–17% for high AI risk) can be classified with high confidence across all validation sites. For TCGA, RS values were estimated from gene expression data as direct Oncotype DX results were not available.

**Supplementary Figure 10:**
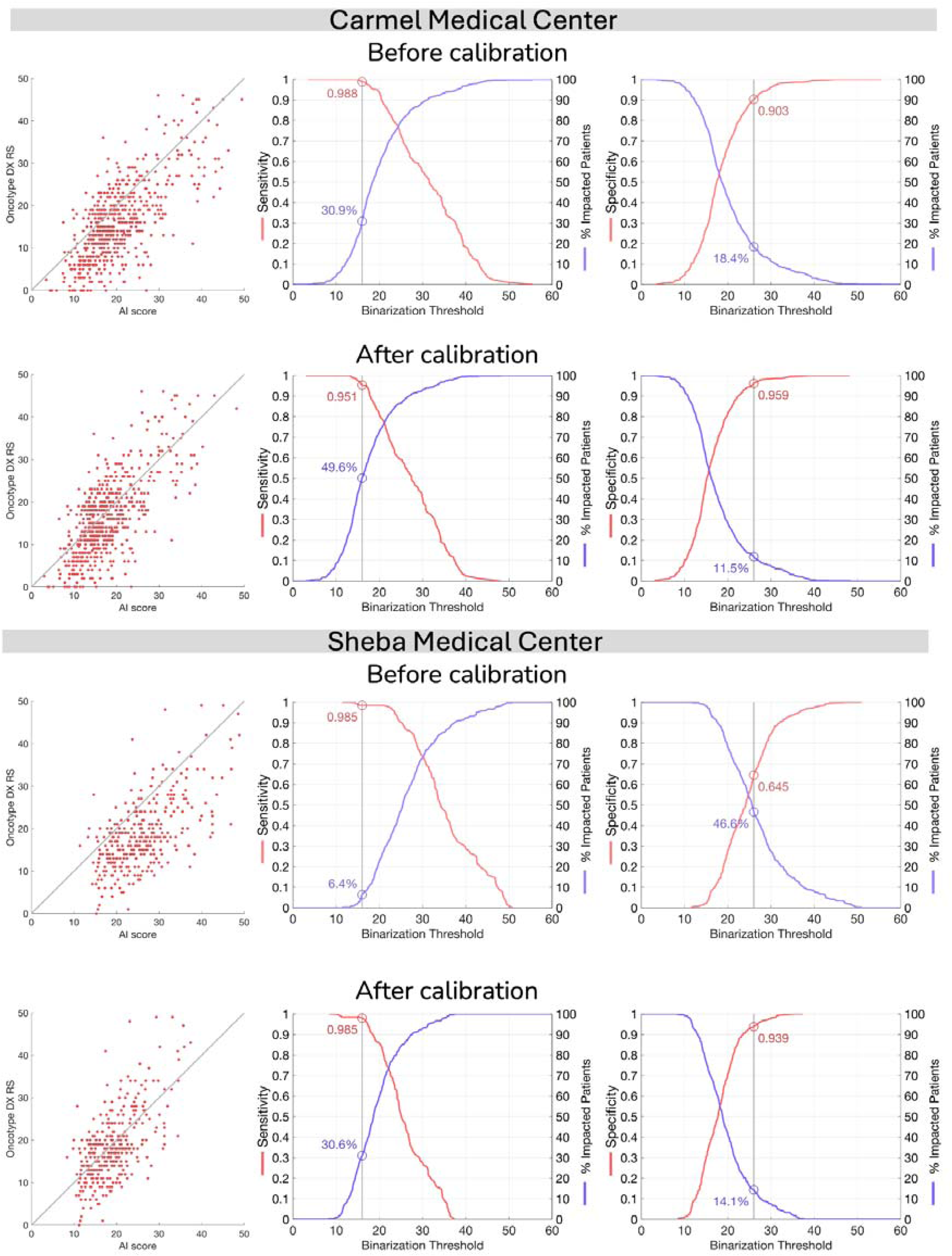
Impact of calibration on AI model performance at Carmel and Cohorts. Comparative analysis of AI scores before and after applying the distribution-matching calibration method at two independent validation sites. Results for both Carmel Medical Center and Sheba Medical Center are presented in three complementary visualizations. **Left**: Scatter plots of the AI scores versus Oncotype DX RS, demonstrating correlation patterns. **Middle**: Sensitivity for identifying RS≥26 showing the proportion of patients with AI scores below each threshold, with an operating point at 16 indicated. **Right**: Specificity for identifying RS≥26 showing the proportion of patients with AI scores above each threshold, with an operating point at 26 highlighted. For each site, the top row shows uncalibrated AI scores, while the bottom row shows results after applying calibration using 100 unlabeled samples. The calibration process maintains ranking-based performance metrics (correlation coefficients) while adjusting AI score distributions to match the development cohort, resulting in more consistent clinical decision thresholds across institutions. The number of analyzed patients (n) and performance metrics are shown for each condition.

**Supplementary Figure 11:**
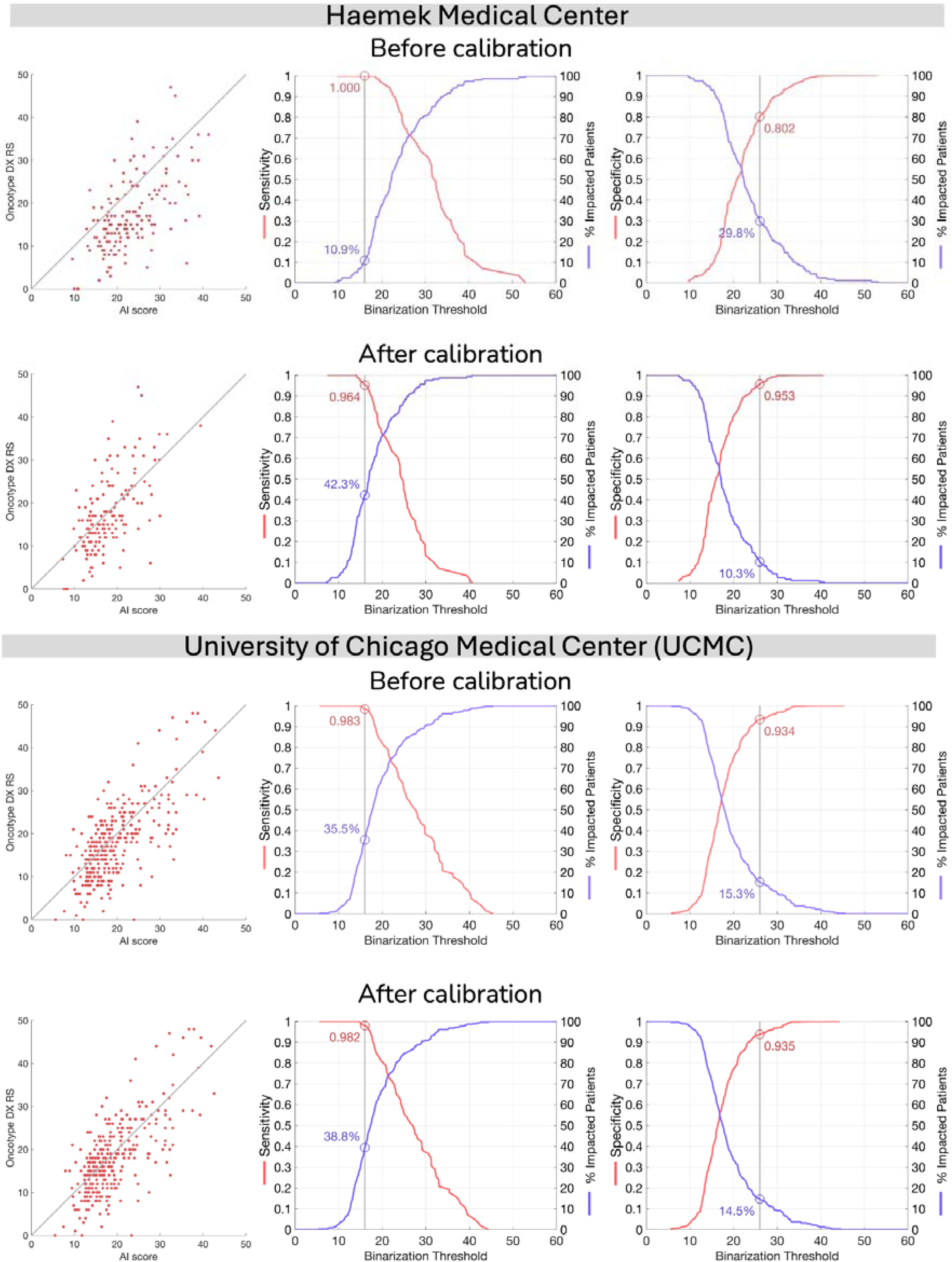
Impact of calibration on AI model performance at Haemek and Cohorts. Comparative analysis of AI scores before and after applying the distribution-matching calibration method at Haemek Hospital and UCMC. Following the same format as Supplementary Figure 10.

**Supplementary Figure 12:**
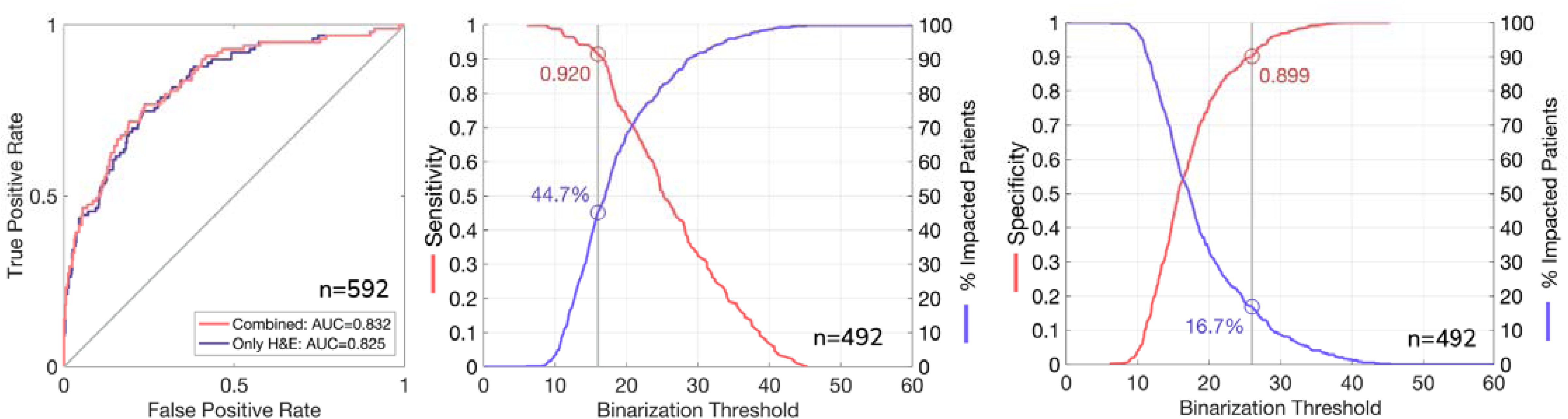
Performance evaluation on TCGA breast cancer cohort using RNA-derived recurrence scores. Performance analysis of the multimodal model on The Cancer Genome Atlas (TCGA) breast cancer cohort, where recurrence scores (RS) were computationally estimated from RNA sequencing data (Methods). **Left**: Receiver operating characteristic (ROC) curve for identifying high-risk disease (RS≥26), comparing the performance of the combined multimodal model (AUC=0.832) versus H&E images alone (AUC=0.825). **Middle**: Sensitivity analysis showing the proportion of patients classified as low-risk at different thresholds, with 44.7% of patients classified with a score below 16 at 92.0% sensitivity. **Right**: Specificity analysis showing the proportion of patients classified as high-risk at different thresholds, with 16.7% of patients classified with a score above 26 at 89.9% specificity.

**Supplementary Figure 13:**
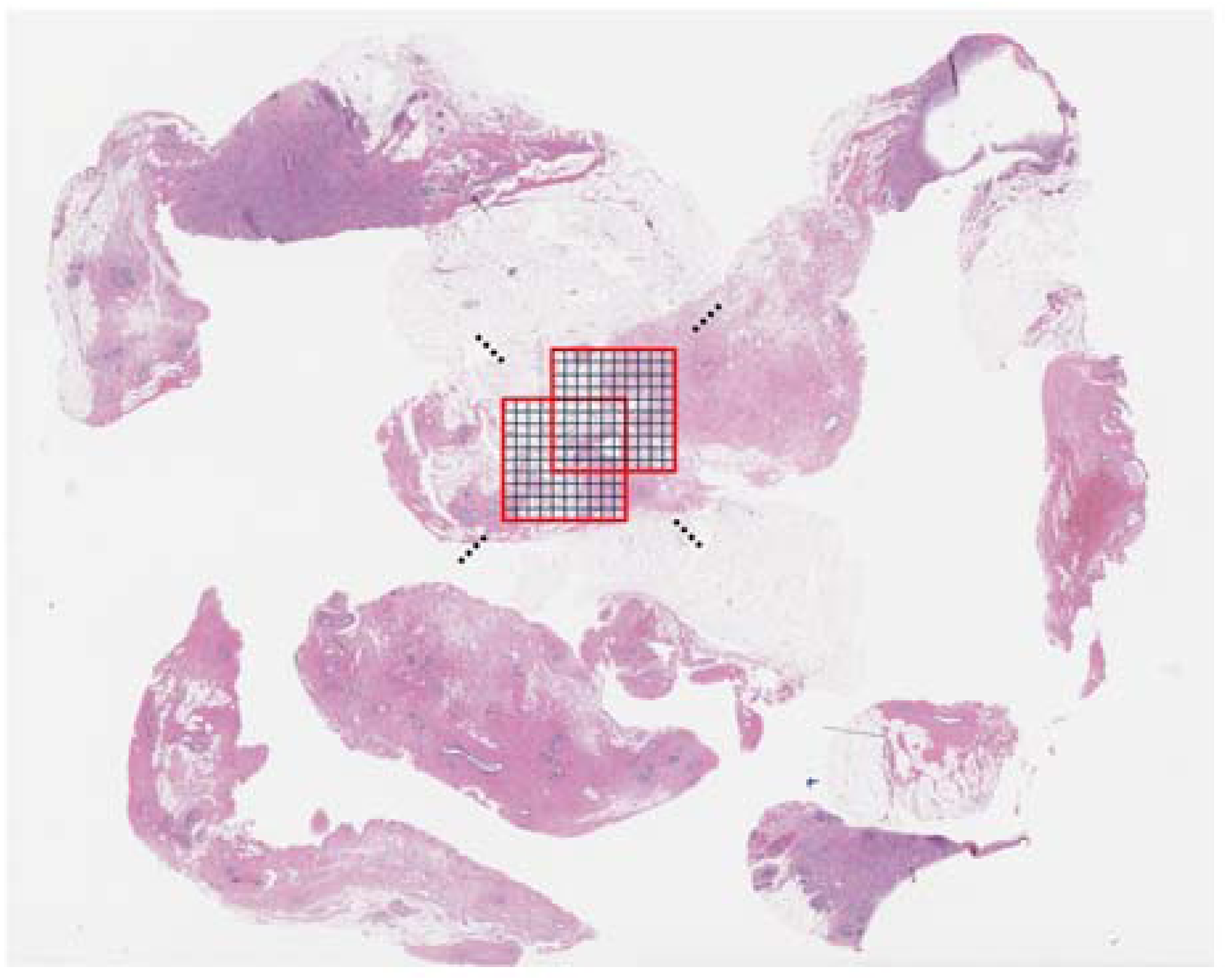
Tissue segmentation and sliding window approach for interpretability analysis. Illustration of the methodology used to generate interpretable heatmaps from H&E whole-slide images. The image shows a breast cancer histopathology slide with a 10×10 tile grid (red squares) overlaid on tumor tissue. Each grid represents a region analyzed as a collective bag of tiles in the multiple instance learning (MIL) framework. The model analyzes these regions with a sliding window approach (stride of 4 tiles) to generate local recurrence AI scores and attention scores. Black dots indicate reference points for the sliding window progression. This approach enables spatial mapping of the model’s decision-making process across the entire tissue specimen, revealing which regions most strongly influence the final risk classification.

