## Supplementary Information for "Deep Learning on Histopathological Images to Predict Breast Cancer Recurrence Risk and Chemotherapy Benefit"

**Supplementary Table 1: Overview of study cohorts and patient inclusion.**

| Dataset | Year of diagnosis | Total Collected | | Included in analysis | | Available Oncotype DX scores? |
|---|---|---|---|---|---|---|
| | | Slides | Patients | Slides | Patients | |
| **For tuning and validation** | | | | | | |
| TAILORx | 2006-2010 | 9,619 | 10,273 | 9,383 | 8,284 | Yes |
| Tune | | | | 6,680 | 5,877 | Yes |
| Validation | | | | 2,703 | 2,407 | Yes |
| **For external validation** | | | | | | |
| Carmel | 2015-2022 | 1,022 | 569 | 1,013 | 565 | Yes |
| Haemek | 2014-2022 | 202 | 169 | 199 | 156 | Yes |
| Sheba | 2014-2020 | 697 | 437 | 676 | 427 | Yes |
| UCMC | 2006-2020 | 601 | 511 | 568 | 490 | Yes |
| ABCTB | 2006-2015 | 3,024 | 2,552 | 2,080 | 1,762 | No |
| TCGA | 1988-2013 | 3,114 | 1,094 | 1,463 | 594 | Estimated from GE |
| **For calibration and additional experiments** | | | | | | |
| Carmel-Calibration | 2017-2021 | 5,790 | 1,616 | 1,176 | 1,176 | No |
| Haemek-Calibration | 2017-2021 | 1,367 | 591 | 327 | 327 | No |
| **Total** | | 25,436 | 17,812 | 16,885 | 13,781 | |

*Summary of all cohorts used in the study, including TAILORx and the external validation cohorts. For each cohort, we present the years of diagnosis, the total number of collected slides and patients, and the final numbers included in the analysis. The table is organized into three sections: (1) TAILORx data used for tuning using cross-validation and for validation on a held-out validation set. (2) External validation cohorts - comprising three Israeli medical centers (Carmel, Haemek, Sheba), two from the United States (UCMC, TCGA), and one from Australia (ABCTB). (3) Additional calibration cohorts - used for validating the distribution-matching calibration method and for calibrating the model for Carmel and Haemek. For TAILORx, Carmel, Haemek, Sheba, and UCMC, Oncotype DX scores were available from clinical testing. For TCGA, RS values were computationally estimated from RNA sequencing gene expression (GE) data. Detailed inclusion/exclusion criteria and quality control steps are shown in Supplementary Figure 1.*

**Supplementary Table 2: Clinicopathological characteristics across study cohorts.**

| Characteristic | TAILORx (N = 8284) | Carmel (N = 565) | Haemek (N = 156) | Sheba (N = 427) | UCMC (N =490) | TCGA (N =594 ) | ABCTB (N =1762 ) |
|---|---|---|---|---|---|---|---|
| RS mean | 18.77±10.01 | 16.14±9.23 | 18.27±10.47 | 19.37±9.86 | 18.40±9.72 | 19.36±6.97 | - |
| RS risk group — no. /total no. (%) | | | | | | | |
| Low (RS < 11) | 1328/8284 (16) | 151/565 (27) | 30/156 (19) | 57/427 (13) | 81/417 (19) | 64/592 (11) | - |
| Intermediate (11 ≤ RS < 26) | 5558/8284 (67) | 332/565 (59) | 98/156 (63) | 282/427 (66) | 261/417 (63) | 429/592 (72) | - |
| High (26 ≤ RS) | 1398/8284 (17) | 82/565 (15) | 28/156 (18) | 88/427 (21) | 75/417 (18) | 99/592 (17) | - |
| Median age (range) — yr | 56 (23–89) | 67 (30–92) | 59.5 (27–87) | 57 (26–83) | 56 (31–89) | 59 (26–90) | 60 (26–95) |
| Age ≤50 yr — no. (%) | 2590 (31) | 66 (12) | 40 (26) | 140 (33) | 155 (32) | 161 (27) | 461 (26) |
| Menopausal status — no. (%) | | | | | | | |
| Premenopausal | 2812 (34) | - | - | - | - | - | - |
| Postmenopausal | 5472 (66) | - | - | - | - | - | - |
| Tumor size — cm | | | | | | | |
| Median (IQR) | 1.5 (1.2–2.1) | - | 1.7 (1.2–2.4) | 1.6 (1.2–2.2) | 1.7 (1.3–2.5) | 3.5 (1.5–3.5) | 2.0 (1.4–3.0) |
| Mean | 1.75±0.82 | - | 1.99±1.26 | 1.87±1.08 | 2.19±1.78 | 3.14±1.24 | 2.61±2.00 |
| Grade — no./total no. (%) | | | | | | | |
| Low | 2099/7931 (26) | 130/510 (25) | 34/135 (25) | 18/378 (5) | 76/490 (16) | 94/570 (16) | 373/1753 (21) |
| Intermediate | 4383/7931 (55) | 332/510 (65) | 78/135 (58) | 266/378 (70) | 325/490 (66) | 311/570 (55) | 877/1753 (50) |
| High | 1449/7931 (18) | 46/510 (9) | 23/135 (17) | 94/378 (25) | 89/490 (18) | 165/570 (29) | 503/1753 (29) |
| ER expression — no./total no. (%) | | | | | | | |
| Negative | 45/8284 (1) | 1/565 (<1) | 0/156 (0) | 0/427 (0) | 1/490 (<1) | 11/594 (2) | 35/1761 (2) |
| Positive | 8095/8284 (99) | 564/565 (>99) | 156/156 (100) | 427/427 (100) | 489/490 (>99) | 583/594 (98) | 1726/1761 (98) |
| PR expression — no./total no. (%) | | | | | | | |
| Negative | 803/7952 (10) | 85/565 (15) | 21/153 (13.7) | 78/425 (18) | 51/490 (10) | 80/592 (14) | 186/1748 (11) |
| Positive | 7149/7952 (90) | 480/565 (85) | 132/153 (86.3) | 347/425 (81) | 439/490 (90) | 512/592 (86) | 1562/1748 (89) |
| Clinical risk — no./total no. (%) | | | | | | | |
| Low | 5531/7946 (70) | - | 68/126 (54) | 191/382 (50) | 227/483 (47) | 96/518 (19) | 600/1671 (36) |
| High | 2415/7946 (30) | - | 58/126 (46) | 191/382 (50) | 256/483 (53) | 422/518 (81) | 1071/1671 (64) |
| Primary surgery — no. (%) | | | | | | | |
| Mastectomy | 2258/8140 (28) | - | - | - | - | - | - |
| Breast conservation | 5882/8140 (72) | - | - | - | - | - | - |
| Adjuvant chemotherapy — no. (%) | | | | | | | |
| Yes | 3631 (44) | - | - | - | 131 (27) | - | - |
| No | 4653 (56) | - | - | - | 359 (73) | - | - |
| Tumor Subtype — no./total no. (%) | | | | | | | |
| IDC | - | 483/562 (86) | 132/156 (85) | 345/418 (83) | 352/490 (72) | 396/594 (67) | 1368/1762 (78) |
| ILC | - | 68/562 (12) | 19/156 (12) | 59/418 (14) | 79/490 (16) | 141/594 (24) | 289/1762 (16) |
| Other | - | 11/562 (2) | 5/156 (3) | 14/418 (3) | 59/490 (12) | 57/594 (10) | 105/1762 (6) |
| Biopsy Type — no./total no. (%) | | | | | | | |
| Core needle biopsy | - | 52/564 (9) | 25/155 (16) | - | - | - | - |
| Surgical specimen | - | 512/564 (91) | 130/155 (84) | - | - | - | - |
| Gender — no./total no. (%) | | | | | | | |
| Female | 8284/8284 (100) | 560/562 (>99) | - | 425/427 (>99) | 487/490 (99) | 589/594 (99) | 1762/1762 (100) |
| Male | 0/8284 (0) | 2/562 (<1) | - | 2/427 (<1) | 3/490 (1) | 5/594 (1) | 0/1762 (0) |
| Nodal status — no./total no. (%) | | | | | | | |
| Negative | 8284/8284 (100) | - | 105/147 (71) | 322/427 (75) | 358/482 (74) | 244/537 (45) | 855/1670 (51) |
| Positive | 0/8284 (0) | - | 42/147 (29) | 105/427 (25) | 124/482 (26) | 293/537 (55) | 815/1670 (49) |

*Comparison of patient and tumor characteristics across all study cohorts. For continuous variables (age, tumor size, RS), data are presented as means ±SD or median (range) where appropriate. For categorical variables, data are shown as the number (percentage) of patients. Clinical risk stratification follows MINDACT trial criteria: low-risk includes node-negative cases with either low-grade tumors ≤3 cm, intermediate-grade tumors ≤2 cm, high-grade tumors ≤1 cm, or node-positive cases with low-grade tumors ≤2 cm; high-risk encompasses all other cases with complete grade, size, and nodal status data. Missing data fields are indicated by dashes (-). Some characteristics were not consistently available across all cohorts, reflecting real-world data collection practices. Tumor subtypes are classified as IDC (invasive ductal carcinoma), ILC (invasive lobular carcinoma), or other histological types. IQR: interquartile range.*

**Supplementary Table 3: Performance metrics of the AI multimodal model across all cohorts, for identifying high genomic risk (RS≥26).**

a. AUC performance

| Cohort | Patients in analysis | AUC (95% CI) |
|---|---|---|
| TAILORx-CV | 5877 | 0.897 (0.888–0.906) |
| TAILORx-Validation | 2407 | 0.898 (0.879–0.913) |
| Carmel | 565 | 0.903 (0.864–0.930) |
| Sheba | 427 | 0.858 (0.809–0.898) |
| Haemek | 156 | 0.866 (0.793–0.924) |
| UCMC | 417 | 0.871 (0.822–0.908) |
| TCGA | 592 | 0.832 (0.779–0.872) |

b. %Patients, Sensitivity, and Negative predictive Value for AI threshold 16

| Cohort | Patients in analysis | AI score < 16 | | | |
|---|---|---|---|---|---|
| | | % Patients | Sensitivity (95% CI) | Specificity (95% CI) | NPV (95% CI) |
| TAILORx-CV | 5877 | 44.9% | 0.949 (0.934–0.961) | 0.531 (0.526–0.538) | 0.980 (0.974 - 0.985) |
| TAILORx-Validation | 2407 | 45.6% | 0.948 (0.922–0.970) | 0.532 (0.524–0.541) | 0.982 (0.973–0.990) |
| Carmel | 565 | 49.6% | 0.951 (0.901–0.990) | 0.571 (0.554–0.592) | 0.986 (0.971–0.996) |
| Sheba | 327 | 30.6% | 0.985 (0.938–1.000) | 0.378 (0.359–0.401) | 0.990 (0.960–1.000) |
| Haemek | 156 | 42.3% | 0.964 (0.857–1.000) | 0.508 (0.472–0.547) | 0.985 (0.939–1.000) |
| UCMC | 317 | 38.8% | 0.982 (0.937–1.000) | 0.467 (0.445–0.493) | 0.992 (0.967–1.000) |
| TCGA | 492 | 44.7% | 0.920 (0.853–0.967) | 0.526 (0.505–0.548) | 0.968 (0.941–0.986) |

c. %Patients, Specificity, and Positive predictive Value for AI threshold 26

| Cohort | Patients in analysis | AI score ≥ 26 | | | |
|---|---|---|---|---|---|
| | | % Patients | Sensitivity (95% CI) | Specificity (95% CI) | PPV (95% CI) |
| TAILORx-CV | 5877 | 13.2% | 0.559 (0.533–0.582) | 0.957 (0.952–0.962) | 0.732 (0.692–0.764) |
| TAILORx-Validation | 2407 | 12.0% | 0.540 (0.504–0.587) | 0.959 (0.953–0.968) | 0.716 (0.666–0.780) |
| Carmel | 565 | 11.5% | 0.549 (0.467–0.645) | 0.959 (0.943–0.980) | 0.692 (0.559–0.854) |
| Sheba | 327 | 14.1% | 0.462 (0.364–0.566) | 0.939 (0.915–0.962) | 0.652 (0.502–0.783) |
| Haemek | 156 | 10.3% | 0.357 (0.222–0.508) | 0.953 (0.927–0.989) | 0.625 (0.390–0.908) |
| UCMC | 317 | 14.5% | 0.518 (0.419–0.645) | 0.935 (0.914–0.962) | 0.630 (0.494–0.791) |
| TCGA | 492 | 16.7% | 0.471 (0.392–0.574) | 0.899 (0.881–0.925) | 0.500 (0.399–0.626) |

*Summary of AI multimodal model performance across the cohorts, presented in three complementary analyses: (a) Discrimination performance measured by area under the ROC curve (AUC) with 95% confidence intervals (CI) for identifying high genomic risk (RS≥26). TAILORx-CV refers to the average performance of the five cross-validation AI models on their corresponding validation folds, in the TAILORx tuning set. For the external cohorts, patients in this analysis also included patients used for calibration since calibration does not affect ranking-based metrics. (b) Low-risk classification performance (AI<16), reporting the percentage of patients classified as low AI risk, sensitivity, specificity, and negative predictive value (NPV) with 95% CIs for identifying RS≥26. This threshold represents patients who could potentially avoid chemotherapy. (c) High-risk classification performance (AI≥26), showing the percentage of patients classified as high AI risk, sensitivity, specificity, and positive predictive value (PPV) with 95% CIs for identifying RS≥26. This threshold represents patients who might benefit from immediate chemotherapy initiation. Calibration-set patients (n=100 each) were excluded from analyses in (b) and (c) for UCMC, Sheba, and TCGA to ensure an unbiased evaluation of threshold-dependent metrics. AI: artificial intelligence, representing the multimodal model.*

**Supplementary Table 4: Comparative analysis of foundation model versus traditional supervised learning approaches.**

| | Supervised AUC (95% CI) | Foundation AUC (95% CI) |
|---|---|---|
| TAILORx-CV (N = 5877) | 0.867 (0.839–0.889) | 0.880 (0.847–0.902) |
| TAILORx-Validation (N = 2407) | 0.851 (0.829–0.872) | 0.876 (0.857–0.895) |
| Carmel (N = 565) | 0.699 (0.635–0.762) | 0.859 (0.811–0.895) |
| Sheba (N = 427) | 0.771 (0.712–0.820) | 0.821 (0.767–0.871) |
| Haemek (N = 156) | 0.715 (0.601–0.804) | 0.862 (0.774–0.926) |

*Performance comparison between our SSL-based foundation approach and a fully supervised learning model. Both models were tuned on the TAILORx training set to classify cases as low-risk (RS<26) or high-risk (RS≥26) from H&E images. TAILORx-CV refers to the average performance of the five cross-validation AI models on their corresponding validation folds, in the TAILORx tuning set. The supervised model follows the architecture and training protocol described in [29]. The table presents the area under the ROC curve (AUC) with 95% confidence intervals for both approaches across TAILORx cross-validation, validation set, and all external validation cohorts. Despite having access to the same large-scale tuning data, the foundation model demonstrates consistently superior performance, particularly in external validation, highlighting the benefits of transfer learning from a trained foundation model for robust generalization across different clinical settings.*

**Supplementary Figure 1: Cohort inclusion criteria and patient flow diagrams.**

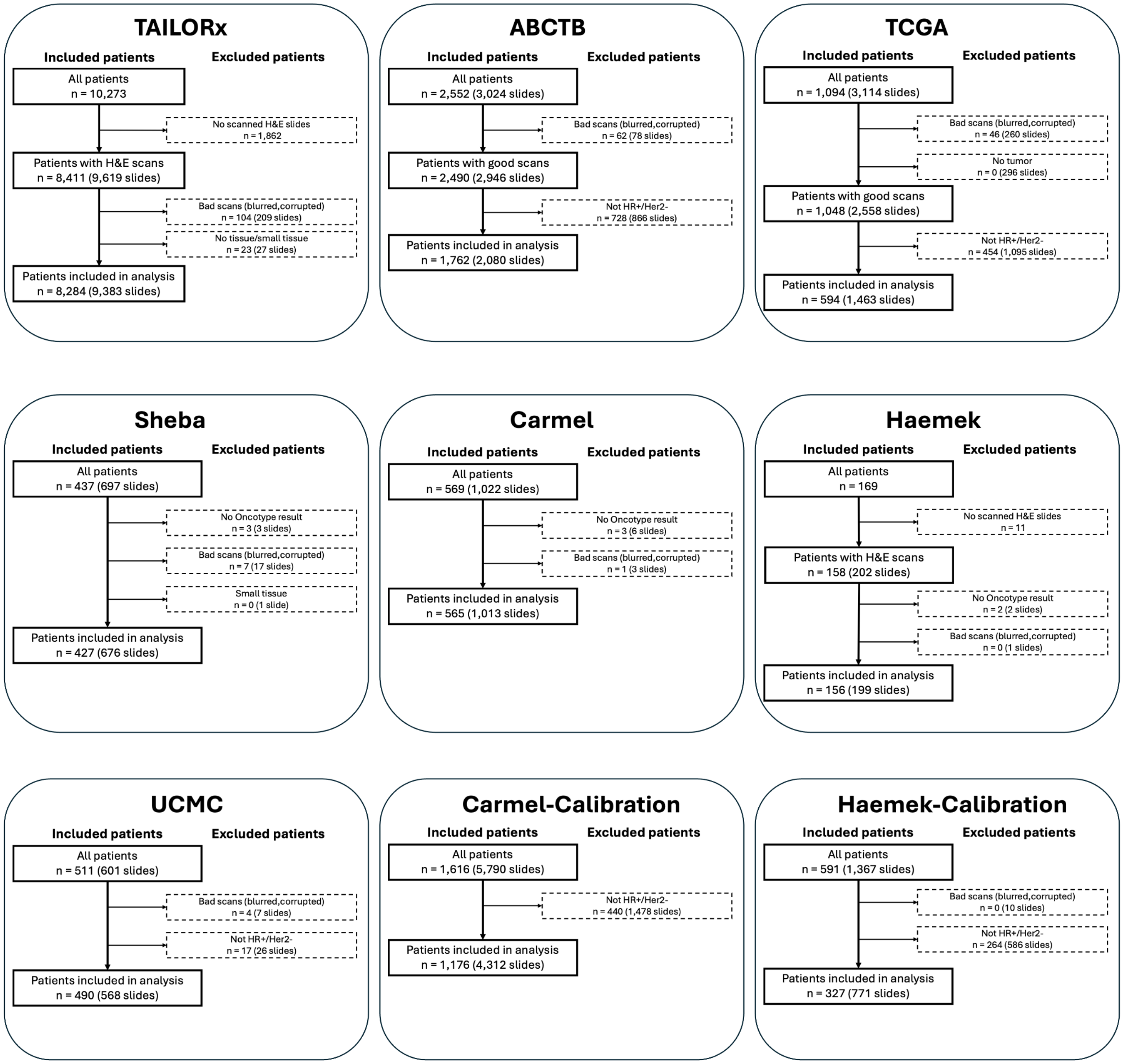


*Flow diagrams showing patient inclusion and exclusion criteria for all study cohorts. For each cohort, we show the initial number of patients and slides collected, followed by sequential exclusion steps with specific reasons and numbers. Starting with TAILORx (N=10,273 patients), we detail exclusions due to missing H&E slides and quality control failures (blurred/corrupted scans, tissue-deficient slides). For the external validation cohorts, we show a similar workflow for Carmel (N=569), Sheba (N=437), Haemek (N=169), UCMC (N=511), ABCTB (N=2,552), and TCGA (N=1,094). The calibration cohorts (Carmel-Calibration and Haemek-Calibration) had different inclusion criteria as they did not require matched RS data. Quality control criteria were consistently applied across all cohorts, focusing on slide scan quality and tissue adequacy. Final numbers of included patients and slides are provided for each cohort. HR: hormone receptor status.*

**Supplementary Figure 2: Performance evaluation of the deep learning models on the TAILORx cross-validation.**

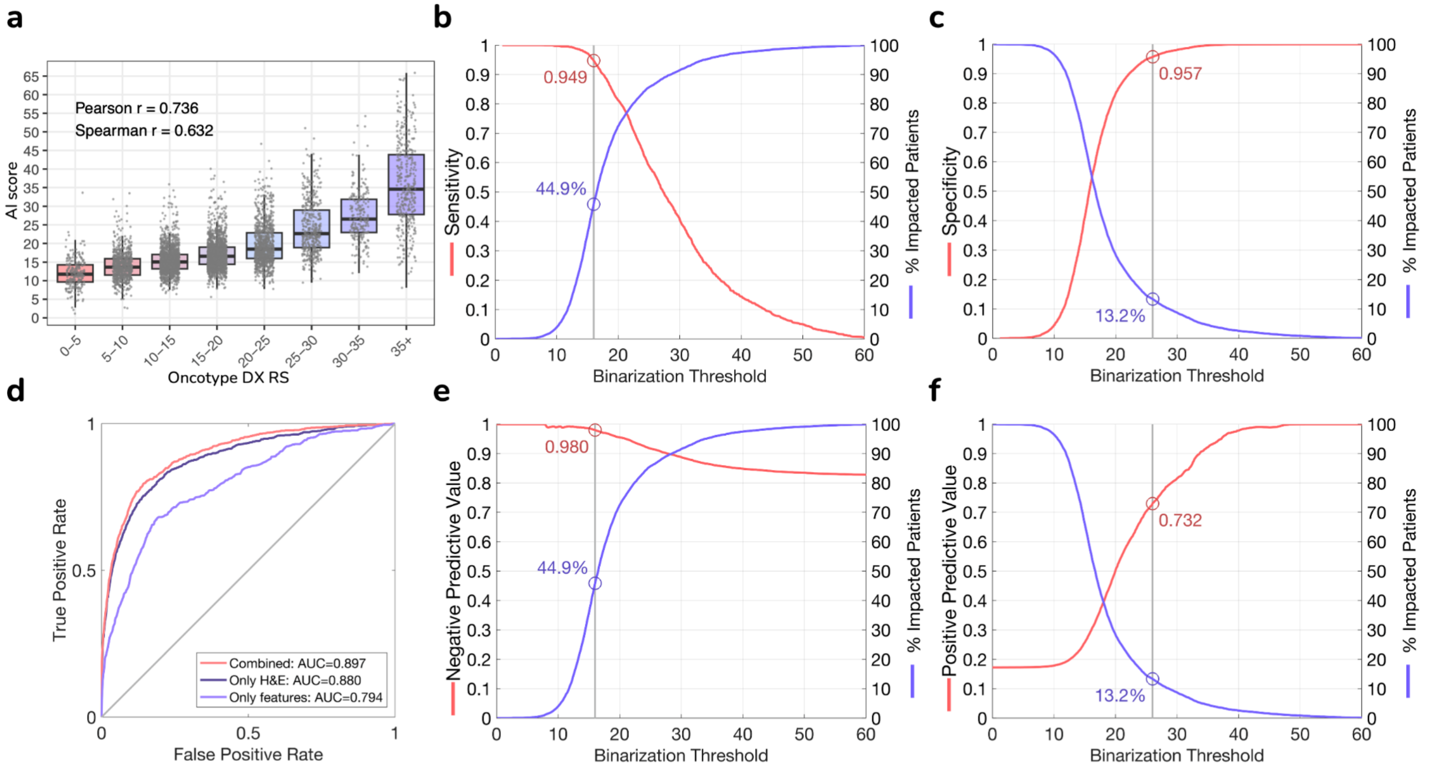


*Model performance assessment during five-fold cross-validation on the TAILORx cohort (n=5,877 patients).* ***(a)*** *Distribution of the AI multimodal model's scores versus Oncotype DX RS on the TAILORx, showing high correlation (Pearson r=0.736, Spearman r=0.652).* ***(b)*** *Clinical utility assessment for low AI risk identification, showing sensitivity and proportion of impacted patients across AI score thresholds. Using a low classification threshold of 16, the model classifies 44.9% of patients as low AI risk with 94.9% sensitivity for identifying RS≥26.* ***(c)*** *Clinical utility assessment for high-risk identification, showing specificity and proportion of impacted patients across AI score thresholds. Using a high classification threshold of 26, the model classifies 13.2% of patients as high AI risk with 95.7% specificity for identifying RS≥26.* ***(d)*** *Receiver operating characteristic (ROC) curves for identifying RS≥26, comparing the multimodal model combining both H&E images and clinicopathologic variables (AUC=0.897), the image model using only H&E images (AUC=0.880), and the clinicopathologic model using only the features (AUC=0.794).* ***(e-f)*** *Negative Predictive Value (NPV) and Positive Predictive Value (PPV) for the low and high classification thresholds.*

**Supplementary Figure 3: Extended survival analysis when including cross-validation patients.**

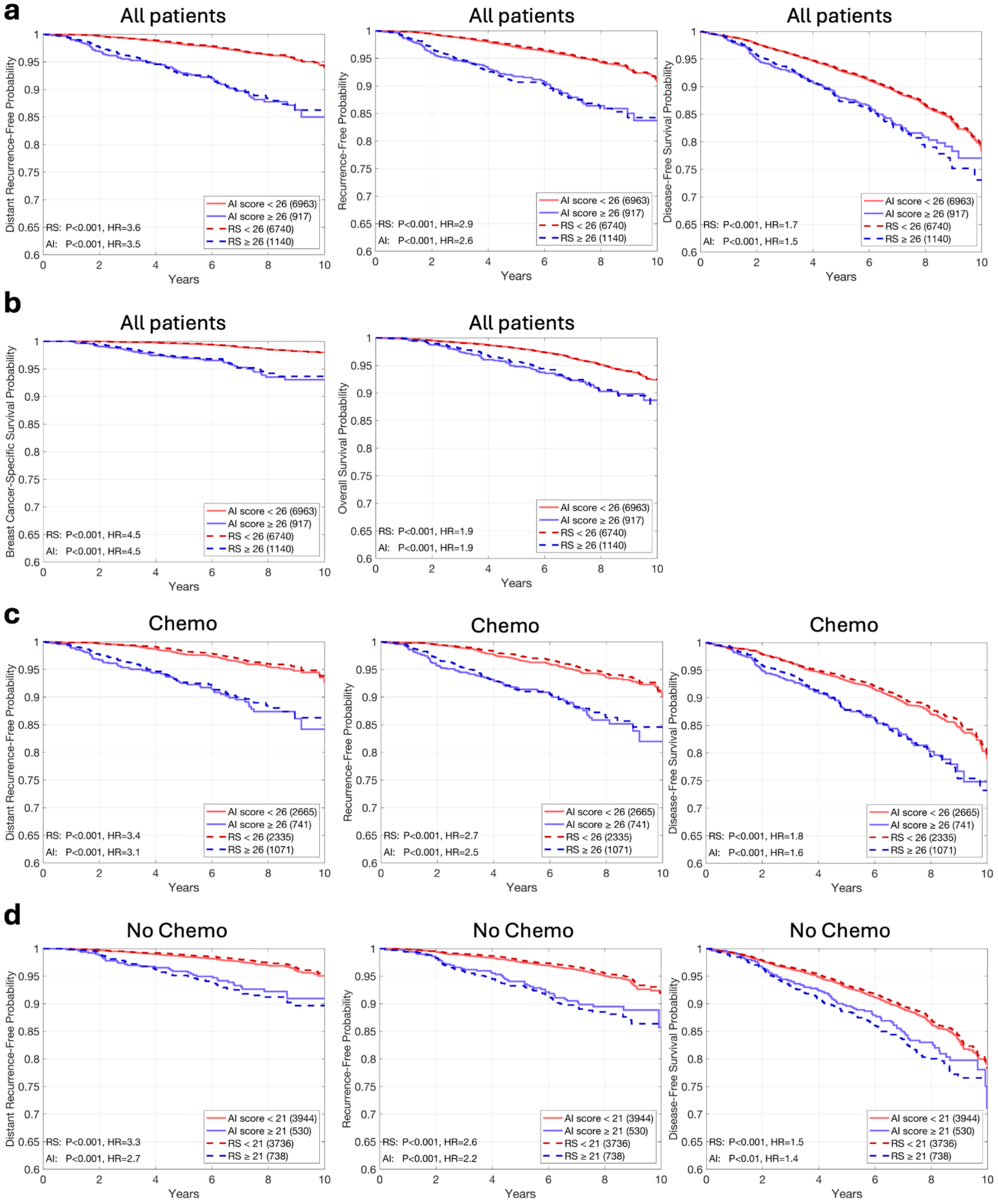

*Kaplan-Meier plots comparing patient stratification using recurrence scores (RS) versus AI scores in the entire TAILORx cohort, including both validation set and cross-validation patients.* ***(a)*** *Overall cohort analysis shows distant recurrence-free interval (DRFI), recurrence-free interval (RFI), and disease-free survival (DFS) probabilities using a stratification threshold of 26.* ***(b)*** *Additional endpoints: breast cancer-specific survival and overall survival.* ***(c)*** *Analysis of the same endpoints in patients who received chemotherapy.* ***(d)*** *Corresponding analysis for patients who did not receive chemotherapy, using a classification threshold of 21. For each subgroup analysis, patient numbers, P values for the stratification, and hazard ratio (HR) values are shown in the legends. The consistent alignment between survival curves derived from RS and AI scores across all analyses demonstrates the robust prognostic ability of the model.*

**Supplementary Figure 4: Extended survival analysis with additional classification thresholds.**

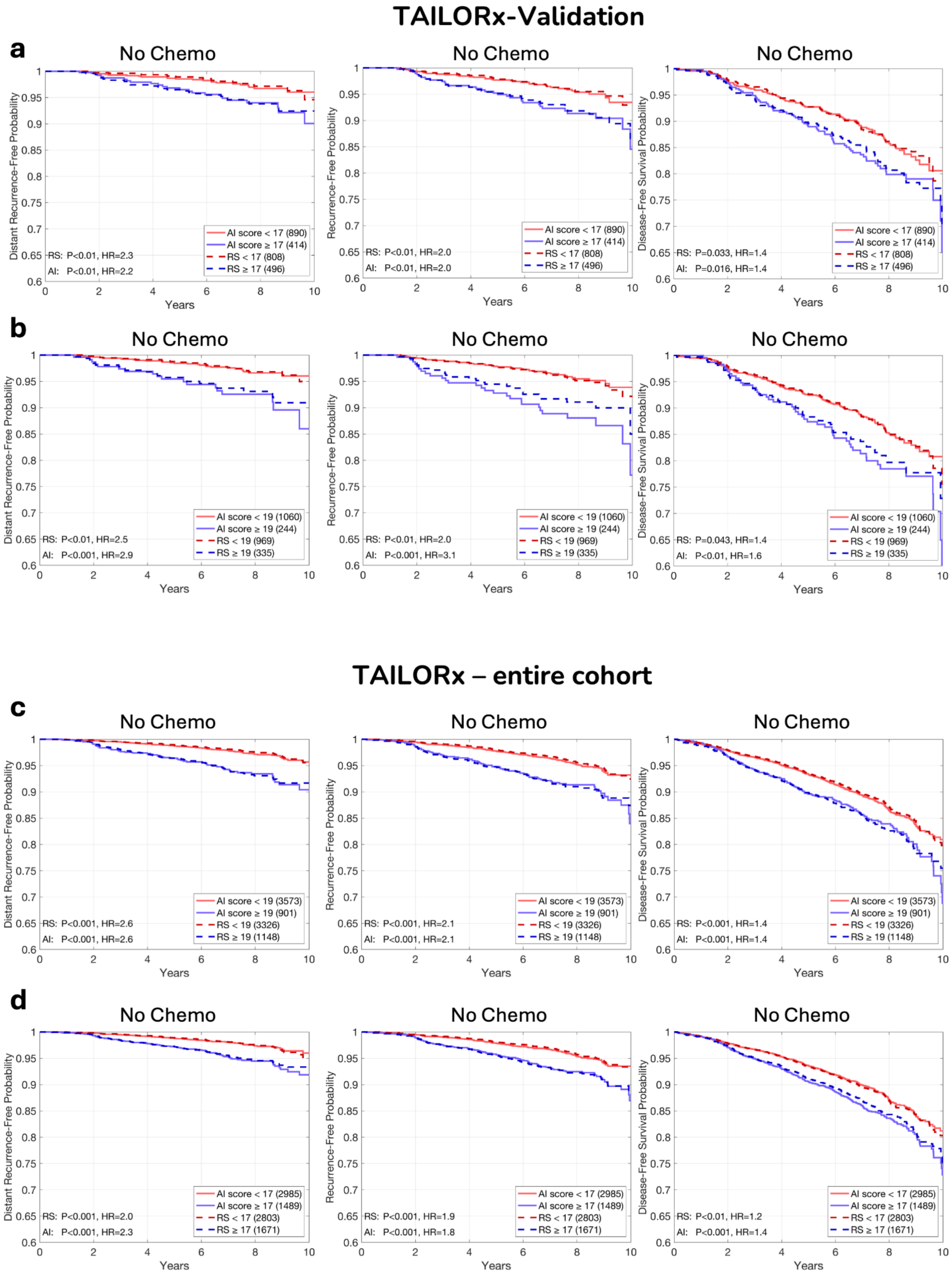


*Kaplan-Meier plots comparing patient stratification using recurrence scores (RS) versus AI scores in TAILORx patients who did not receive chemotherapy. This figure complements Figure 2c and Supplementary Figure 3d by evaluating additional classification thresholds (17 and 19) beyond the primary threshold of 21. For each endpoint (distant recurrence-free interval, recurrence-free interval, and disease-free survival) and threshold value, patient stratification is shown using both RS and AI scores. Each Kaplan-Meier plot includes the number of patients per risk group and hazard ratios (HR) with corresponding P-values from log-rank tests. The consistent performance across multiple thresholds demonstrates the robustness of the AI model's prognostic capability at various decision points.*

**Supplementary Figure 5: Chemotherapy Benefit for Pre- and Post-Menopausals with Intermediate RS: Additional Endpoints**

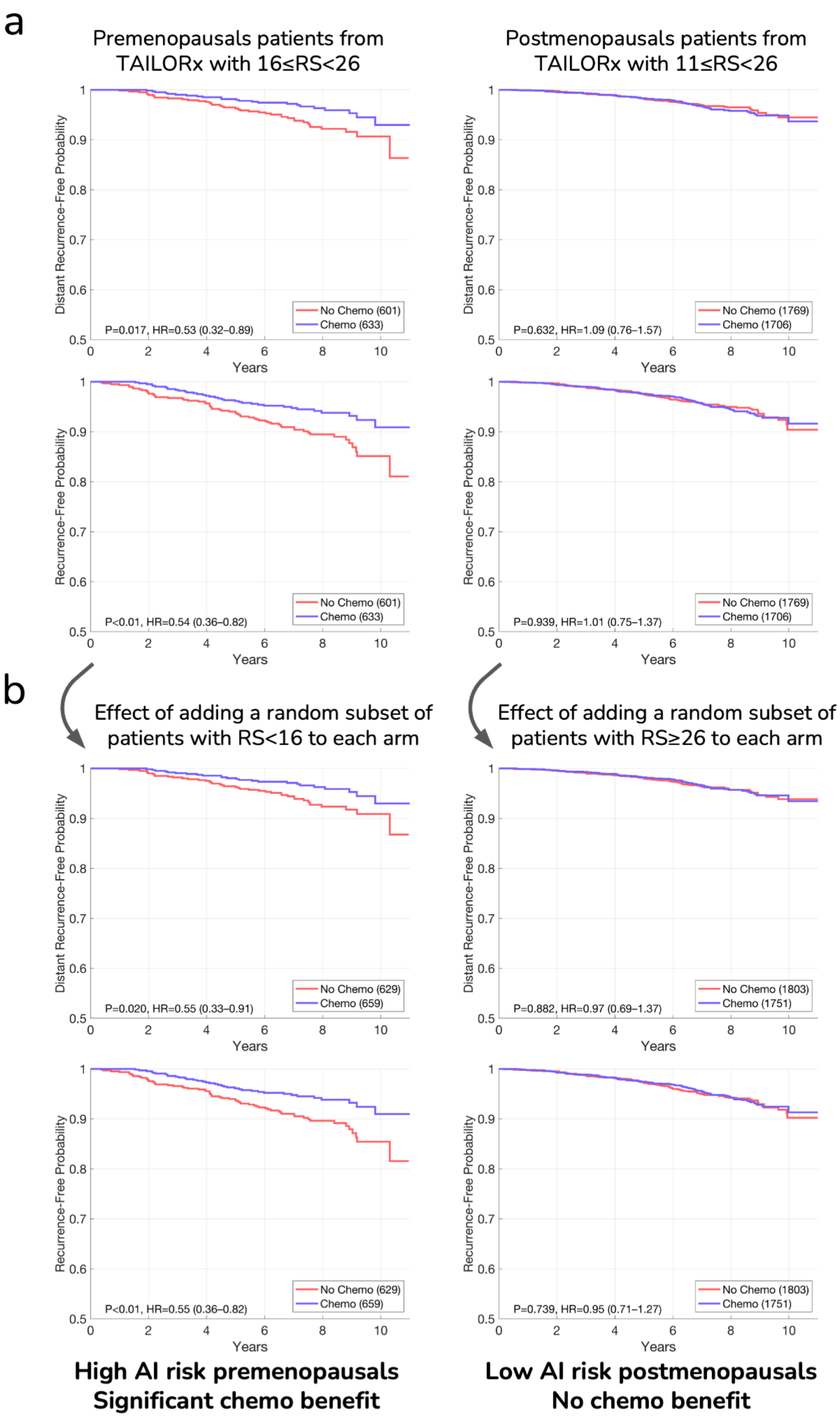


*This figure complements Figure 3 in the main paper, using additional endpoints: distant recurrence-free interval and recurrence-free interval.*

**Supplementary Figure 6: Distribution analysis of genomic and model-estimated recurrence scores.**

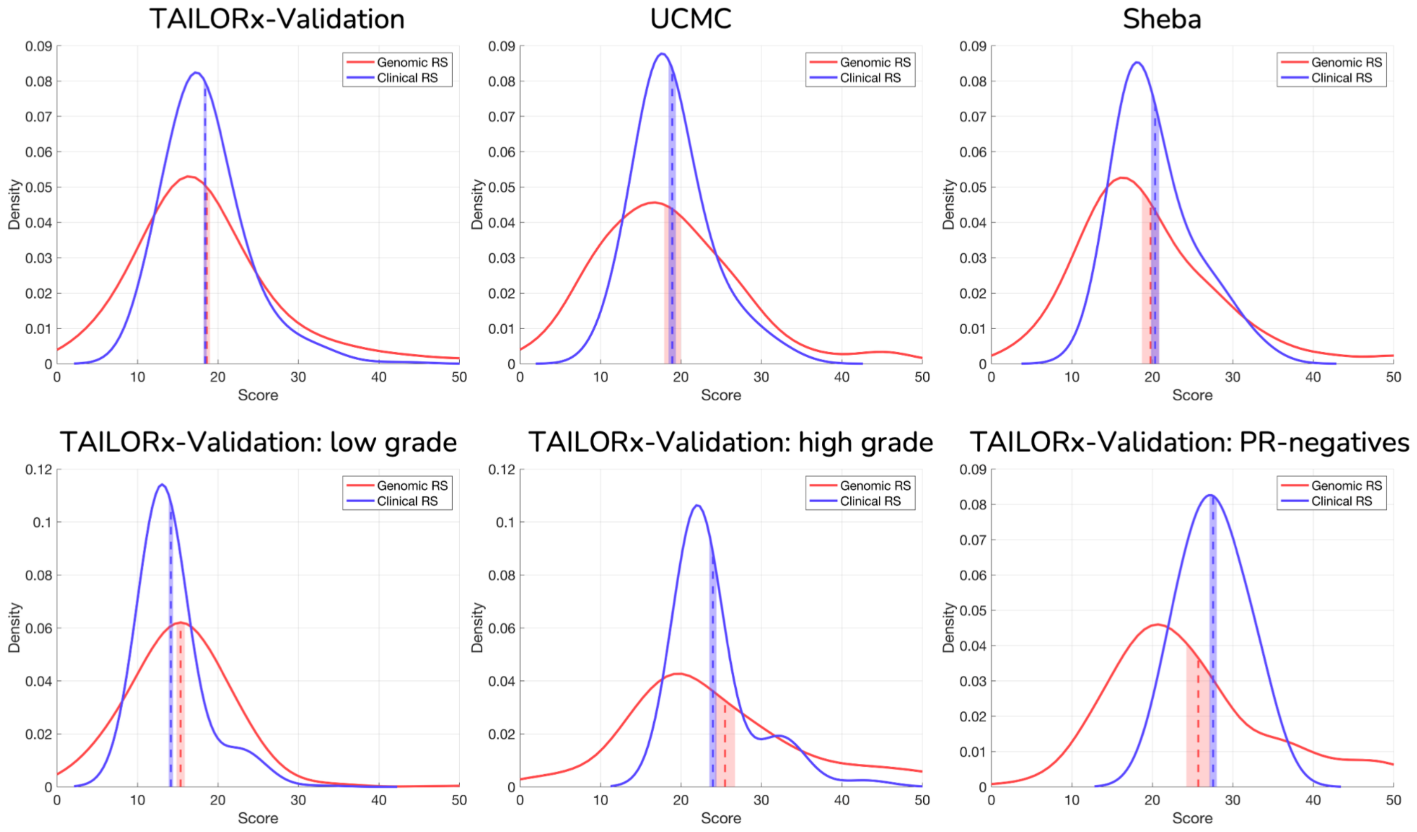


*Probability density distribution analysis comparing Oncotype DX RS ('genomic RS') and estimation of RS using the clinicopathologic model, which was trained using only clinical features ('Clinical RS').* ***Top row:*** *Distribution comparison for the TAILORx validation set (n=2,407), UCMC (n=490), and Sheba (n=427) cohorts.* ***Bottom row:*** *Analysis of specific patient subgroups within the TAILORx validation set, stratified by histological grade (low-grade and high-grade) and PR status (PR-negative). For each plot, means are displayed with their 95% confidence intervals (vertical dashed lines), and kernel density estimation curves show the full distribution of scores. Despite differences in the overall shape of distributions between genomic and estimated RS, the mean values show strong concordance. This analysis demonstrates that while the clinicopathologic model may not accurately infer the RS, it reliably captures its average risk scores across different cohorts and patient subgroups.*

**Supplementary Figure 7: analysis of calibration sample size requirements.**

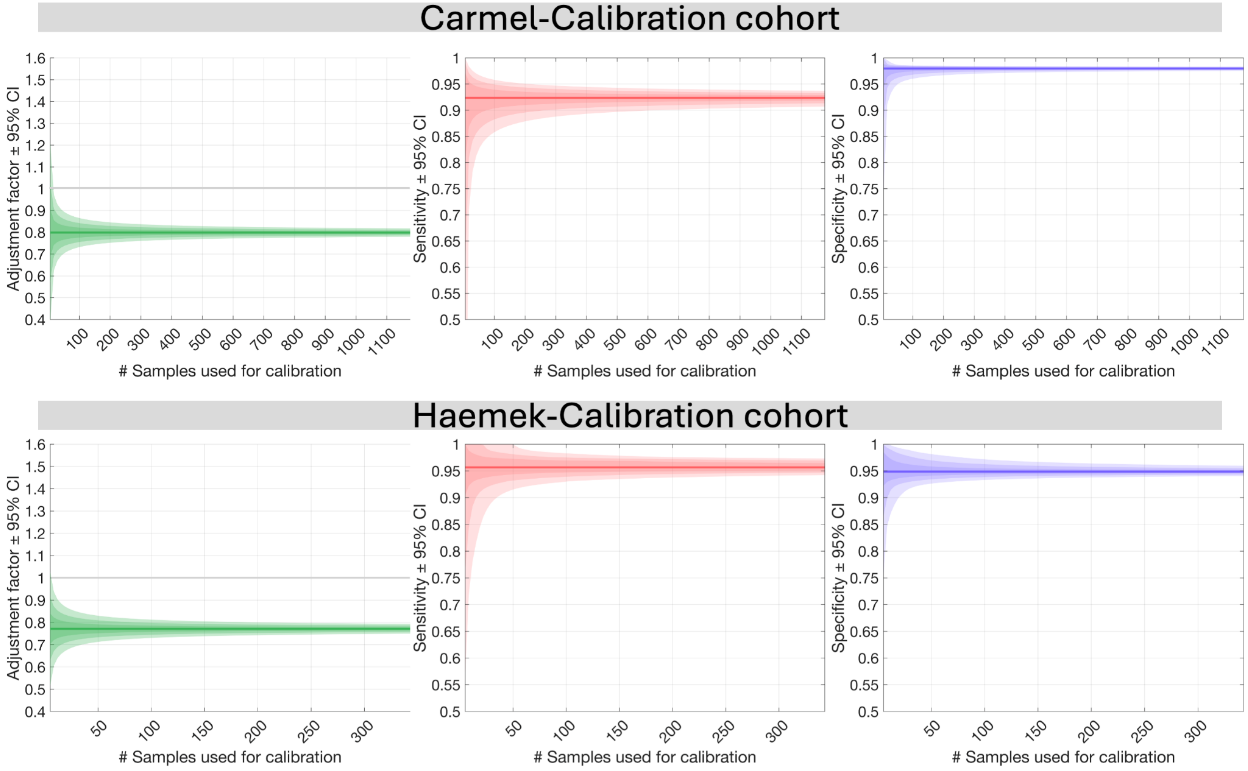


*Investigation of how the number of samples used for calibration affects model performance. Results are shown for two independent calibration cohorts: Carmel-Calibration (n=1,176) and Haemek-Calibration (n=327). For each cohort, three key metrics are analyzed as a function of calibration sample size.* ***Left****: Adjustment factor α with 95% confidence intervals (CI), showing the stability of the calibration coefficient.* ***Middle****: Sensitivity for identifying RS≥26 on Carmel and Haemek cohorts using the low classification threshold (16), along with 95% CIs.* ***Right****: Specificity for identifying RS≥26 on Carmel and Haemek cohorts using the high classification threshold (26), along with 95% CIs. Results demonstrate that the CIs become smaller with more samples used for calibration. We used 100 patients from each cohort for calibrating our multimodal model. The consistency across both calibration cohorts, despite their different sizes and patient characteristics, validates the robustness of our calibration approach. These calibration samples do not require matched genomic testing results, facilitating implementation in resource-limited settings.*

**Supplementary Figure 8: Correlation between predicted and genomic recurrence scores across external validation cohorts.**

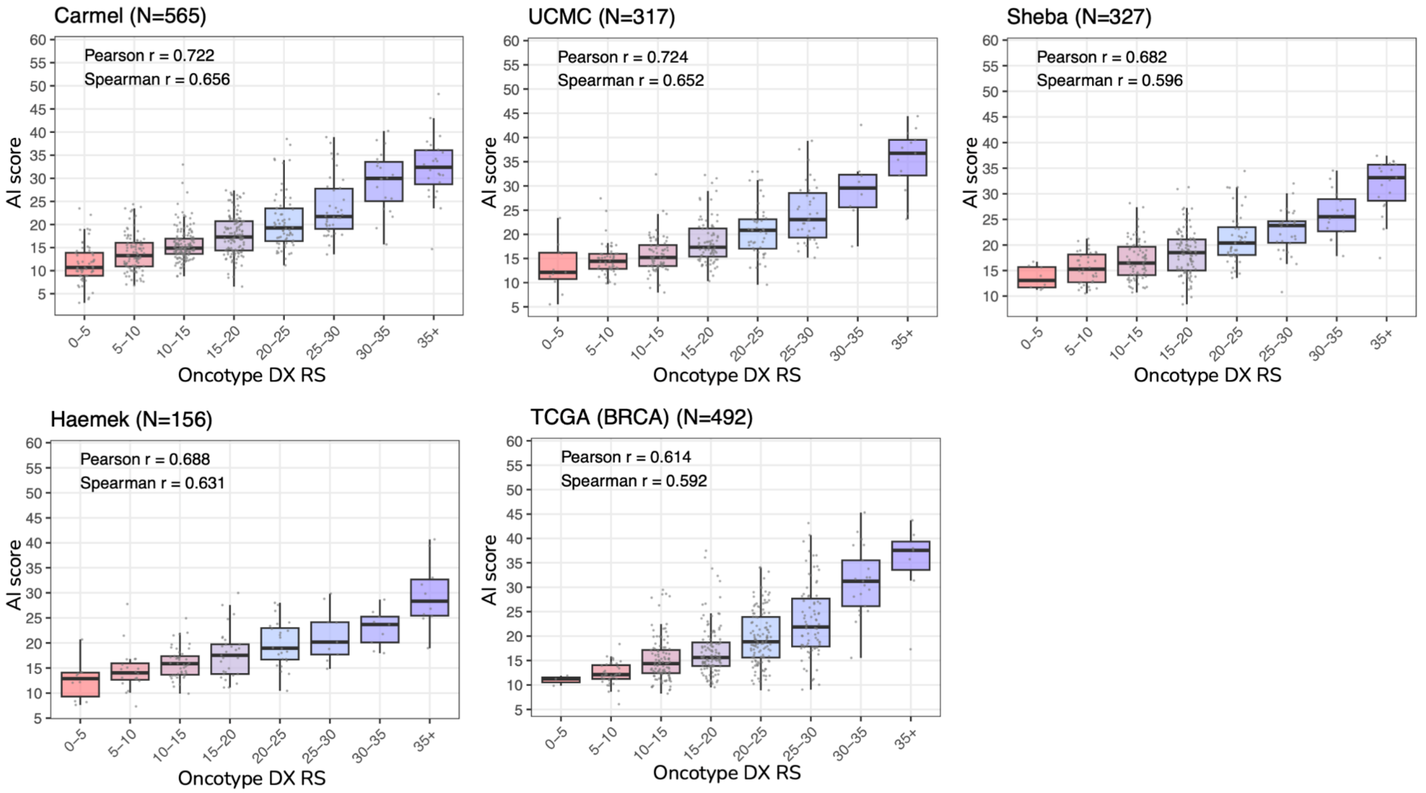


*Box plots demonstrating the relationship between AI scores and Oncotype DX RS values across the five external validation cohorts: Carmel (N=565), UCMC (N=317), Sheba (N=327), Haemek (N=156), and TCGA (N=492). For each cohort, the AI score distributions are shown as box plots grouped by RS ranges, with Pearson (r) and Spearman (ρ) correlation coefficients displayed. The boxes show quartiles (25th, 50th, and 75th percentiles), whiskers extend to 1.5 times the interquartile range, and outliers are shown as individual points. The strong correlation between the AI score and RS values is maintained across all cohorts (Pearson r ranging from 0.688 to 0.754), despite differences in patient populations, timeframes, and laboratory protocols. For TCGA, RS values were estimated from RNA sequencing data, as direct Oncotype DX results were not available.*

**Supplementary Figure 9: Negative and Positive Predictive value analysis of AI model thresholds across validation cohorts.**

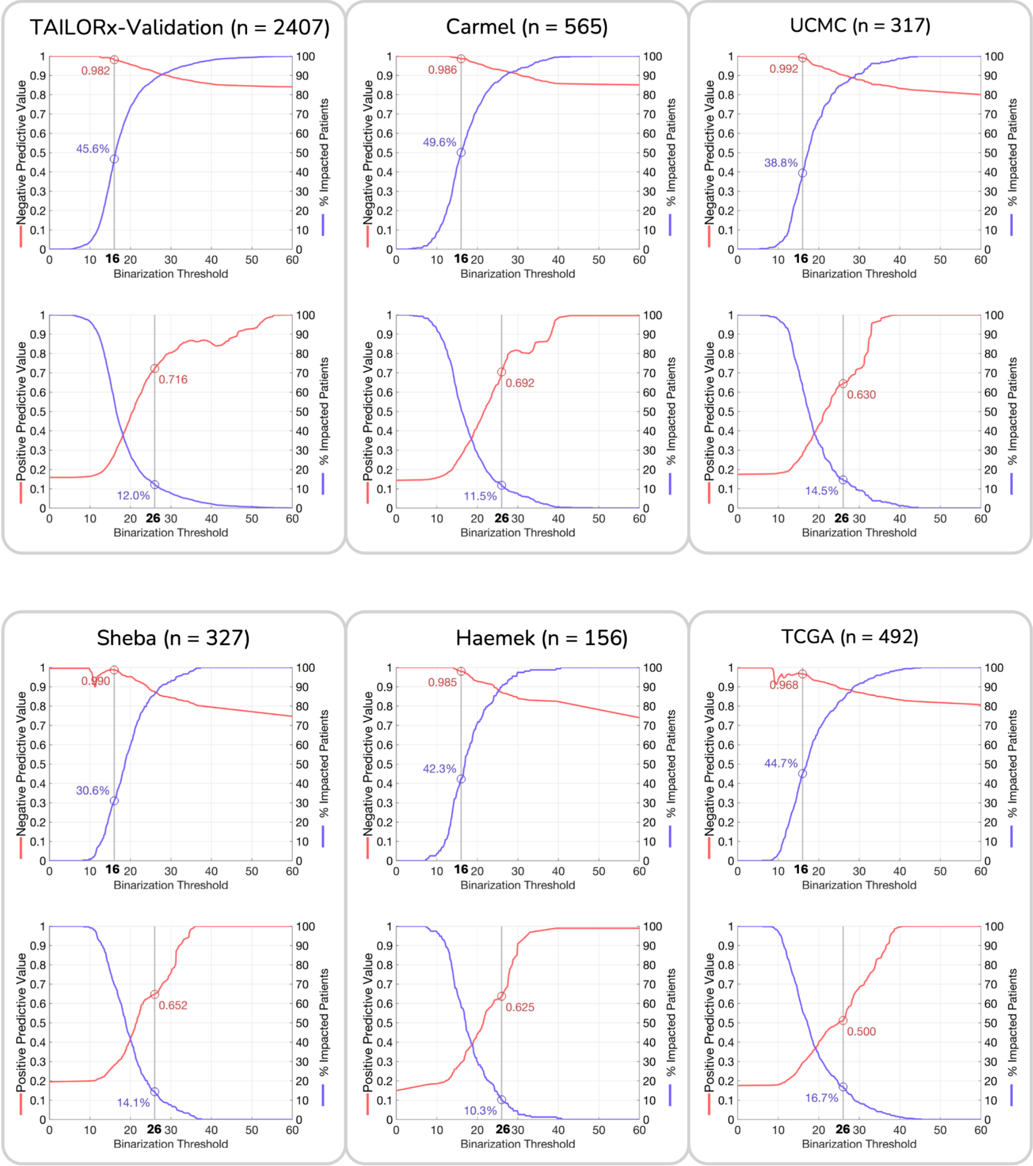


*Assessment of the negative and positive predictive values across all validation cohorts: TAILORx validation set (n=2,407), Carmel (n=565), UCMC (n=317), Sheba (n=327), Haemek (n=156), and TCGA (n=492).* ***Left panels****: Negative predictive value (NPV) analysis for identifying low genomic risk disease (RS<26), showing the relationship between NPV and percentage of patients classified below each threshold, with an operating point at 16 highlighted.* ***Right panels****: Positive predictive value (PPV) analysis for identifying high genomic risk disease (RS≥26), showing the relationship between PPV and the percentage of patients classified above each threshold, with an operating point at 26 highlighted. The consistency of negative and positive predictive values across cohorts demonstrates the robustness of these decision thresholds. The plots show that substantial proportions of patients (30%–50% for low AI risk and 10–17% for high AI risk) can be classified with high confidence across all validation sites. For TCGA, RS values were estimated from gene expression data as direct Oncotype DX results were not available.*

**Supplementary Figure 10: Impact of calibration on AI model performance at Carmel and Sheba Cohorts.**

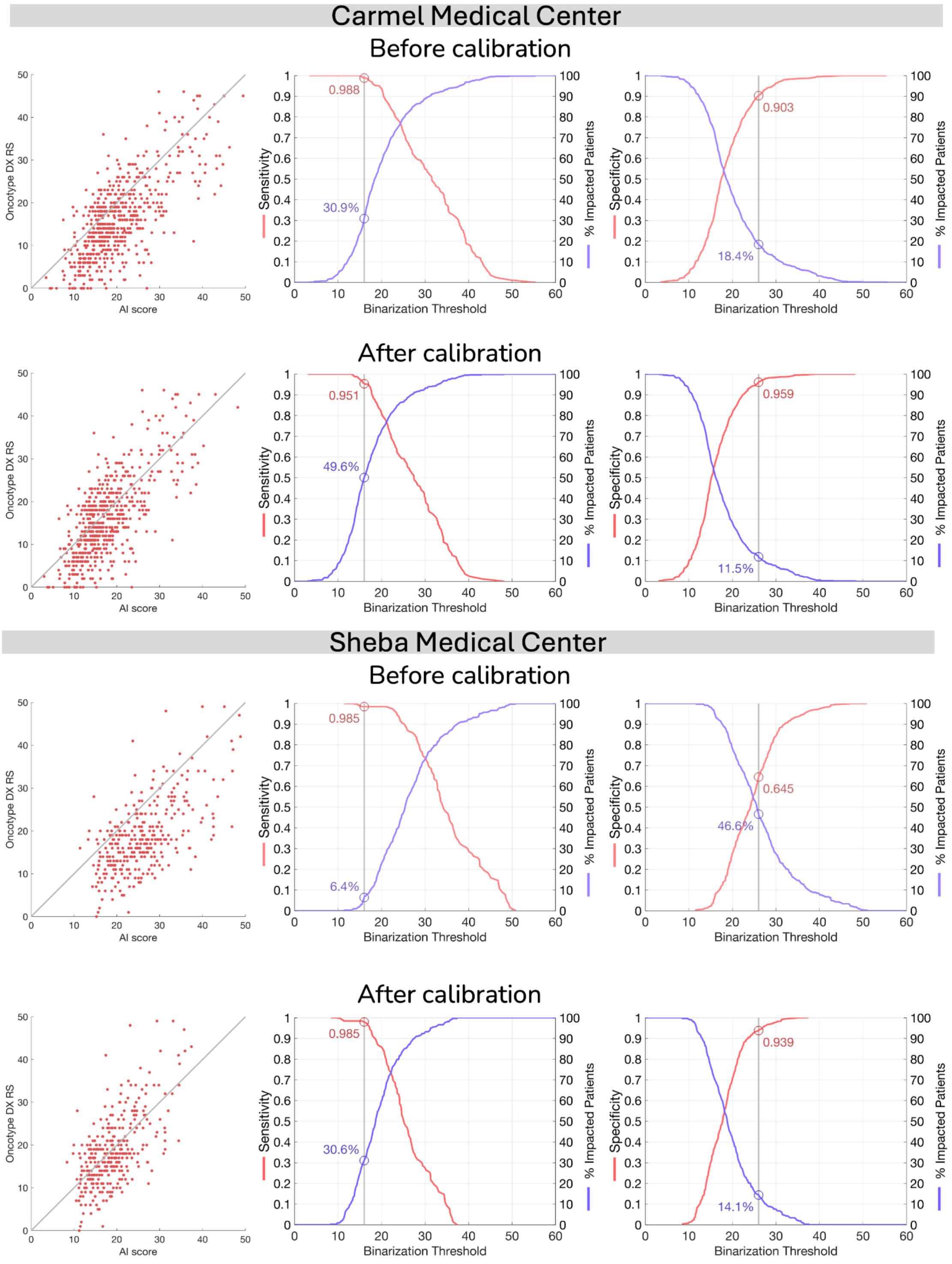


*Comparative analysis of AI scores before and after applying the distribution-matching calibration method at two independent validation sites. Results for both Carmel Medical Center and Sheba Medical Center are presented in three complementary visualizations.* ***Left****: Scatter plots of the AI scores versus Oncotype DX RS, demonstrating correlation patterns.* ***Middle****: Sensitivity for identifying RS≥26 showing the proportion of patients with AI scores below each threshold, with an operating point at 16 indicated.* ***Right****: Specificity for identifying RS≥26 showing the proportion of patients with AI scores above each threshold, with an operating point at 26 highlighted. For*

*each site, the top row shows uncalibrated AI scores, while the bottom row shows results after applying calibration using 100 unlabeled samples. The calibration process maintains ranking-based performance metrics (correlation coefficients) while adjusting AI score distributions to match the development cohort, resulting in more consistent clinical decision thresholds across institutions. The number of analyzed patients (n) and performance metrics are shown for each condition.*

**Supplementary Figure 11: Impact of calibration on AI model performance at Haemek and UCMC Cohorts.**

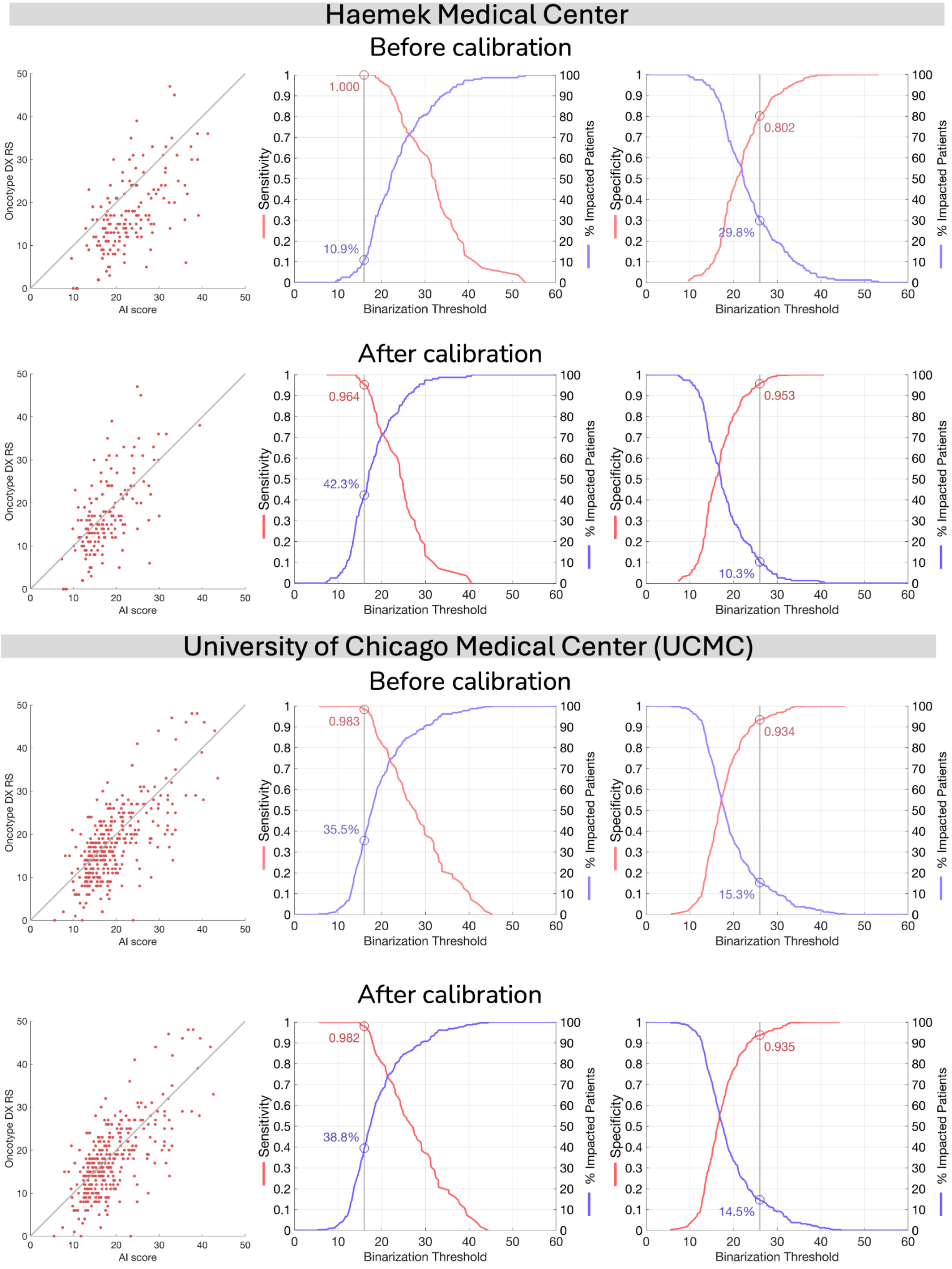


*Comparative analysis of AI scores before and after applying the distribution-matching calibration method at Haemek Hospital and UCMC. Following the same format as Supplementary Figure 10.*

**Supplementary Figure 12: Performance evaluation on TCGA breast cancer cohort using RNA-derived recurrence scores.**

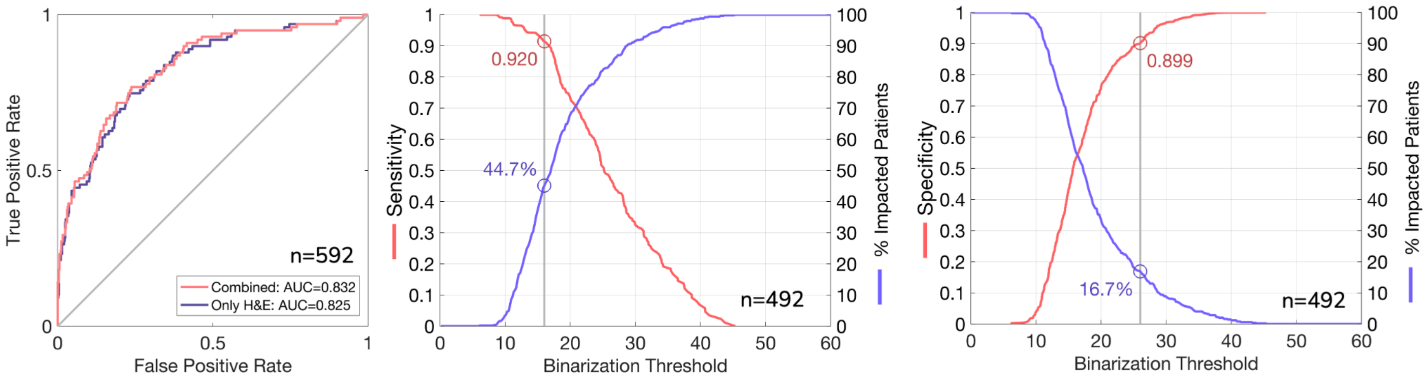


*Performance analysis of the multimodal model on The Cancer Genome Atlas (TCGA) breast cancer cohort, where recurrence scores (RS) were computationally estimated from RNA sequencing data (Methods).* ***Left****: Receiver operating characteristic (ROC) curve for identifying high-risk disease (RS≥26), comparing the performance of the combined multimodal model (AUC=0.832) versus H&E images alone (AUC=0.825).* ***Middle****: Sensitivity analysis showing the proportion of patients classified as low-risk at different thresholds, with 44.7% of patients classified with a score below 16 at 92.0% sensitivity.* ***Right****: Specificity analysis showing the proportion of patients classified as high-risk at different thresholds, with 16.7% of patients classified with a score above 26 at 89.9% specificity.*

**Supplementary Figure 13: Tissue segmentation and sliding window approach for interpretability analysis.**

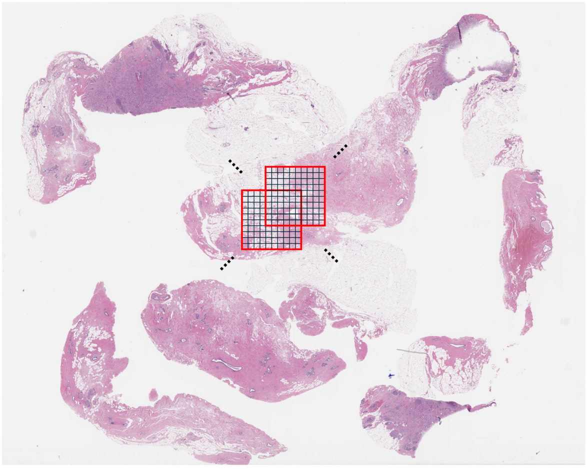

*Illustration of the methodology used to generate interpretable heatmaps from H&E whole-slide images. The image shows a breast cancer histopathology slide with a 10×10 tile grid (red squares) overlaid on tumor tissue. Each grid represents a region analyzed as a collective bag of tiles in the multiple instance learning (MIL) framework. The model analyzes these regions with a sliding window approach (stride of 4 tiles) to generate local recurrence AI scores and attention scores. Black dots indicate reference points for the sliding window progression. This approach enables spatial mapping of the model's decision-making process across the entire tissue specimen, revealing which regions most strongly influence the final risk classification.*
